# Unsupervised machine learning analysis to identify patterns of ICU medication use for fluid overload prediction

**DOI:** 10.1101/2024.03.21.24304663

**Authors:** Kelli Keats, Shiyuan Deng, Xianyan Chen, Tianyi Zhang, John W. Devlin, David J. Murphy, Susan E. Smith, Brian Murray, Rishikesan Kamaleswaran, Andrea Sikora, MRC-ICU Investigator Team

**Author notes:** corresponding author Andrea Sikora, PharmD, MSCR, BCCCP, FCCM, FCCP, 1120 15th Street, HM-118 Augusta, GA 30912.

## Abstract

**INTRODUCTION:** Intravenous (IV) medications are a fundamental cause of fluid overload (FO) in the intensive care unit (ICU); however, the association between IV medication use (including volume), administration timing, and FO occurrence remains unclear.

**METHODS:** This retrospective cohort study included consecutive adults admitted to an ICU ≥72 hours with available fluid balance data. FO was defined as a positive fluid balance ≥7% of admission body weight within 72 hours of ICU admission. After reviewing medication administration record (MAR) data in three-hour periods, IV medication exposure was categorized into clusters using principal component analysis (PCA) and Restricted Boltzmann Machine (RBM). Medication regimens of patients with and without FO were compared within clusters to assess for temporal clusters associated with FO using the Wilcoxon rank sum test. Exploratory analyses of the medication cluster most associated with FO for medications frequently appearing and used in the first 24 hours was conducted.

**RESULTS:** FO occurred in 127/927 (13.7%) of the patients enrolled. Patients received a median (IQR) of 31 (13-65) discrete IV medication administrations over the 72-hour period. Across all 47,803 IV medication administrations, ten unique IV medication clusters were identified with 121-130 medications in each cluster. Among the ten clusters, cluster 7 had the greatest association with FO; the mean number of cluster 7 medications received was significantly greater in patients in the FO cohort compared to patients who did not experience FO (25.6 vs.10.9. p<0.0001). 51 of the 127 medications in cluster 7 (40.2%) appeared in > 5 separate 3-hour periods during the 72-hour study window. The most common cluster 7 medications included continuous infusions, antibiotics, and sedatives/analgesics. Addition of cluster 7 medications to a prediction model with APACHE II score and receipt of diuretics improved the ability for the model to predict fluid overload (AUROC 5.65, p =0.0004).

**CONCLUSIONS:** Using ML approaches, a unique IV medication cluster was strongly associated with FO. Incorporation of this cluster improved the ability to predict development of fluid overload in ICU patients compared with traditional prediction models. This method may be further developed into real-time clinical applications to improve early detection of adverse outcomes.

**KEY POINTS:** *Questions:* Can machine learning detect the presence of time-dependent medication administration patterns that are associated with risk of fluid overload in critically ill patients?

*Findings:* Using unsupervised machine learning, a unique IV medication cluster was identified that, when combined with the APACHE II score and diuretic use, improved the ability to predict fluid overload in ICU patients.

*Meaning:* These findings suggest that machine learning may be an important tool for analyzing IV mediation administration patterns to predict development of fluid overload. Such models may provide insight into areas where medication administration practices could be optimized to mitigate the risk of fluid overload in this patient population.

## INTRODUCTION

While intravenous (IV) medications are integral to the management of critically ill patients, the associated diluent volume contributes to the development of fluid overload (FO) and its sequalae, including mortality, increased intensive care unit (ICU) length of stay (LOS), increased acute kidney injury (AKI), and increased likelihood of mechanical intubation.^1–3^ Mitigating fluid overload with timely achievement of euvolemia is associated with improved outcomes.^4–7^ Given the complexity and prolific nature of mediation use in the ICU, data driven strategies are increasingly being employed to parse meaningful patterns for fluid overload prediction.^8–10^

While research is ongoing regarding identification of predictors for fluid overload, minimal research has evaluated the impact of medications as potential contributors.^11,12^ These studies have shown that medication regimen complexity, as measured by the medication regimen complexity-ICU (MRC-ICU), was related to fluid overload risk, using both traditional regression and supervised machine learning approaches.^8–10^ This score has also been shown to predict mortality^13^, LOS^14^, and prolonged duration of mechanical ventilation.^15–21^ Moreover, pharmacophenotyping based approaches including MRC-ICU and employing a common data model (CDM) for ICU medications (ICURx) have previously been created to allow for unsupervised cluster analysis machine learning that showed unique patterns of medication use and ICU complications, including FO.^22,23^ Therefore, quantifying patient-specific, medication-related data may be an important strategy in the prediction of fluid overload in critically adults.

No study has evaluated timing of medication administration in relation to fluid overload by reviewing the entire medication administration record (MAR) to identify patterns associated with medication administration. Unsupervised machine learning may be an optimal strategy for identifying factors associated with medication use and timing in relation to fluid overload. The purpose of this study was to employ unsupervised machine learning methods to uncover medication administration patterns that are correlated with the occurrence of FO. We hypothesized that unique clusters of medication use, particularly early in the ICU stay, would have a strong association with FO development.

## MATERIALS AND METHODS

This was a retrospective, observational study of adults admitted to critical care units at the University of North Carolina Health System (UNCHS) who had fluid overload data recorded. The protocol for this study was reviewed and approved by the UNHCS Institutional Review Board (approval number: (Study Number 20-2330); approval date: September 2020). Waivers of informed consent and HIPAA authorization were granted based on study design. All procedures were conducted with ethical standards of the UNHCS Institutional Review Board and the most recent version of the Helsinki Declaration of 1975.^24^ The reporting of this study adheres to the STrengthening and reporting of OBservational data in Epidemiology statement (STROBE).^25^

### Population

A trained Carolina Data Warehouse (CDW) analyst developed a random sample of 1,000 adult ICU patients (≥18 years) between October 2015 and October 2020 and extracted requested data from electronic health record (EHR) data (Epic Systems, Verona, WI). Patients were excluded if the data provided was not from their index ICU admission. These methods have been previously published.^9,10^

### Data Collection and Outcomes

The primary outcome was presence of fluid overload at 72 hours after initial ICU admission. Fluid overload was defined as a positive fluid balance (intake > output) in milliliters (mL) greater than or equal to 7% of the patient’s admission body weight in kilograms (kg).^1,3^ For example, a patient weighing 80kg at ICU admission with a positive fluid balance at 72 hours of 12,000 mL (or 12kg) would be classified as having fluid overload (positive fluid balance of 12kg is 15% of initial body weight).

Relevant patient demographics were extracted including: age, sex, race, ICU type, admission diagnosis, utilization of end-organ support including mechanical ventilation and renal replacement therapy, presence of AKI, use of vasopressors, Acute Physiology and Chronic Health Evaluation (APACHE) II score at 24 hours, and sequential organ failure assessment (SOFA) score at 24 hours. Additionally, patient outcomes, including in-hospital mortality, maximum fluid overload over 72 hours, and ICU LOS were collected.

The 72 hour study period was divided into 24 sets of three-hour intervals. Within this timeframe, the frequency of IV medication administration was calculated for each patient. All IV medications (any type of medication) as well as both oral and IV diuretics (e.g., furosemide, torsemide, chlorothiazide) were organized based on time of administration and separated into 3-hour groups (e.g., vancomycin given at hour 1 of ICU admission would be considered a different entity than vancomycin given at hour 12 of ICU admission). This allowed for both the medication and the timing of medication administration to be included in the unsupervised machine learning analysis. Medications were reviewed to combine any listed medications that were the same dose and volume (e.g., cefepime 1000mg/100 mL in normal saline (NS) mini-bag plus and cefepime 1000m/100mL NS infusion would be considered the same drug).

Antimicrobials that were infused over an extended interval (e.g., piperacillin-tazobactam) were not combined with the same antimicrobial infused over a standard duration. Additionally, the ICURx CDM was incorporated into analysis to provide additional information regarding specific ICU medications, including medication class and mode of administration (e.g. IV push versus continuous infusion).^17,26^

### Data Analysis

#### Unsupervised machine learning analysis

Principal Component Analysis (PCA) was performed at the patient level to create principal components (PCs) with a cumulative variance of over 85%.^27,28^ This dimension reduction approach was essential as it laid a robust foundation for the subsequent stages of our analysis, enabling us to effectively manage the complexity of the dataset.^27,28^ PCA helps remove redundant information and reduces the risk of overfitting, making the dimension-reduced representation more robust to noise and irrelevant features.^27,28^ Building upon the reduced dimensionality established by the PCA, we proceeded with the implementation of the Restricted Boltzmann Machine (RBM) which allowed us to identify the underlying structure in medication administration.^29,30^ By using the insights captured by the PCs, the RBM unveiled concealed layers.^29,30^ Following thorough hyperparameter tuning, which included adjustments to the number of neurons (hidden units), learning rate, and other factors, this process culminated in the successful classification of medications into distinct clusters.^31^ RBM can learn sparse representations of data, which means that only a subset of neurons (hidden units) is active at any given time.^29,30^ This sparsity can lead to more robust and interpretable representations, particularly in cases of the medication administration record where there are redundant or noisy features.^29,30^

The entire process is summarized in **Figure 1**.

**Figure 1.**
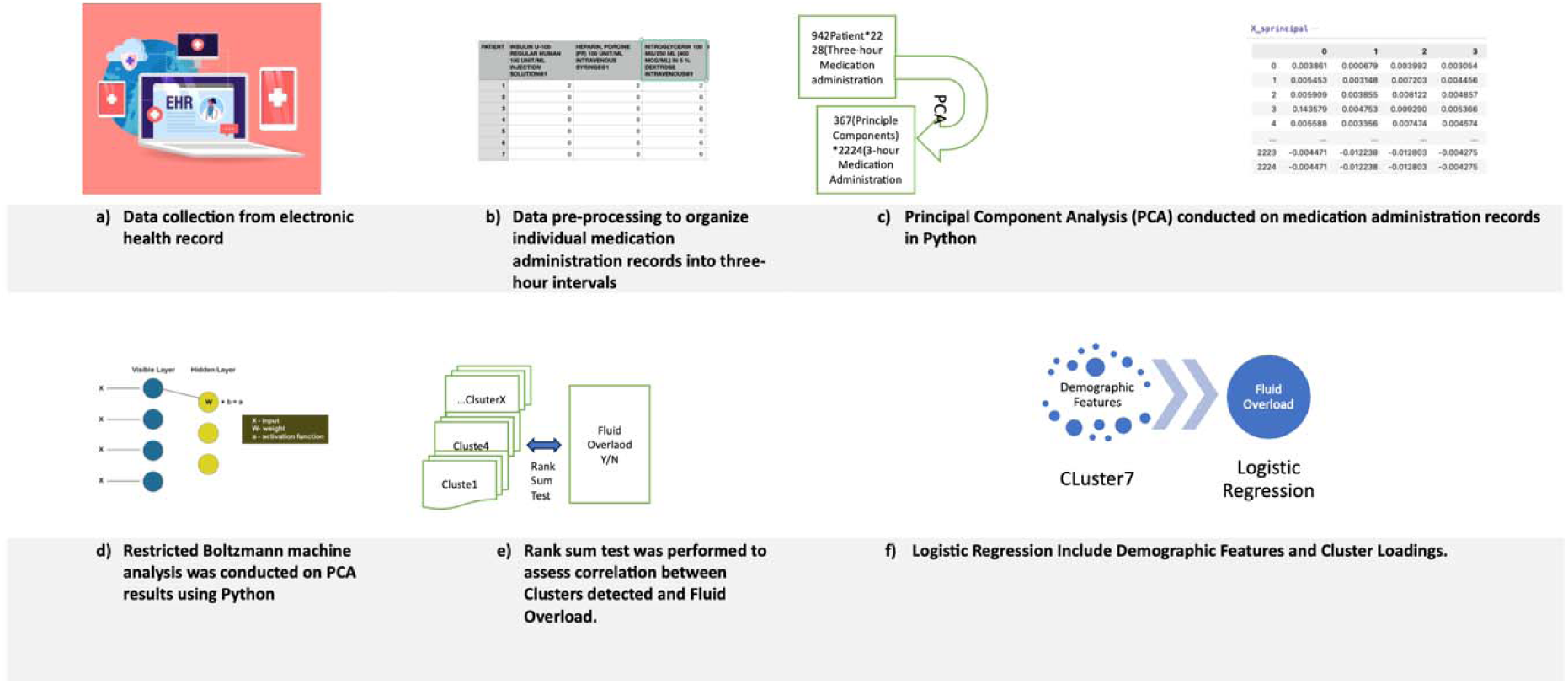
Workflow for unsupervised analysis of medications for prediction of fluid overload.

#### Test for the association between clusters and fluid overload

The rank sum test was employed to assess association between each cluster and the occurrence of fluid overload. Clusters demonstrating positive correlations were recognized through p-values lower than 0.05, accompanied by higher mean ranks among patients with fluid overload compared to those without fluid overload. Additionally, logistic regression analyses were performed to support the rank sum test, facilitating the identification and examination of the cluster with the highest association. For these ten logistic regression models (which corresponded to the ten medication clusters), the binary dependent outcome was fluid overload and each cluster’s standardized medication proportion was the independent variable. All analyses were performed in Python (version 3.0) and R (version 4.2.1).

#### Predictive modeling

Proportion of medications within the cluster most associated with FO was added to a logistic regression that included APACHE II score & diuretic use to determine if this feature would add to the ability of the model to predict fluid overload in individual patients. This was also done by time period (24, 48, 72 hours) to determine when the proportion of medications matching the cluster most associated with FO was most important in relation to development of fluid overload.

#### Descriptive characterization of clusters most associated with FO

Upon identification of medication clusters associated with FO, descriptive statistics were planned to explore and characterize these clusters. Analyses included categorization of medications in each cluster by medication class and analysis of frequency of medications occurring within each cluster (ex. Vancomycin appeared within the cluster X times). Additionally, the clusters were split into 24 hour periods to analyze which medications appeared in the cluster within specific ICU days (i.e. medications that appeared only within the first 24 hours of admission versus medications that appeared within the cluster multiple times throughout the 72 hour analysis period) to assess for a temporal relationship between medication administration and fluid overload. An exploratory analysis including variables of timing such as intermittent and bolus administration was also conducted.

## RESULTS

Among the 927 patients included in the study after removal of patients without fluid balance information (see Appendix for consort diagram), a total of 127 individuals (13.7%) experienced fluid overload. In the fluid overload cohort, the median fluid balance at 72 hours was 5934.17mL (3359.3-9156.4mL) vs. 300mL (IQR −894.1-1576.6 mL) in the non-fluid overload cohort. A total of 47,803 medication administrations occurred within the first 72 hours of ICU stay. Of these medication administrations, there were a total of 2,229 distinct combinations of medication plus timing of administration (ex. Cefazolin hours 0-3 of ICU stay, cefazolin hours 4-6 of ICU stay, etc.). Over the first 72 hours of ICU stay, all patients received a median of 31 distinct IV medication administrations (interquartile range: 13-65), with patients in the fluid overload group receiving a higher number of medication administrations compared to the non-fluid overload group (**Table 1**). Patients were mostly cared for in the medical ICU. Patients with fluid overload had a higher severity of illness as demonstrated by the APACHE II and SOFA scores at 24 hours, higher frequency of end-organ support including mechanical ventilation and renal replacement therapy, longer ICU LOS, and worse patient-centered outcomes including morbidity (e.g., AKI) and mortality. Patients in the fluid overload group received more medications overall as well as more vasopressors, sedatives, antibiotics, fluids, analgesics, gastric agents, anticonvulsants, and antidotes/rescue therapy compared to the non-FO group. **Table 1** provides a complete summary of demographics.

**Table 1.** Study population characteristics.

| Feature | All<br>(n=927) | Fluid overload<br>(n=127) | No fluid<br>overload (n=800) | p-value |
| --- | --- | --- | --- | --- |
| Age, mean (SD) | 60.9 (17.5) | 55.7 (18.3) | 61.7 (17.2) | 0.0008 |
| Female, n (%) | 398 (42.9) | 68 (53.5) | 330 (36.7) | 0.01 |
| Race, n (%) |  |  |  |  |
| Caucasian | 611 (65.9) | 72 (56.7) | 539 (59.9) | 0.04 |
| Black | 218 (23.5) | 35 (27.6) | 183 (20.3) |  |
| Other | 98 (10.6) | 20 (15.7) | 78 (8.7) |  |
| ICU type, n (%) |  |  |  |  |
| Medical | 368 (39.7) | 58 (45.7) | 310 (34.4) | <0.0001 |
| Cardiac | 286 (30.8) | 14 (11.0) | 272 (30.2) |  |
| Surgical | 97 (10.5) | 34 (26.8) | 63 (7) |  |
| Neurosciences | 91 (9.8) | 7 (5.5) | 84 (9.3) |  |
| Burn | 65 (7) | 11 (8.7) | 54 (6.0) |  |
| Other | 20 (2.2) | 3 (2.4) | 17 (1.9) |  |
| Admission diagnosis, n (%) |  |  |  |  |
| Cardiovascular | 228 (24.6) | 7 (5.5) | 221 (24.6) | <0.0001 |
| Neurology | 117 (12.6) | 9 (7.1) | 108 (12) |  |
| Pulmonary | 74 (8.0) | 11 (8.7) | 63 (7) |  |
| Trauma | 49 (5.3) | 15 (11.8) | 34 (3.8) |  |
| Infection including sepsis | 70 (7.6) | 10 (7.9) | 60 (6.7) |  |
| Gastrointestinal | 69 (7.4) | 18 (14.2) | 51 (5.7) |  |
| Neoplasm | 49 (5.3) | 6 (4.7) | 43 (4.8) |  |
| Dermatology | 15 (1.6) | 5 (3.9) | 10 (1.1) |  |
| Renal | 23 (2.5) | 3 (2.4) | 20 (2.2) |  |
| Endocrine | 21 (2.3) | 4 (3.1) | 17 (1.9) |  |
| Other | 212 (22.9) | 39 (30.7) | 273 (30.3) |  |
| Use of mechanical ventilation, n (%) | 305 (32.9) | 72 (56.7) | 233 (25.9) | <0.0001 |
| Renal Replacement Therapy, n (%) | 35 (3.8) | 16 (12.6) | 19 (2.4) | <0.0001 |
| Acute Kidney Injury, n (%) | 148 (16) | 45 (35.4) | 103 (12.9) | <0.0001 |
| Medication Administrations, mean (SD) | 51.2 (56.4) | 96.4 (81.3) | 44 (47.7) | <0.0001 |
| Medication Classes, mean (SD) |  |  |  |  |
| Analgesics | 7.9 (12.3) | 17.6 (18.6) | 6.4 (10.1) | <0.0001 |
| Antiarrhythmics | 0.6 (3.5) | 0.3 (1.7) | 0.6 (3.7) | 0.22 |
| Antibiotics | 3.2 (5.3) | 6.4 (7) | 2.7 (4.8) | <0.0001 |
| Anticoagulants | 3.1 (9) | 3.3 (9.7) | 3.1 (8.9) | 0.82 |
| Anticonvulsants | 0.1 (0.8) | 17.6 (1.7) | 6.4 (0.4) | <0.0001 |
| Antidotes/ Rescue Therapies | 0 (0.1) | 0 (0) | 0 (0.1) | 0.05 |
| Antifungals | 0 (0.3) | 0.1 (0.4) | 0 (0.3) | 0.50 |
| Antihypertensives | 1.3 (6.2) | 0.5 (5.3) | 1.4 (6.3) | 0.08 |
| Antiplatelets | 0.1 (0.8) | 0 (0) | 0.1 (0.9) | 0.06 |
| Diabetic Agents | 2.6 (4.9) | 2.7 (4.9) | 2.6 (5) | 0.84 |
| Diuretics | 0 (0.3) | 0 (0.1) | 0 (0.3) | 0.42 |
| Fluids | 11.2 (14.2) | 20.3 (17.2) | 9.7 (13.1) | <0.0001 |
| Gastric Agents | 1.6 (2.5) | 2.5 (2.6) | 1.5 (2.4) | <0.0001 |
| Hypertonic Saline | 0.2 (2.4) | 0.3 (2.6) | 0.2 (2.4) | 0.87 |
| Immunosuppressants | 0 (0.1) | 0 (0.4) | 0 (0) | 0.32 |
| Inotropic Agents | 0.7 (4.4) | 0.3 (2.2) | 0.8 (4.7) | 0.05 |
| Neuromuscular Blocking Agents) | 0.1 (0.5) | 0.1 (0.3) | 0.1 (0.5) | 0.49 |
| Sedatives | 9.4 (17.7) | 19.2 (27.3) | 7.8 (15) | <0.0001 |
| Somatostatic Agents | 0 (0.1) | 0 (0.3) | 0 (0.2) | 0.27 |
| Total parenteral nutrition | 0.1 (1.1) | 0.4 (2.7) | 0 (0.4) | 0.16 |
| Vasopressors | 8.9 (23.3) | 22.1 (40.6) | 6.8 (18.4) | <0.0001 |
| **Severity scores at 24 hours of ICU admission** |  |  |  |  |
| **APACHE II**, mean (SD) | 14.5 (6.4) | 17.4 (6.9) | 14 (6.2) | <0.0001 |
| **SOFA**, mean (SD) | 8.1 (3.3) | 10.5 (3.4) | 7.8 (3.2) | <0.0001 |
| **Patient outcomes** |  |  |  |  |
| **In-hospital Mortality**, n (%) | 88 (9.5) | 23 (18.1) | 65 (8.1) | 0.0007 |
| **Maximum Fluid Overload at 72 h**, mL, mean (SD) | 2364.9 (3995.5) | 9519.9 (5629.2) | 1229 (2018.7) | <0.0001 |
| **ICU length of stay**, days, mean (SD) | 2.9 (9.8) | 9.7 (13.9) | 4.5 (8.7) | <0.0001 |
| Data are presented as n (%) or mean (standard deviation (SD)) unless otherwise stated.<br>SOFA sequential organ failure assessment, APACHE II Acute Physiology and Chronic Health Evaluation; ICU intensive care unit. P-values from t.test (variables with mean and SD) and chi-square test (variables with count and %). |  |  |  |  |

The PCA was conducted to identify clusters of medications. While early models of the PCA included patient-specific information including SOFA score, age, sex, etc., these factors were not significant in identifying the clusters and were excluded from the final model, which included only medications and timing of administration. The proposed unsupervised machine learning modeling yielded 10 distinct clusters (**Figure 1**). There were a median of 532 (interquartile range (IQR) 520-539.8) medications in each cluster when medications were associated with an administration time (ex. vancomycin 1g at hour 3 is considered a separate medication than vancomycin 1g at hour 8), and 121-130 medications in each cluster when administration time was not considered. 97 medications were identical in every cluster when timing was not considered, but when timing of medication administration was factored in, the clusters were significantly different with no medication plus timing combinations being identical in all clusters. Figures 2 & 3 show overlap between clusters when categorizing medications within each cluster by medication administrations (Figure 2) and medication names (Figure 3). The medications appearing in each cluster are listed in the **Digital Supplementary Materials**. Additionally, medications were categorized by class, and these proportions are reported in **Table 2**.

**Figure 2.**
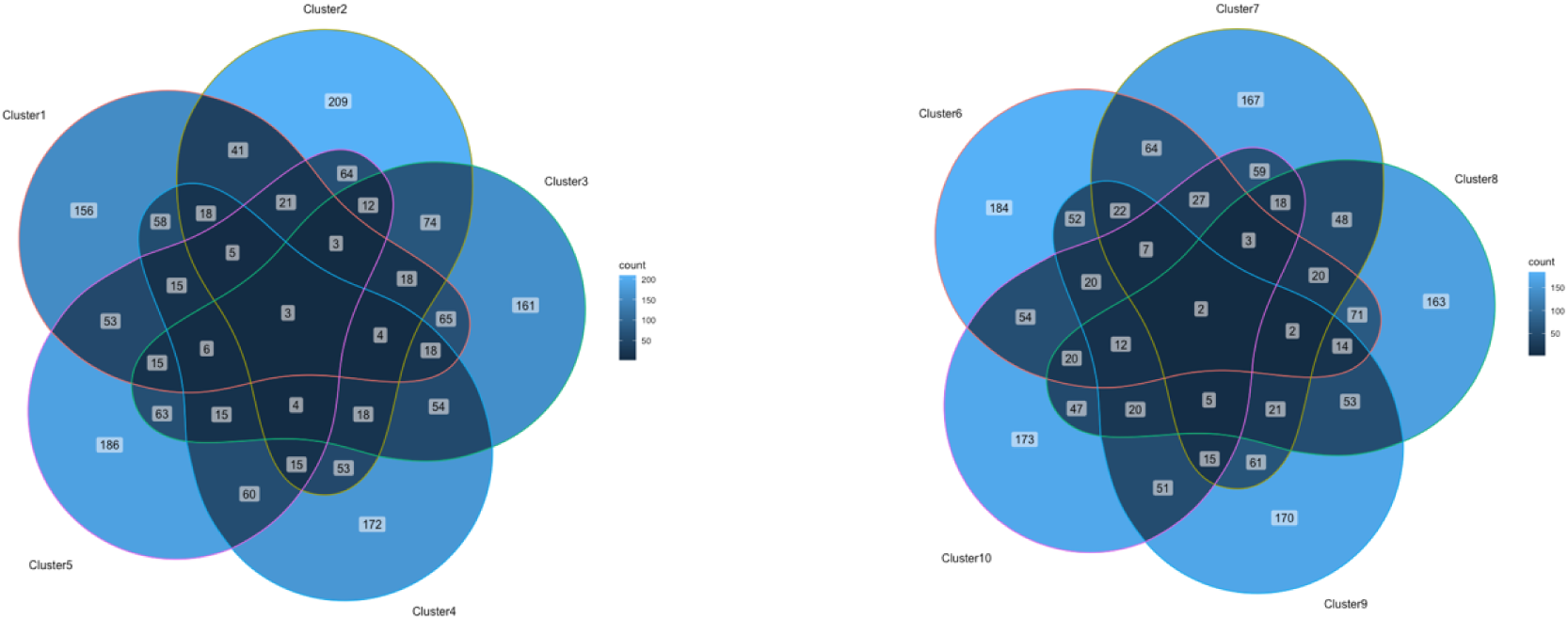
Venn diagrams of medication overlap within the 10 clusters by medication name & timing Venn Diagrams Illustrating Medication Overlaps Between Clusters 1-5 and Clusters 6-10, with Numerical Values Indicating the Count of Shared *Medication Administrations* (Both medication name and time period of administration)

**Figure 3.**
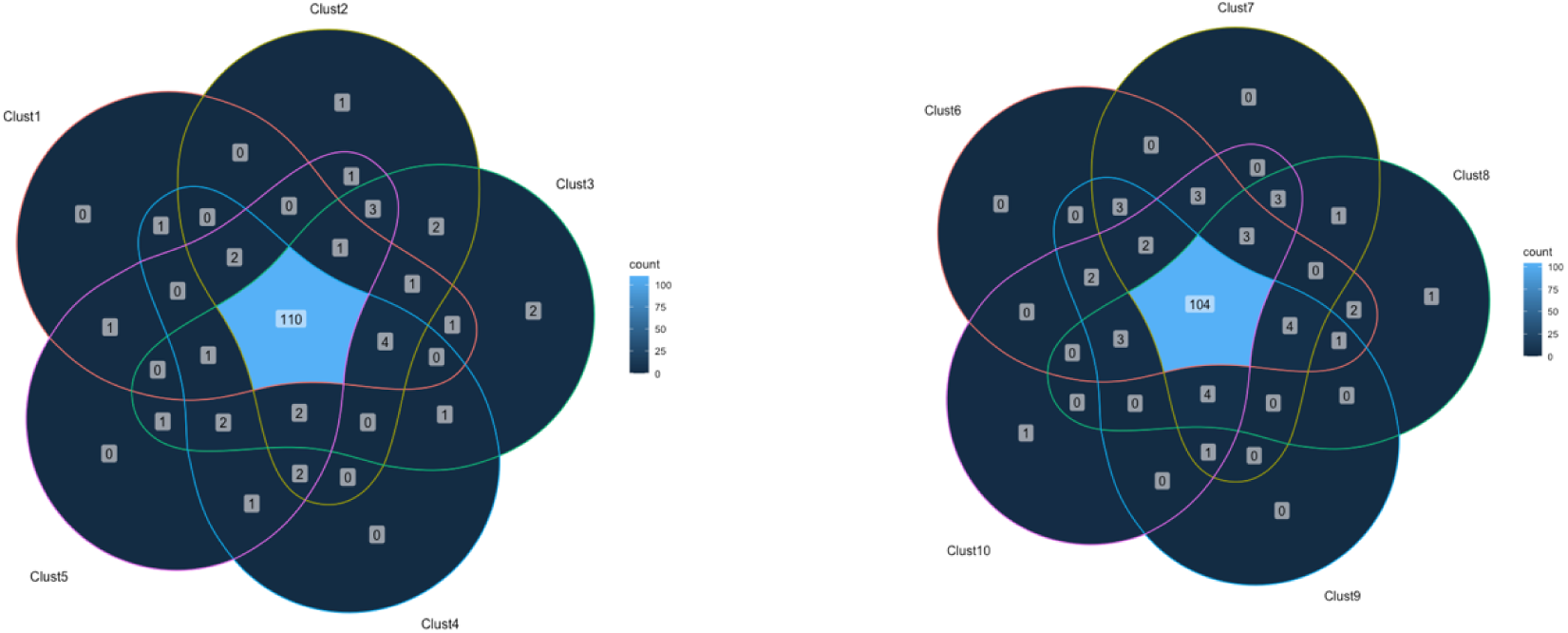
Venn diagrams of medication overlap within the 10 clusters by medication name only Venn Diagrams Illustrating Medication Overlaps Between Clusters 1-5 and Clusters 6-10, with Numerical Values Indicating the Count of Shared *Medication Names*

**Table 2.**
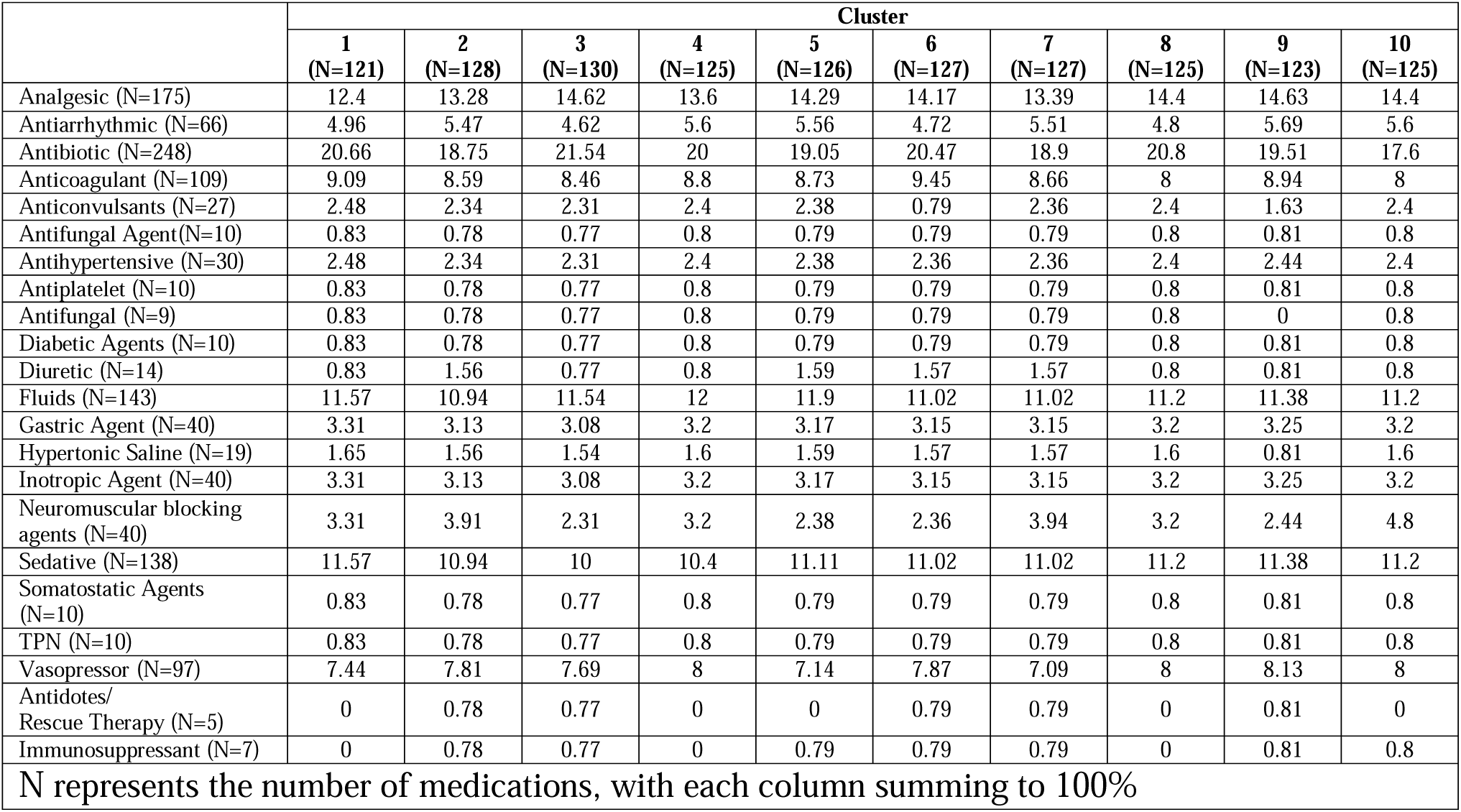
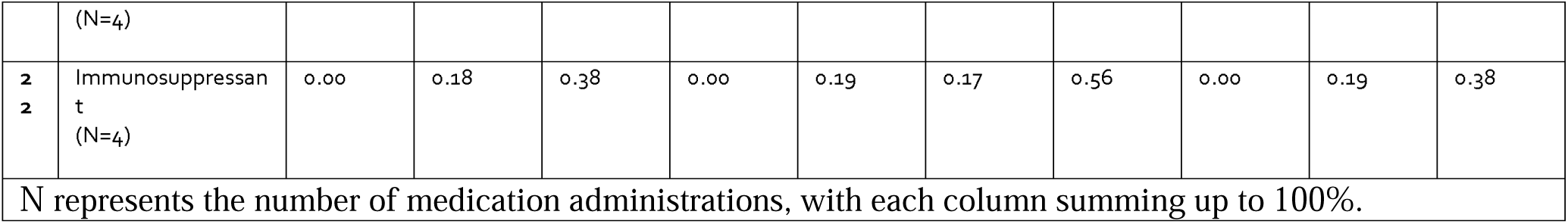
Distribution of medication classes within each medication cluster.

Clusters 5 and 7 had a positive association with fluid overload based on the rank sum test (**Table 3**). Patients who experienced fluid overload received a higher mean number of Cluster 5 (18.7 medications vs. 7.7 medications, p < 0.0001) and Cluster 7 medications (25.6 medications vs. 10.9 medications, p < 0.0001) in comparison to patients who did not experience fluid overload. Ten logistic regression models were employed, yielding similar results to those of the rank sum test (**Table 4**). While Cluster 5 also was associated with fluid overload, its association was weaker than that of Cluster 7. Analyses of Cluster 5 can be found in the **Supplementary Appendix**.

**Table 3.**
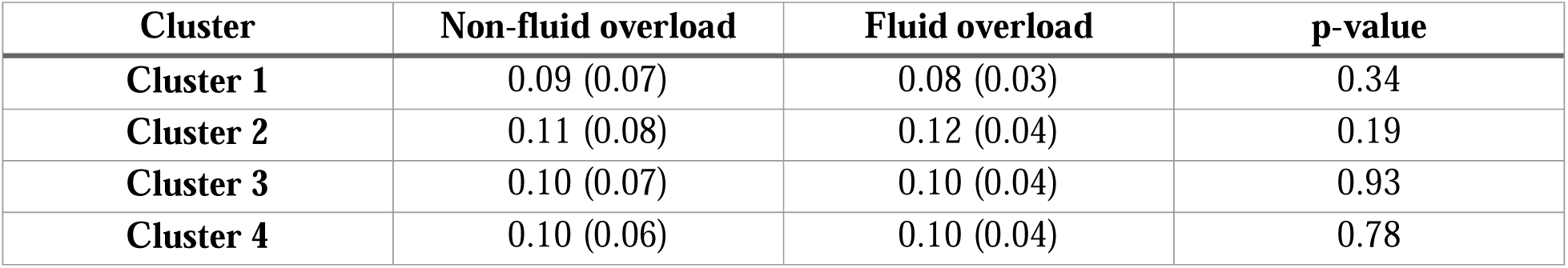
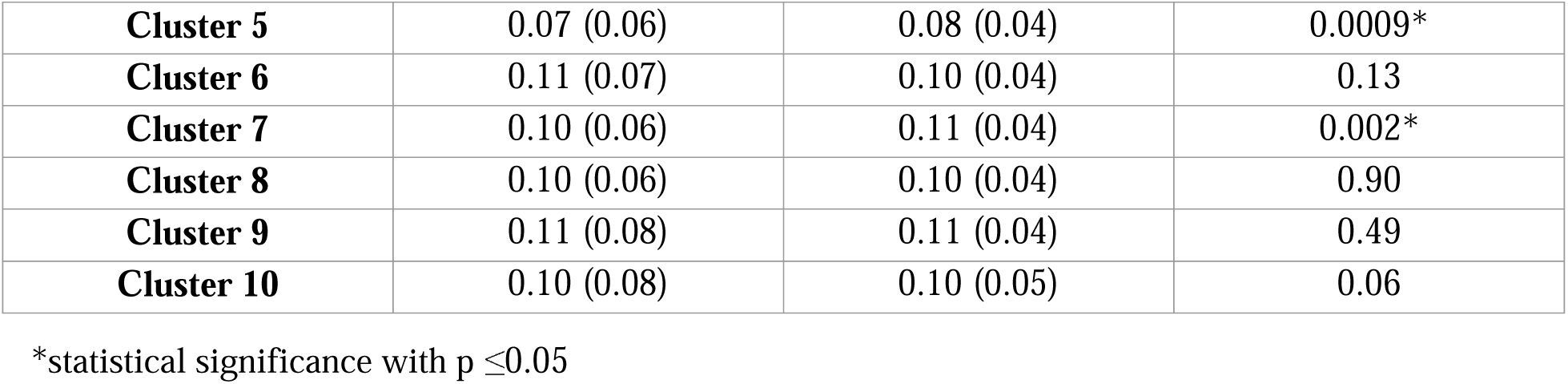
Rank Sum Test for Each Medication Cluster. Rank Sum Test Results for Medication Clusters (Standardized Proportions across Patients) and Fluid Overload

| Cluster | Non-fluid overload | Fluid overload | p-value |
| --- | --- | --- | --- |
| **Cluster 1** | 0.09 (0.07) | 0.08 (0.03) | 0.34 |
| **Cluster 2** | 0.11 (0.08) | 0.12 (0.04) | 0.19 |
| **Cluster 3** | 0.10 (0.07) | 0.10 (0.04) | 0.93 |
| **Cluster 4** | 0.10 (0.06) | 0.10 (0.04) | 0.78 |
| **Cluster 5** | 0.07 (0.06) | 0.08 (0.04) | 0.0009* |
| **Cluster 6** | 0.11 (0.07) | 0.10 (0.04) | 0.13 |
| **Cluster 7** | 0.10 (0.06) | 0.11 (0.04) | 0.002* |
| **Cluster 8** | 0.10 (0.06) | 0.10 (0.04) | 0.90 |
| **Cluster 9** | 0.11 (0.08) | 0.11 (0.04) | 0.49 |
| **Cluster 10** | 0.10 (0.08) | 0.10 (0.05) | 0.06 |

\*statistical significance with $p \leq 0.05$

**Table 4.** Logistic Regression for Each Cluster. Results of 10 Fitted Logistic Regression Models: Dependent Variable - Fluid Overload, Independent Variable - Medication Clusters (Standardized Proportions across Patients).

| **10 Logistic Regression Models** | **Estimated Value** | **P Value** |
| --- | --- | --- |
| **Cluster 1** | -2.10 | 0.22 |
| **Cluster 2** | 0.42 | 0.73 |
| **Cluster 3** | -1.41 | 0.38 |
| **Cluster 4** | 0.16 | 0.92 |
| **Cluster 5** | 2.68 | 0.07 |
| **Cluster 6** | -2.97 | 0.08 |
| **Cluster 7** | 2.96 | 0.05* |
| **Cluster 8** | -0.87 | 0.58 |
| **Cluster 9** | -0.12 | 0.93 |
| **Cluster 10** | 0.56 | 0.64 |

\*statistical significance with $p \leq 0.05$

Notably, Cluster 7, which consisted of 127 unique IV medications, exhibited the highest estimated value and the smallest p-value, signifying its substantial contribution to the development of fluid overload. The medications found within Cluster 7 were diverse, with high representation among continuous infusions, antibiotics, as well as sedatives and analgesics (**Table 2**). A total of 51 medications (40%) were administered in >5 separate 3-hour intervals, and fifteen medications within Cluster 7 were administered exclusively within the initial 24 hours of ICU admission (**Table 5**). Patients with fluid overload were more likely to have medications appear within Cluster 7 than patients without fluid overload (**Table 6). Table 7** provides a list of medications appearing on each of the first 3 days of ICU admission within Cluster 7. Figure 4 shows all of the medications within Cluster 7 and how frequently they appeared based on timing of administration. Figure 5 reveals medications and timing of medication administrations within Cluster 7 ordered from most frequent to least frequent appearance within the Cluster. **Table 6** splits the medications from Cluster 7 into each day of ICU stay (first 72 hours), and Figure 6 includes timing of medication administrations separated by medication class. Figure 7 represents the frequency at which each 3-hour time slot appeared within Cluster 7 (ex. 27 medications given within 0-3 hours of ICU admission appeared within Cluster 7 compared to only 17 medications within the 69-72 hour time slot).

**Figure 4.**
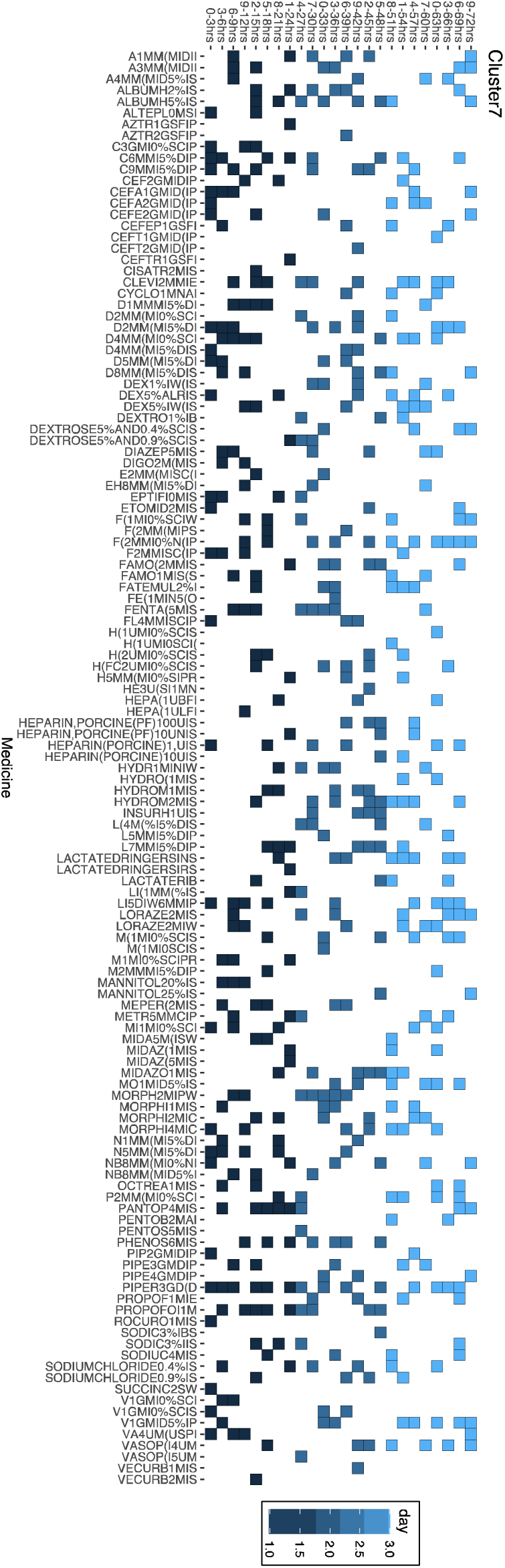
Cluster 7 medications organized by timing of medication administrations. Medication Record of Cluster 7 Distribution Over 72 Hours, with boxes indicating administration of medication at specific time slot

**Figure 5.**
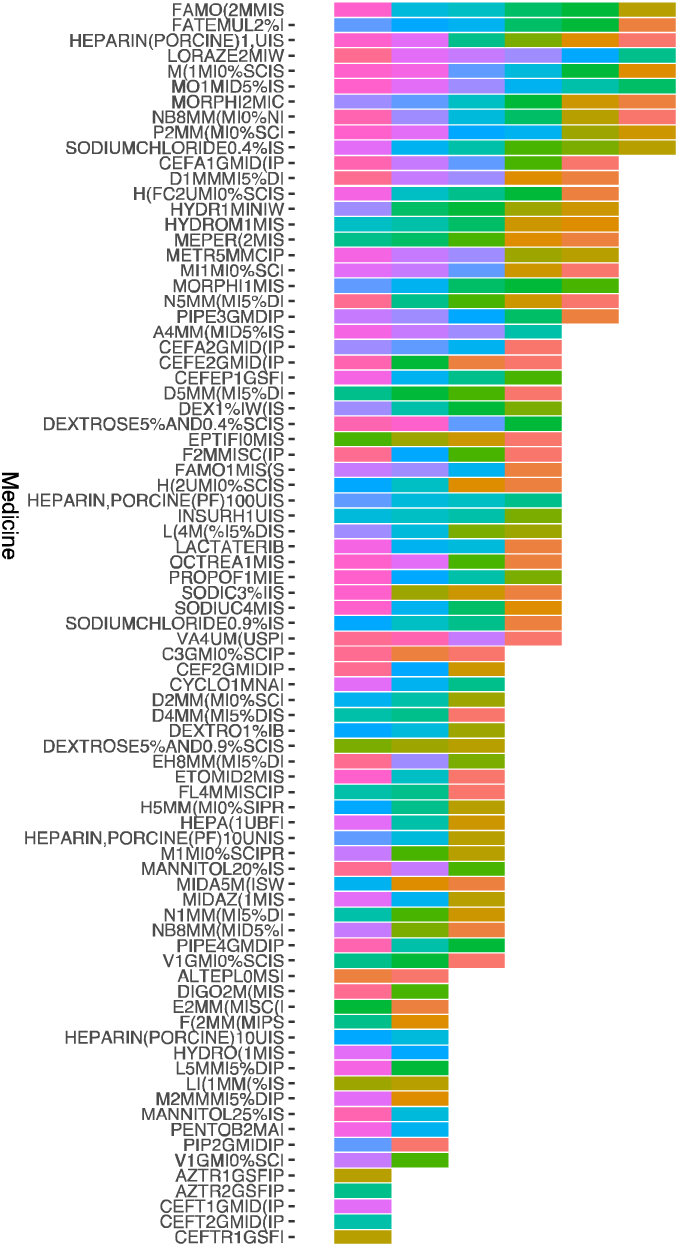
Cluster 7 medications organized by frequency and timing of administration. Distribution of Medication Records for Cluster 7 Over 72 Hours. Horizontal Axis: Medication Names. Vertical Axis: Frequency of Appearance in Time Slots

**Figure 6.**
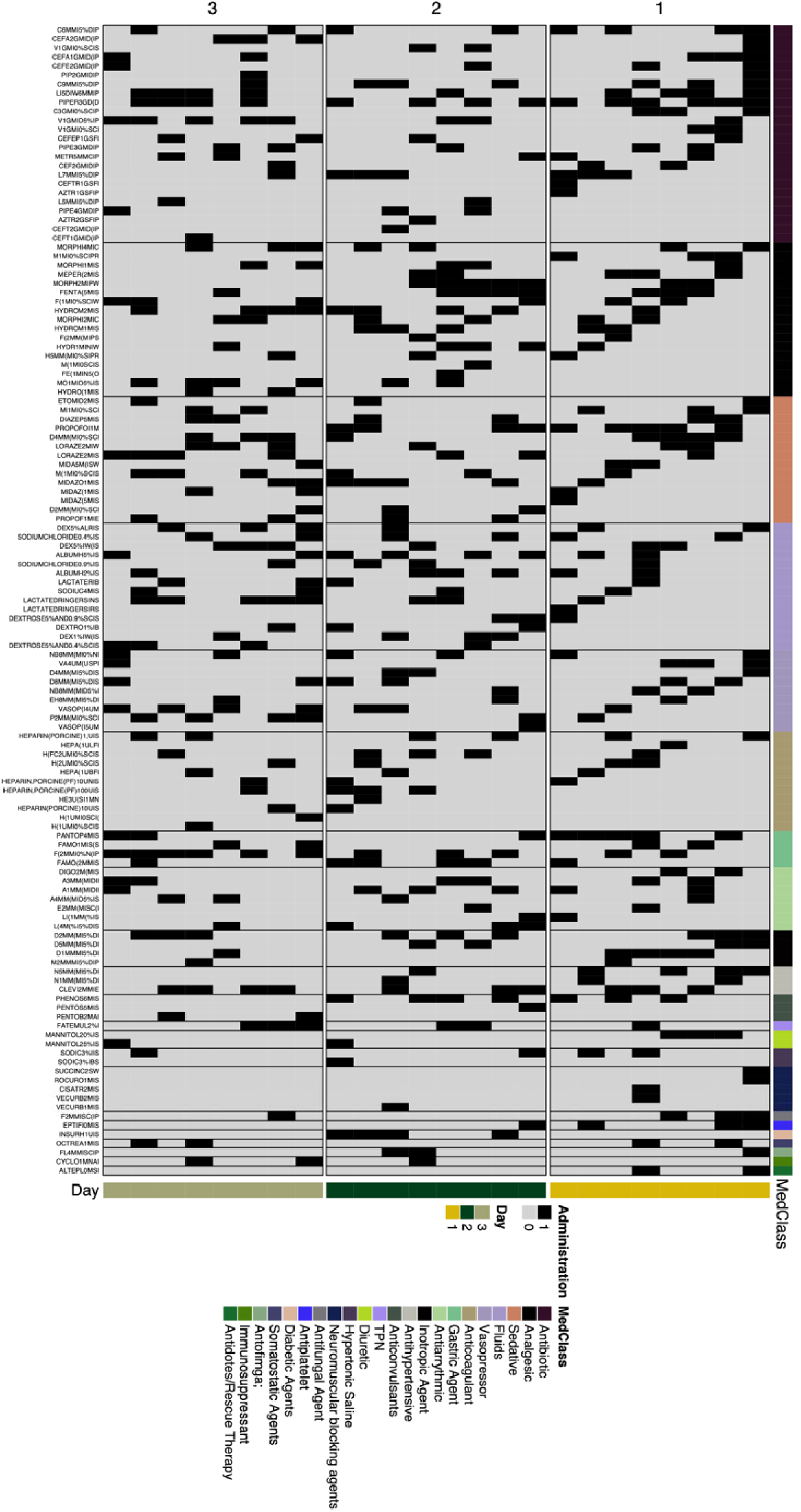
Cluster 7 medications organized by timing of administration and medication class.

**Figure 7.**
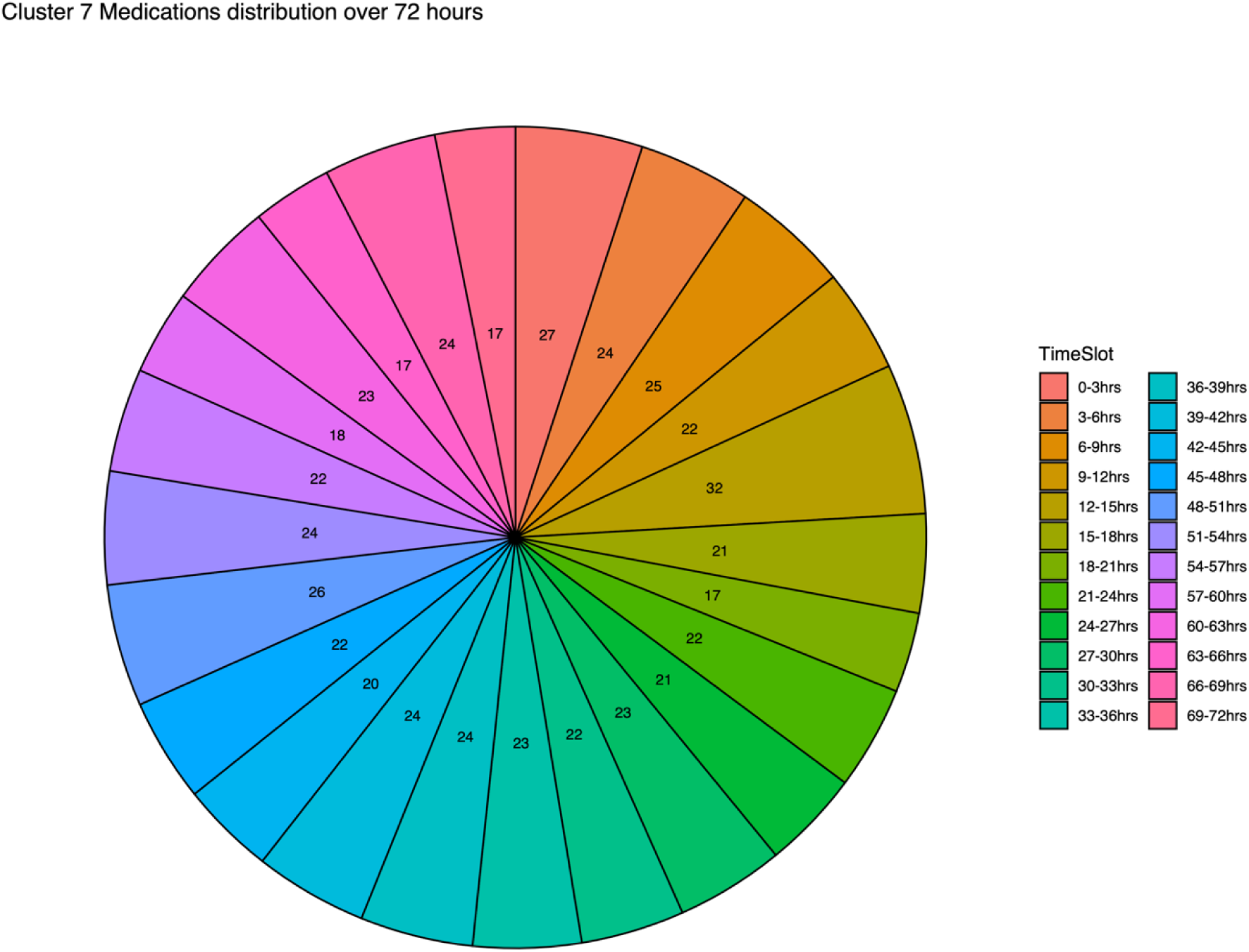
Cluster 7 medications organized by proportion of medications from each 3-hour time period The Custer 7 medication administration is distributed over a span of 72 hours, divided into twenty-four three-hour time slots. These slots are arranged clockwise, starting from the 0-3 hour slot and ending at the 68-72 hour slot. The term "area" represents the quantity of medication detected within each respective time slot.

**Table 5.**
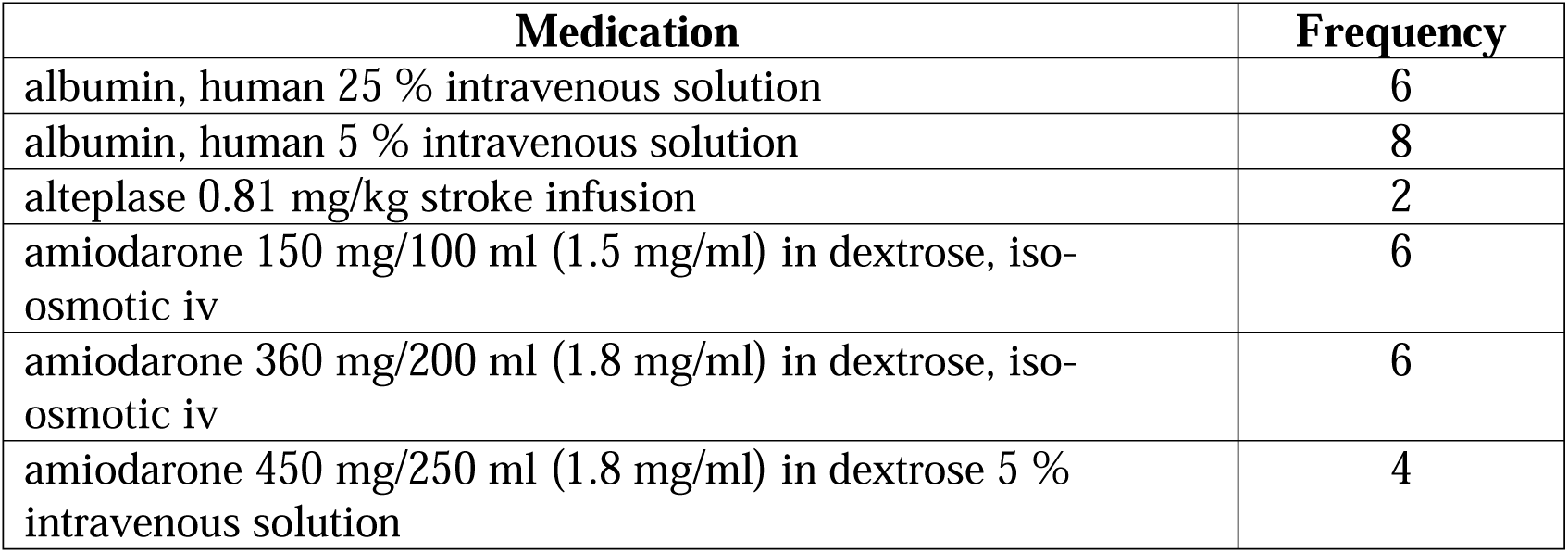
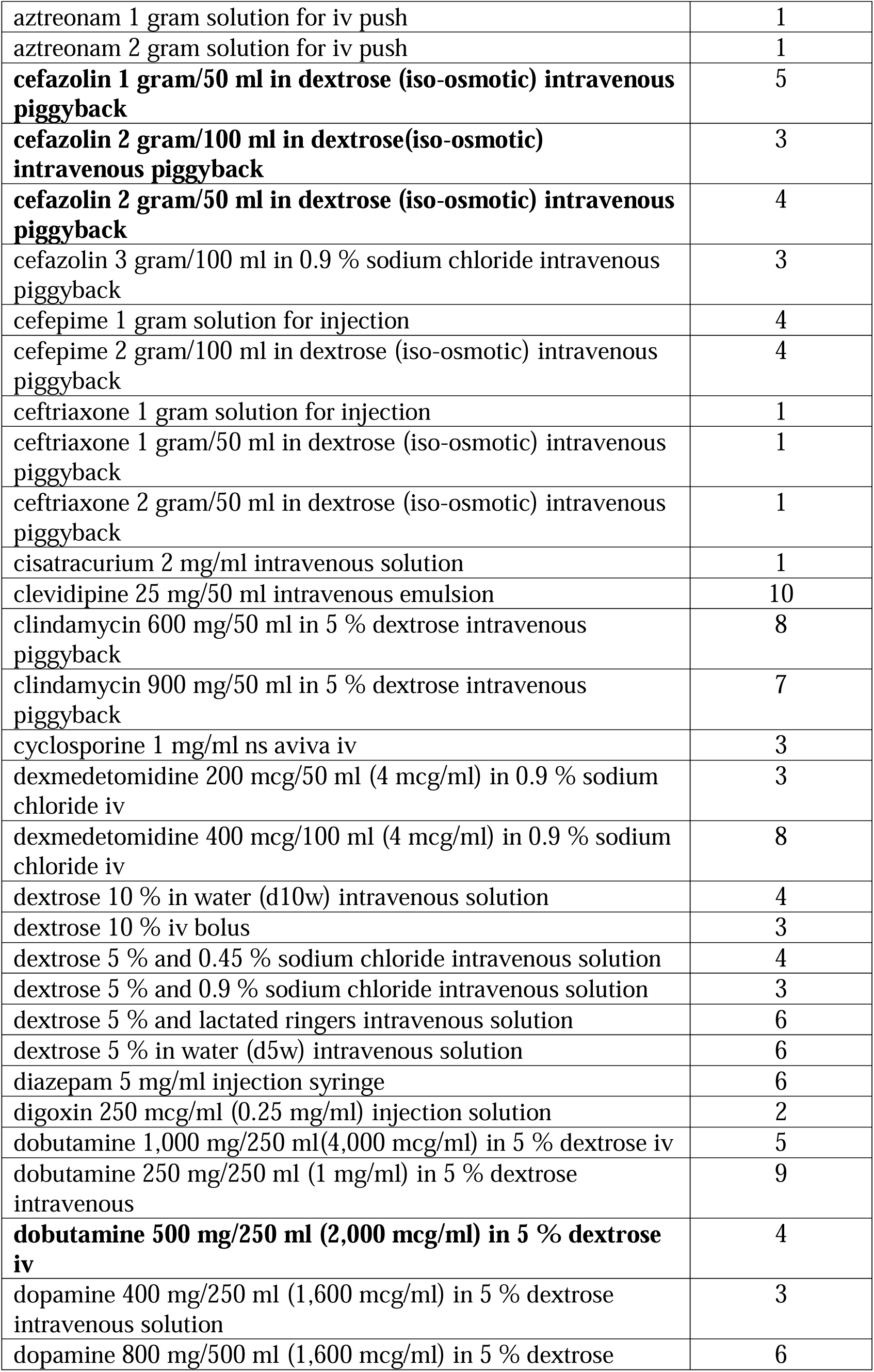
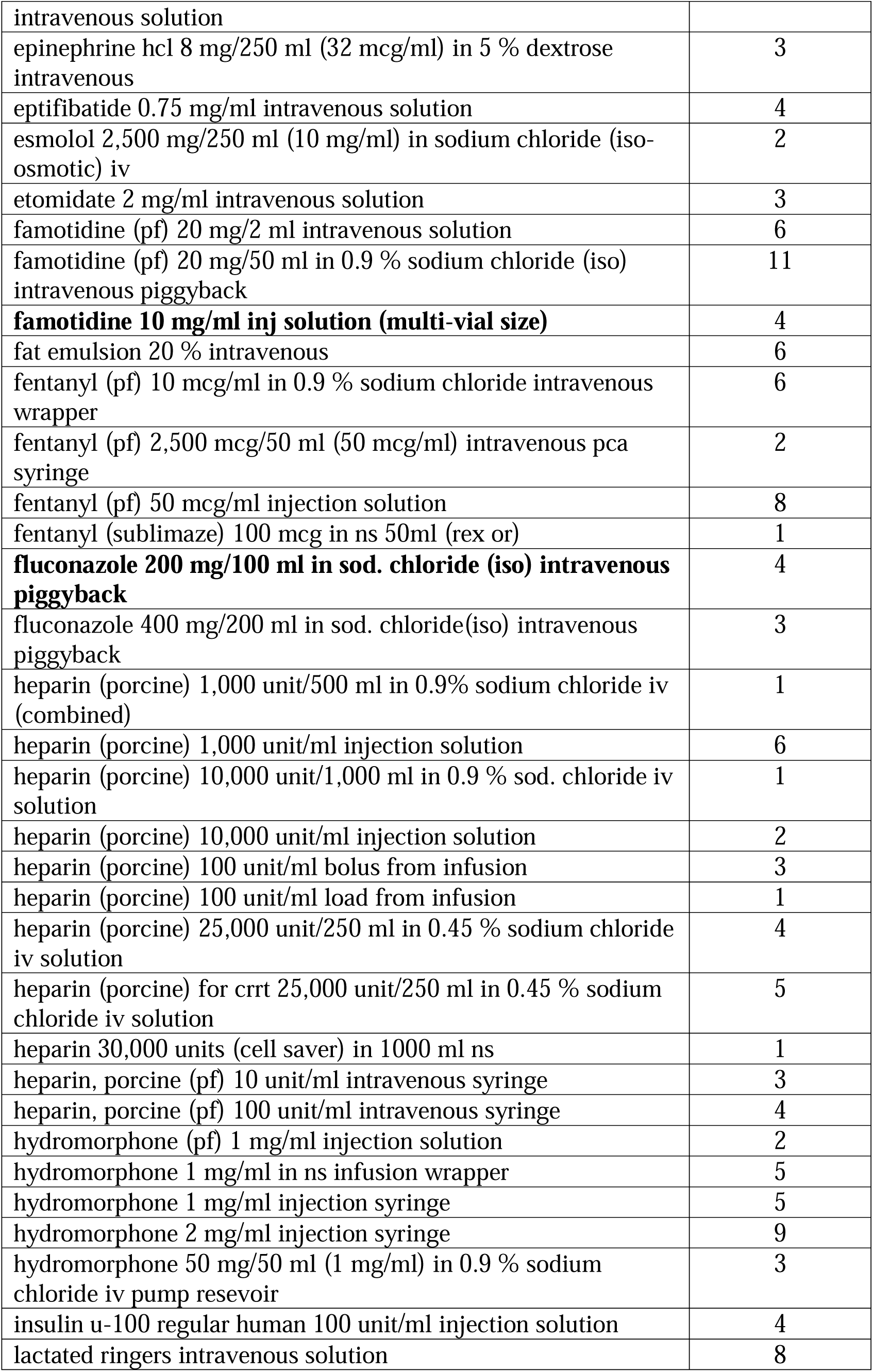
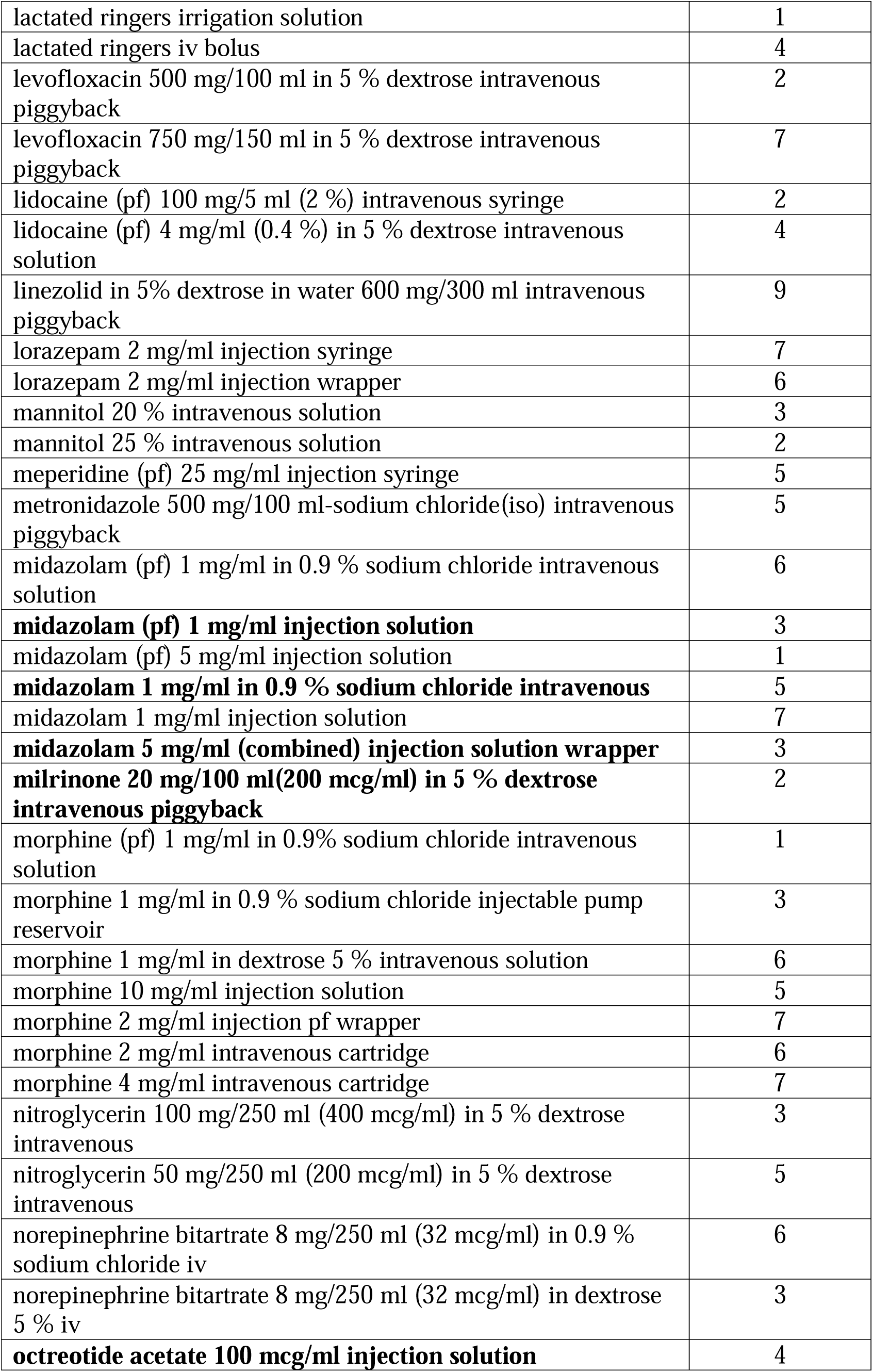
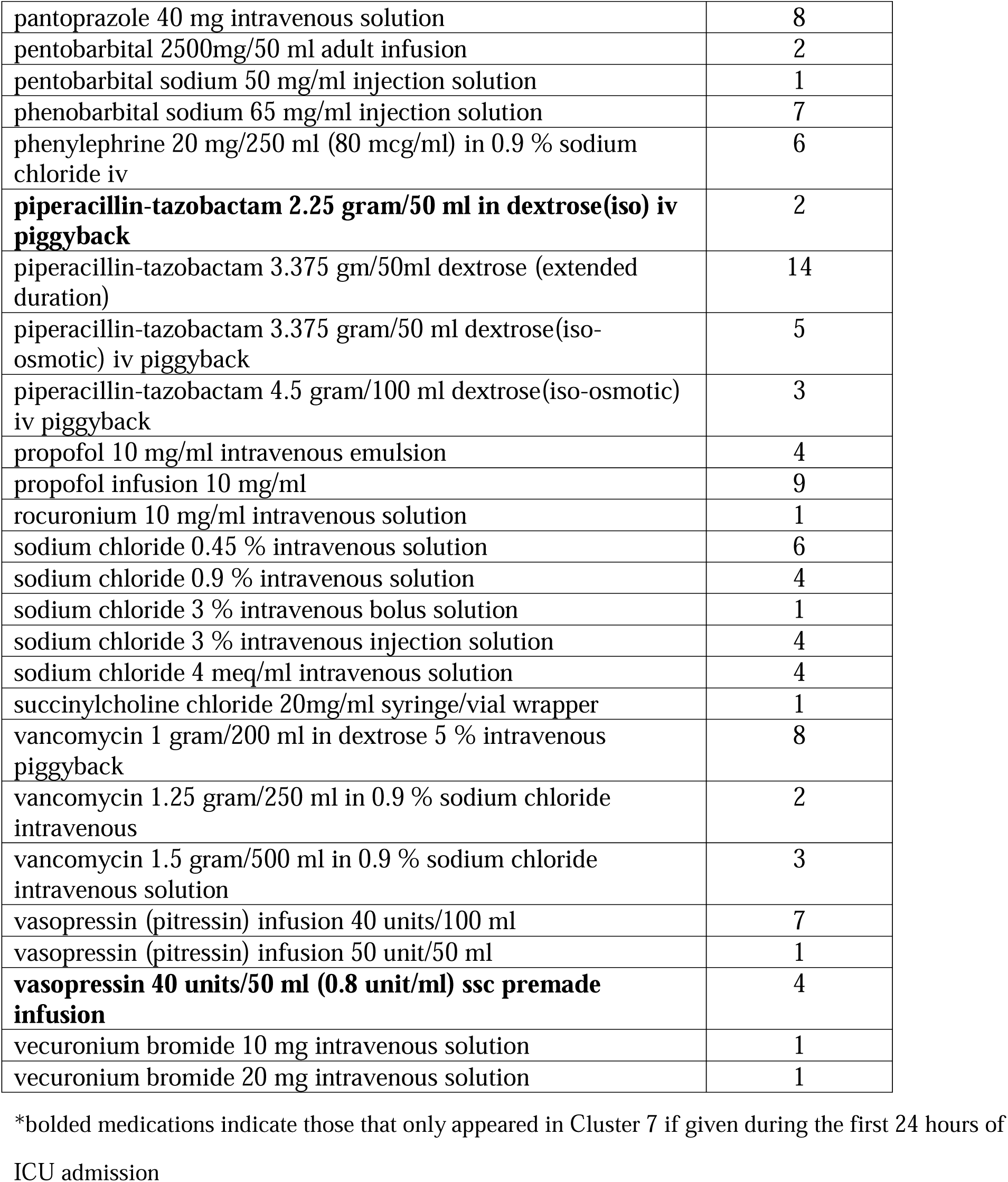
Frequency of Medications Appearing in Cluster 7.

**Table 6.** Proportion of Medications Appearing in Cluster 7 by Day and Fluid Overload Status.

| Proportion of Medications Matching Cluster 7, median | Fluid Overload Group | Non-Fluid Overload Group |
| --- | --- | --- |

| (IQR) |  |  |
| --- | --- | --- |
| Hours 0-24 of ICU admission | 52.4 (33.9-70.4) | 33.3 (14.3-51.2) |
| Hours 25-48 of ICU admission | 50 (22.2-65.4) | 18.8 (5.6-40.7) |
| Hours 49-72 of ICU admission | 32.4 (11.8-50) | 7.7 (0-22.5) |
| Hours 0-72 of ICU admission | 46.3 (30.4,60.7) | 25.2 (11.8-40) |

Proportions are calculated on a patient-specific level; the percentage is reported is the median proportion of medications matching Cluster 7 using individual patient data rather than aggregate data. For example, of the medications received in the first 24 hours of ICU stay, 52.4% of those medications were also present in Cluster 7 for the median patient in the fluid overload group, compared to 33.3% of medications in the non-fluid overload group.

**Table 7.**
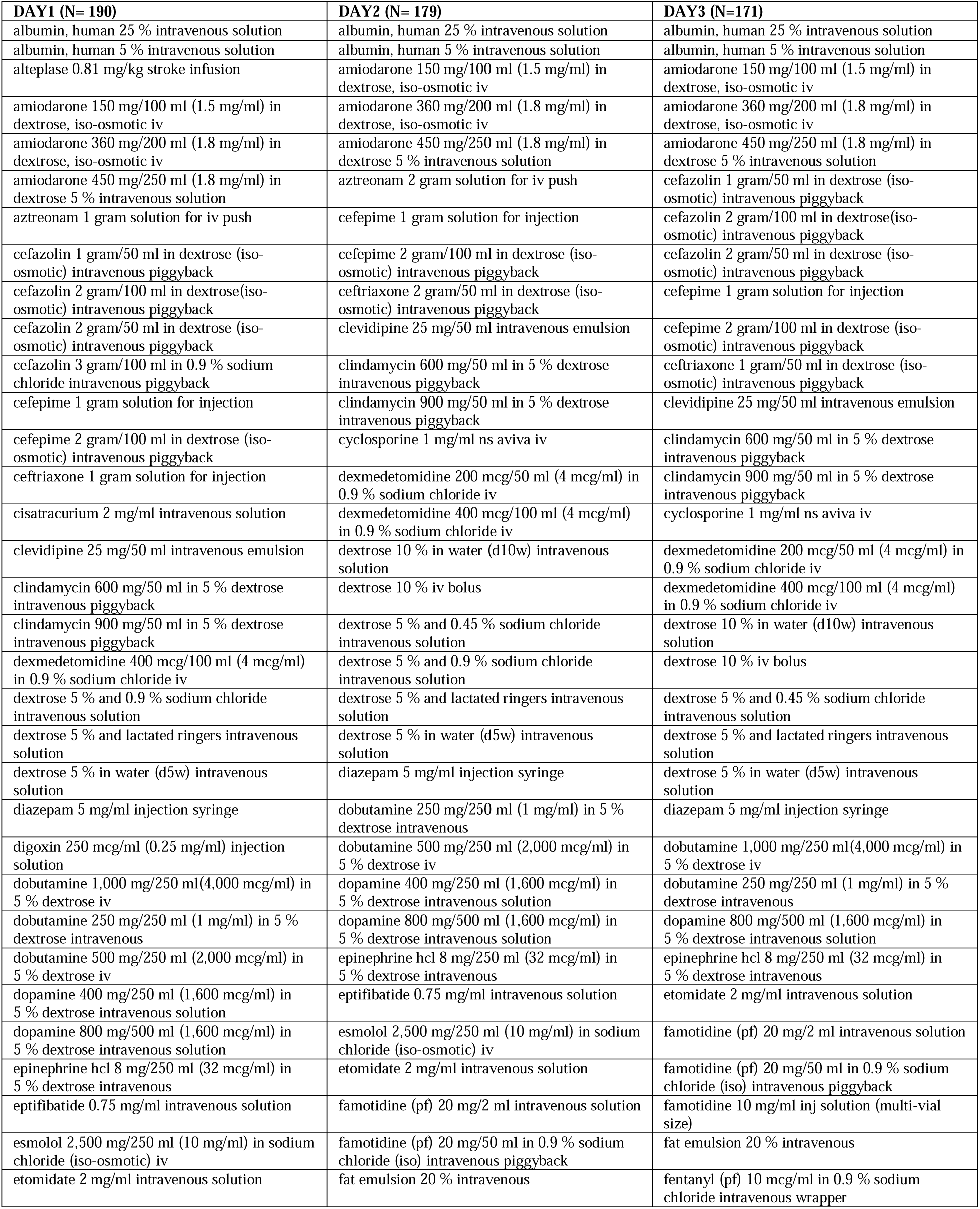
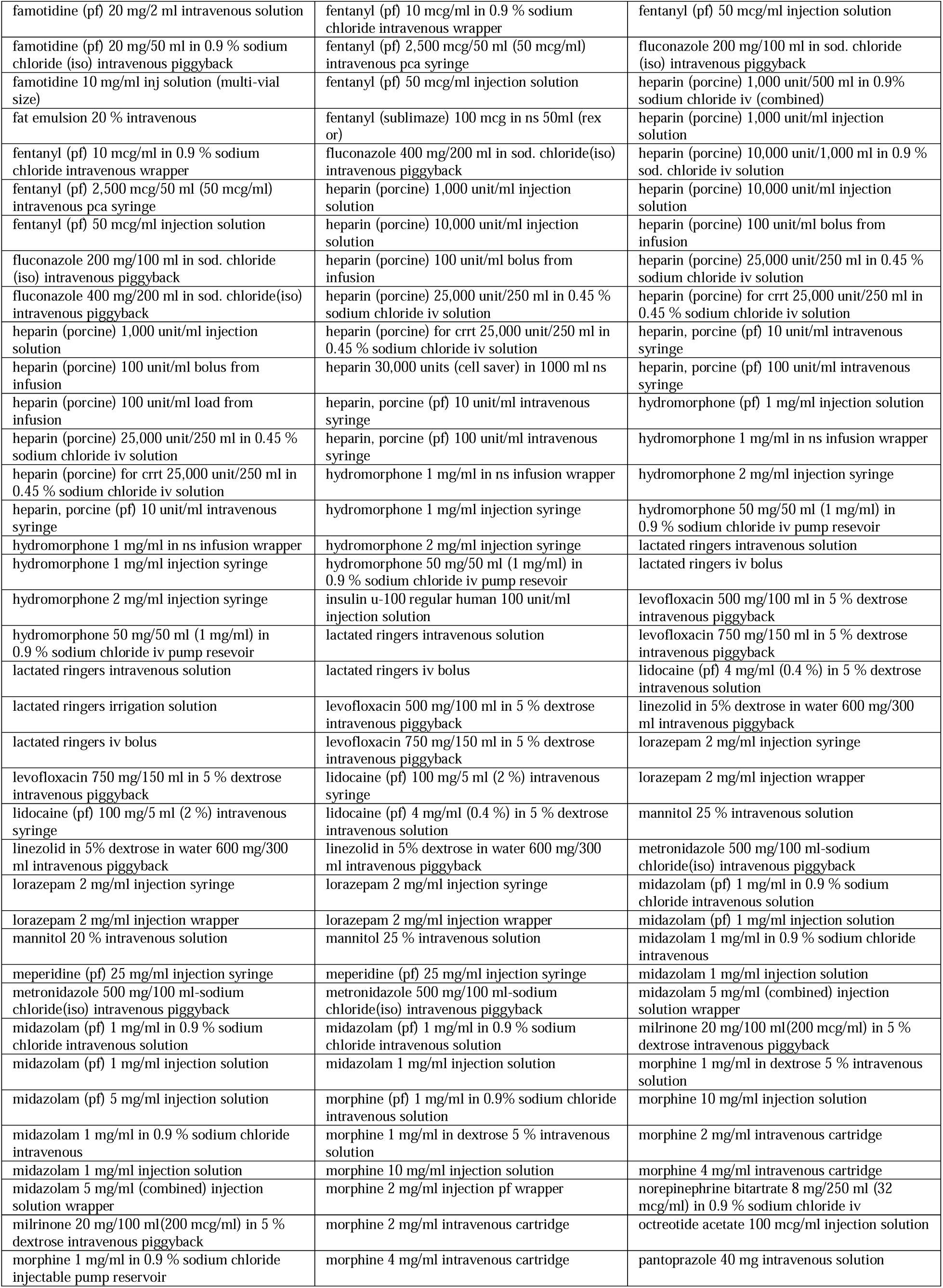
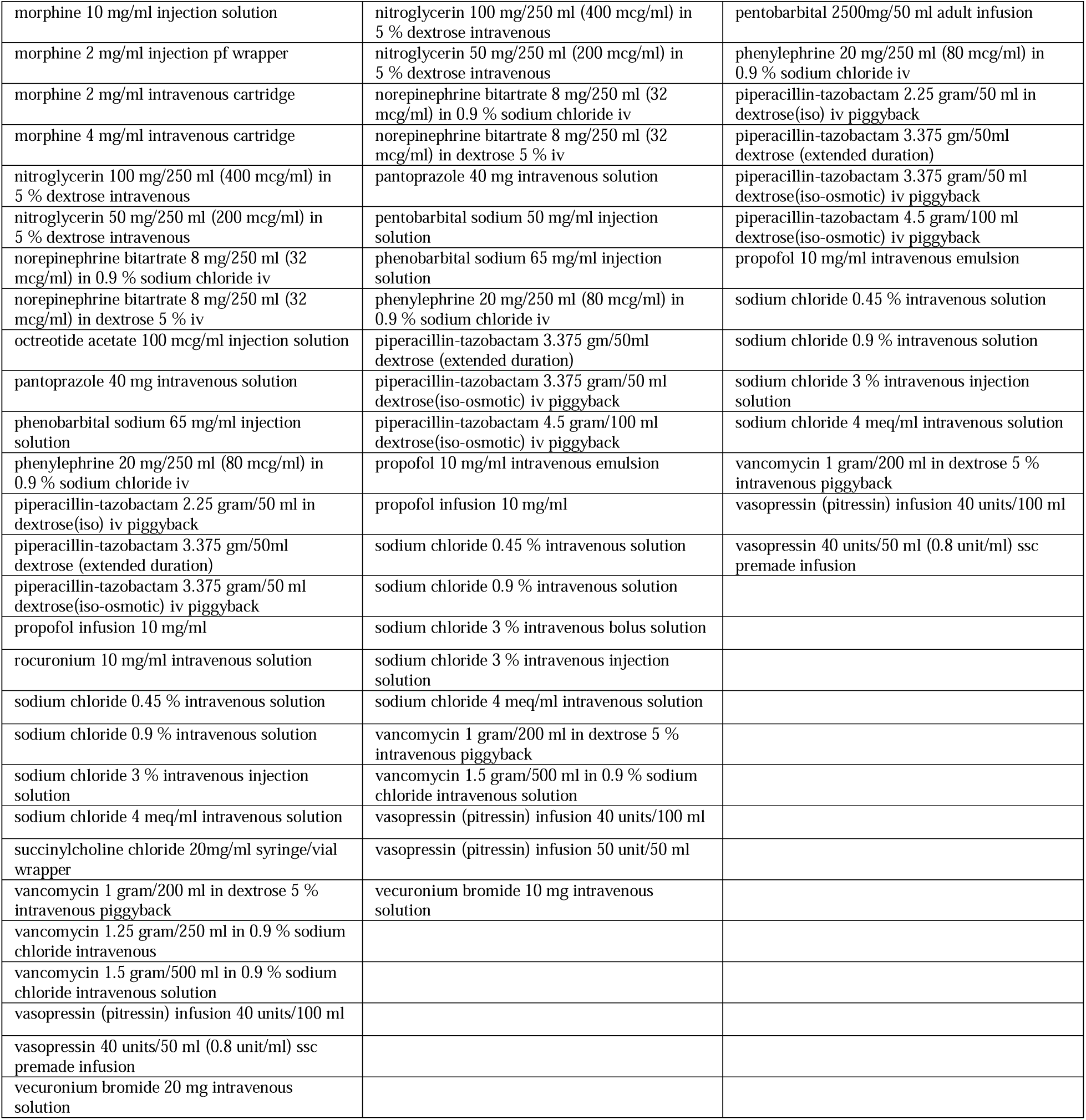
Cluster 7 medications by ICU day. Distribution of Medications in Cluster 7 Across ICU Days (Day 1: 0-24 hours, Day 2: 24-48 hours, Day 3: 48-72 hours), N is total number of medications that appeared within that day

Additionally, Cluster 7 improved predictive models for fluid overload. A logistic regression model including Cluster 7, APACHE score at 24 hours of ICU admission, and levels of diuretic administration demonstrated an improvement in the model (**Table 8**). This was evidenced by a reduction in the AIC from 673.6 to 663.43, with a notably significant estimated p-value of <0.0005. Additionally, integrating this feature in the model led to an enhanced ROC curve, elevating the AUC from 0.7193 to 0.7413 (Figure 8). An additional visualization of the impact of Cluster 7 on predictive modeling can be seen in Figure 9. Additionally, when dividing Cluster 7 into proportion of medications within Cluster 7 given at each day of ICU stay, a higher proportion of Cluster 7 medications on Days 1 and 3 of ICU admission was associated with increased risk of fluid overload (**Table 9**). Figure 10 shows the distribution of patients in each group (fluid overload and non-fluid overload) based on proportion of their medications that matched Cluster 7. Figure 11 is an example of the marginal effect of proportion of medications matching Cluster 7 and association with fluid overload when normalized to APACHE II score of 14 and receipt of no diuretics.

**Figure 8.**
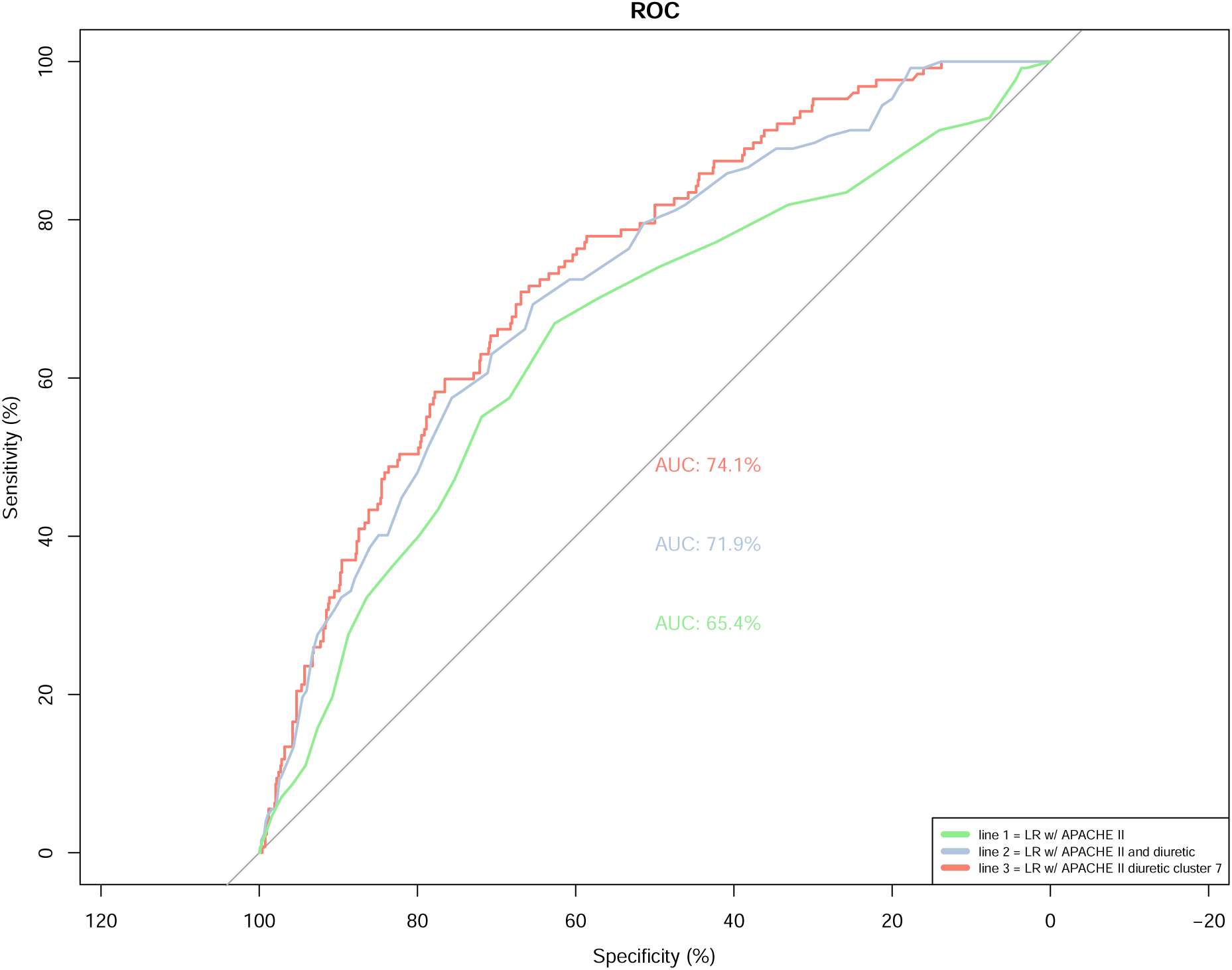
Logistic regression model for Cluster 7. Logistic regression for incidence of fluid overload, including Cluster 7, APACHE II score, and diuretic level

**Figure 9.**
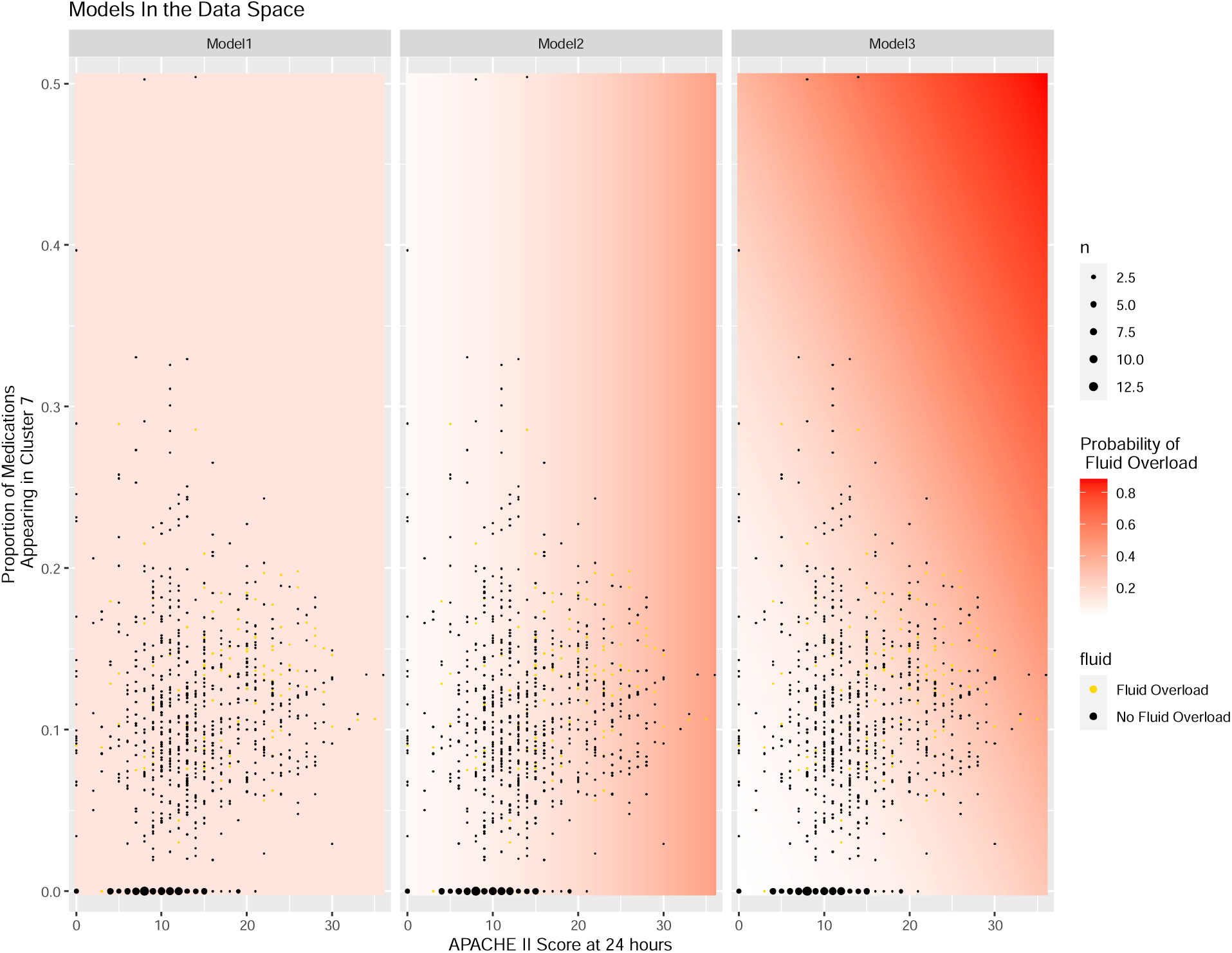
Visualization of significance of cluster 7 proportion and APACHE II score at 24 hours in logistic regression model in predicting fluid overload.

**Figure 10.**
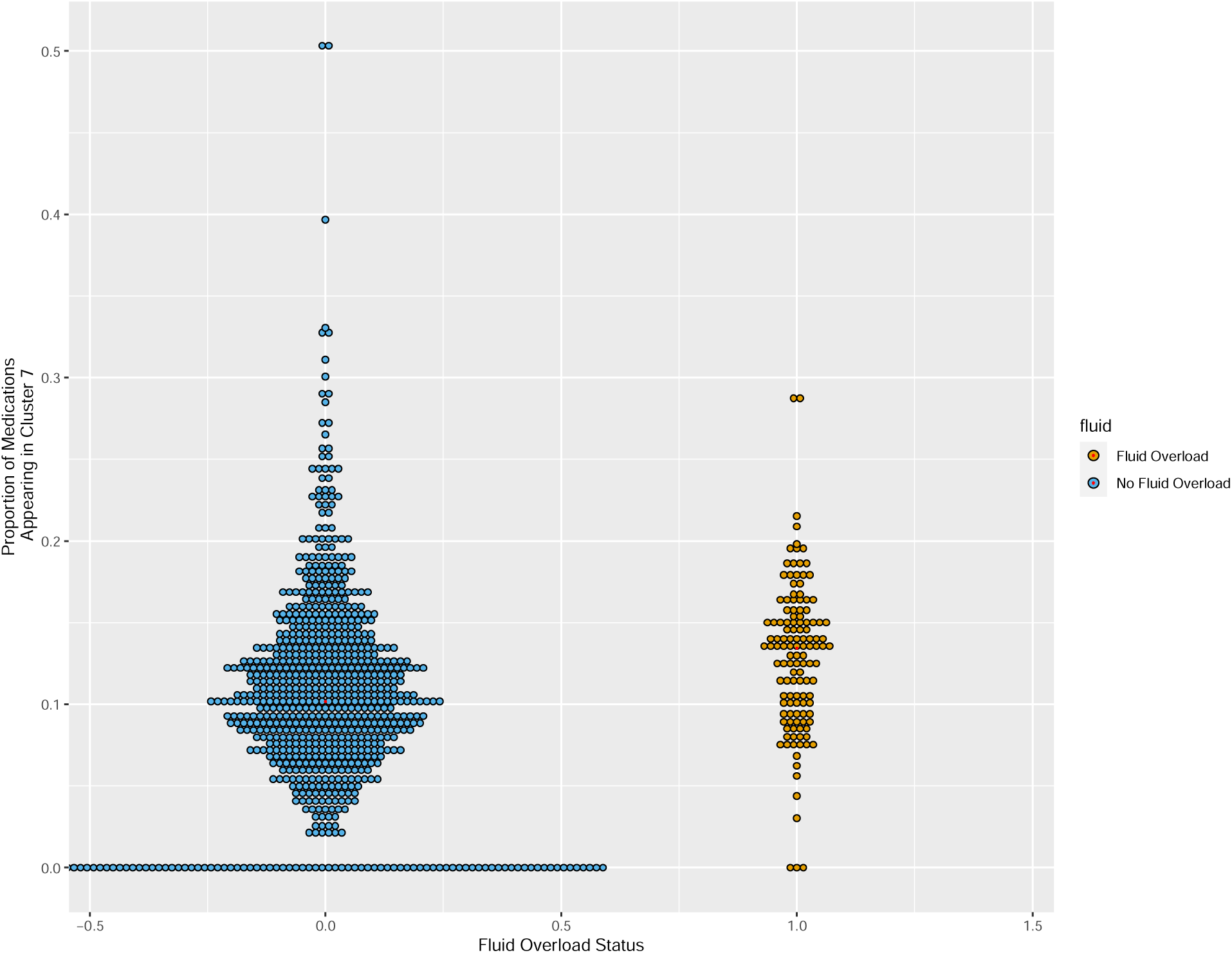
Distribution of patients in each group (non-fluid overload versus fluid overload) based on proportion of individual medications that appeared within Cluster 7.

**Figure 11.**
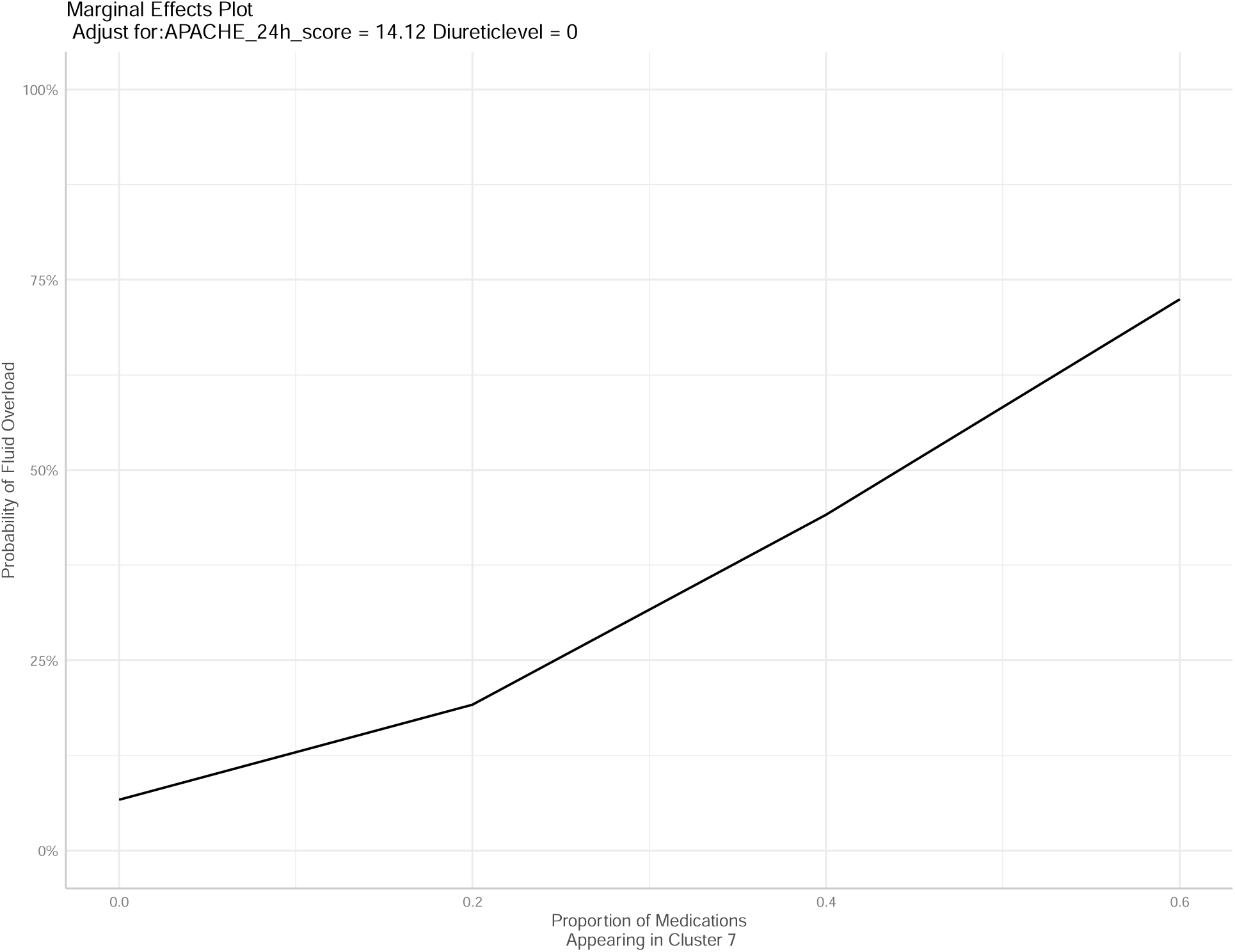
Marginal effect of cluster 7 proportion on fluid overload. Likelihood of an individual patient developing fluid overload, normalized to APACHE II score of 14 and no receipt of diuretics.

**Table 8.** Logistic Regressions for Prediction of Fluid Overload with/without Cluster 7 information.

| Model | Logistic Regression for Fluid Overload with APACHE II Score at 24 hours and Diuretic Level | Logistic Regression for Fluid Overload with APACHE II Score at 24 hours, Diuretic Level, and Proportion of Medications Appearing in Cluster 7 |
| --- | --- | --- |

|  | Estimated Value | P-value | Estimated Value | P-value |
| --- | --- | --- | --- | --- |
| (Intercept) | -3.00 | <0.0001 | -3.61 | <0.0001 |
| APACHE Score at 24 hours | 0.095 | <0.0001 | 0.01 | <0.0001 |
| Diuretic level (0-5) | -0.46 | 0.04 | -0.42 | 0.06 |
| Diuretic level (>5) | -17.25 | 0.98 | -17.18 | 0.98 |
| Proportion of Medications Appearing in Cluster 7 |  |  | 5.65 | 0.0004 |

Diuretic level 0-5: the patient received $\leq 5$ doses of a diuretic medication within the first 72 hours of ICU stay
Diuretic level >5: the patient received >5 doses of a diuretic medication within the first 71 hours of ICU stay

**Table 9.** Logistic Regressions for Prediction of Fluid Overload Using Proportion of Medications Appearing in Cluster 7 at Specified Time Periods.

| Model | Logistic Regression for Fluid Overload with APACHE II Score at 24 hours and Diuretic Level |  |
| --- | --- | --- |
|  | Estimated Value | P-value |
| (Intercept) | -3.34 | <0.0001 |
| APACHE Score at 24 hours | 0.035 | 0.052 |
| Diuretic level (0-5) | -0.49 | 0.037 |
| Diuretic level (>5) | -16.75 | 0.98 |
| Proportion of Medications Appearing in Cluster 7 in Day 1 of ICU Admission | 1.20 | 0.037 |
| Proportion of Medications Appearing in Cluster 7 in Day 2 of ICU Admission | 0.40 | 0.52 |
| Proportion of Medications Appearing in Cluster 7 in Day 3 of ICU Admission | 2.61 | <0.0001 |

Diuretic level 0-5: the patient received $\leq 5$ doses of a diuretic medication within the first 72 hours of ICU stay
Diuretic level >5: the patient received >5 doses of a diuretic medication within the first 71 hours of ICU stay

## DISCUSSION

This first of its kind analysis represents the integration of four novel concepts in the domain of data-driven medication use optimization: (1) the application of unsupervised machine learning methods to the entire MAR (including drug and dose), (2) incorporation of temporal data for medication administration, (3) fluid overload prediction in the ICU, and (4) application of the ICURx CDM. These methods identified a cluster of medications that both statistically and clinically correlated with fluid overload and serves as a proof-of-concept for future implementation studies evaluating how machine learning approaches could be integrated with real-time EHR data to provide predictions at the bedside.

Building on unsupervised machine learning methods that analyzed just the names (i.e., excluding dose, formulation, route) of medications received in the first 24 hours^17,23,32^, this is first time that unsupervised machine learning methods have been applied to the comprehensive medication regimen (i.e., including dose, formulation, route) up to 72 hours with an intent to explore how patterns in medication use relate to clinically relevant outcomes. These findings bring together two bodies of research: pharmacophenotyping as a means of identifying high risk patients, and fluid overload prediction using machine learning methods. In two prior pharmacophenotyping approaches, six pharmacophenotypes were identified that had unique patterns of associations with patient outcomes; however, these groupings were notably quite large with limited ability for clinical interpretation. Here, we found a more interpretable cluster, particularly when temporal data were added. Indeed, we observed that incorporating timing of medication administration into the unsupervised analysis provided further insight into development of fluid overload and specific medications, which may have a more substantial impact if given early within the ICU stay, and as such, marks an important exploration into the temporal component of medication administration as it relates to outcomes.

The discerned connection between the distinct IV medication cluster and the heightened risk of fluid overload underscores the need for a proactive and precise approach to medication management in the ICU. Such an approach may entail meticulous evaluation of factors such as timing, dosage, and the selection of specific IV medications, especially those falling within the identified subgroup. These findings align with other fluid overload prediction algorithms, which showed improvement when using machine learning and also that medications were highly ranked on feature importance graphs.^32–34^ While all of the clusters contained a similar list of medications (Figure 3), these clusters became more distinct and unique when the timing of medications was included in the original cluster development (Figure 2). When including the timing of medications, Cluster 7 was statistically correlated with fluid overload and also improved the prediction model for fluid overload. This may indicate that the timing of medications is more important than we realize and that artificial intelligence may represent the key to discovering these complex relationships. Cluster 7 had a higher number of medications administered within the first 24 hours compared to hours 25-48 or 49-72, which may be reflective of the importance of the first 24 hours of ICU stay. This temporal distribution of medication administration implies a potential association between early medication use and subsequent instances of fluid overload. Additionally, the medications that appeared within Cluster 7 included a large number of medications that clinically would be associated with fluid overload, including fluids themselves and continuous infusions such as vasopressors and inotropes. This adds to the validity of the clustering methods as the results are clinically correlated. As the proportion of medications appearing in Cluster 7 increased, patients were more likely to develop fluid overload as indicated in Figure 11, although this association is harder to discern when the proportion of medications appearing in Cluster 7 is >20% due to the limited number of patients who met this criteria. The greatest likelihood of developing fluid overload occurred when patients had between 10% and 20% of their medications matching the Cluster 7 list. Logistic regressions for various breakpoints of proportion of medications matching Cluster 7 can be found in the **Supplementary Appendix**. From a clinical perspective, this could allow for incorporation of clinical decision support by alerting practitioners to patients that have proportions of >10% matching Cluster 7, warranting increased monitoring and evaluation of need for concentrating medications, restricting fluids, or administration of diuretics. Overall, this lends more credence that medication data have a role in improving ICU modeling.^13,35^

Finally, this study represents further application of the ICURx CDM, which was employed to provide the algorithms with further information during the clustering process.^17,36^ While information from ICURx CDM was not included in the final methodology for the clustering process itself, it was used to provide further information about specific medications that were used for subsequent analyses (IV push versus continuous infusion and sorting of medications into classes). Results from this analysis could not fully evaluate the impact of medications in different volumes of fluid which could be important clinically (e.g., giving cefepime as an IV push medication compared to an intermittent infusion in 100mL of NS over 30 minutes) as the initial data set did not include administration rate.

Our study has several limitations including a small sample size and retrospective data collection. Additionally, due to the retrospective nature of this dataset, we chose a numerical definition for fluid overload as opposed to a clinical assessment which may have under-identified those with clinical fluid overload. Subsequently, bias may exist due to the availability of fluid balance data for the included patients. Causal relationships cannot be assessed by the current study, so it is unknown whether the high fluid overload observed in Patient Cluster 7 was partly caused by the unique distribution of medication patterns versus other factors (although notably, Cluster 7 shared similarities among groups). Additionally, while it is very encouraging that we were able to identify a cluster of medications that was statistically significantly associated with fluid overload through AI methods, at this time, there are multiple limitations in trying to apply this information to a clinical scenario. While there are many hypotheses generated from this information, including which medications may have an undiscovered temporal effect with fluid overload, there must be further research to apply this information at the bedside to have a clinical impact. Even with these limitations, this analysis marks the first time the complete medication profile has been incorporated into outcomes analysis for ICU patients. Future analyses with more granular cluster groupings or more programmed directives incorporating data from a myriad of ICUs and centers may improve the face validity. Artificial intelligence may provide clinical outcome prediction and serve as a supplement to clinicians given its ability to process large amounts of data in real-time.^37–40^ Ability to predict events in a critical care setting is highly relevant and desirable given the challenge to predict outcomes in patients with rapidly changing disease states and management.^41,42^ Overall, this evaluation is a first step and proof-of-concept exploration into how unsupervised clustering methods may be applied to ICU medications, particularly as it relates to the addition of temporal data.

## CONCLUSION

Unsupervised machine learning uncovered a distinctive cluster of IV medications that exhibited a robust correlation with the occurrence of fluid overload in the ICU setting. Delineating how medications and their administration timing may influence development of fluid overload using data driven methods may support future fluid overload prediction and mitigation strategies.

## Data Availability

All data produced in the present study are available upon reasonable request to the authors

## Acknowledgements

Data acquisition were supported by NC TraCS, funded by Grant Number UL1TR002489 from the National Center for Advancing Translations Sciences at the National Institutes of Health, and Data Analytics at the University of North Carolina Medical Center Department of Pharmacy.

## Supplemental Digital Information

**Table 1.**
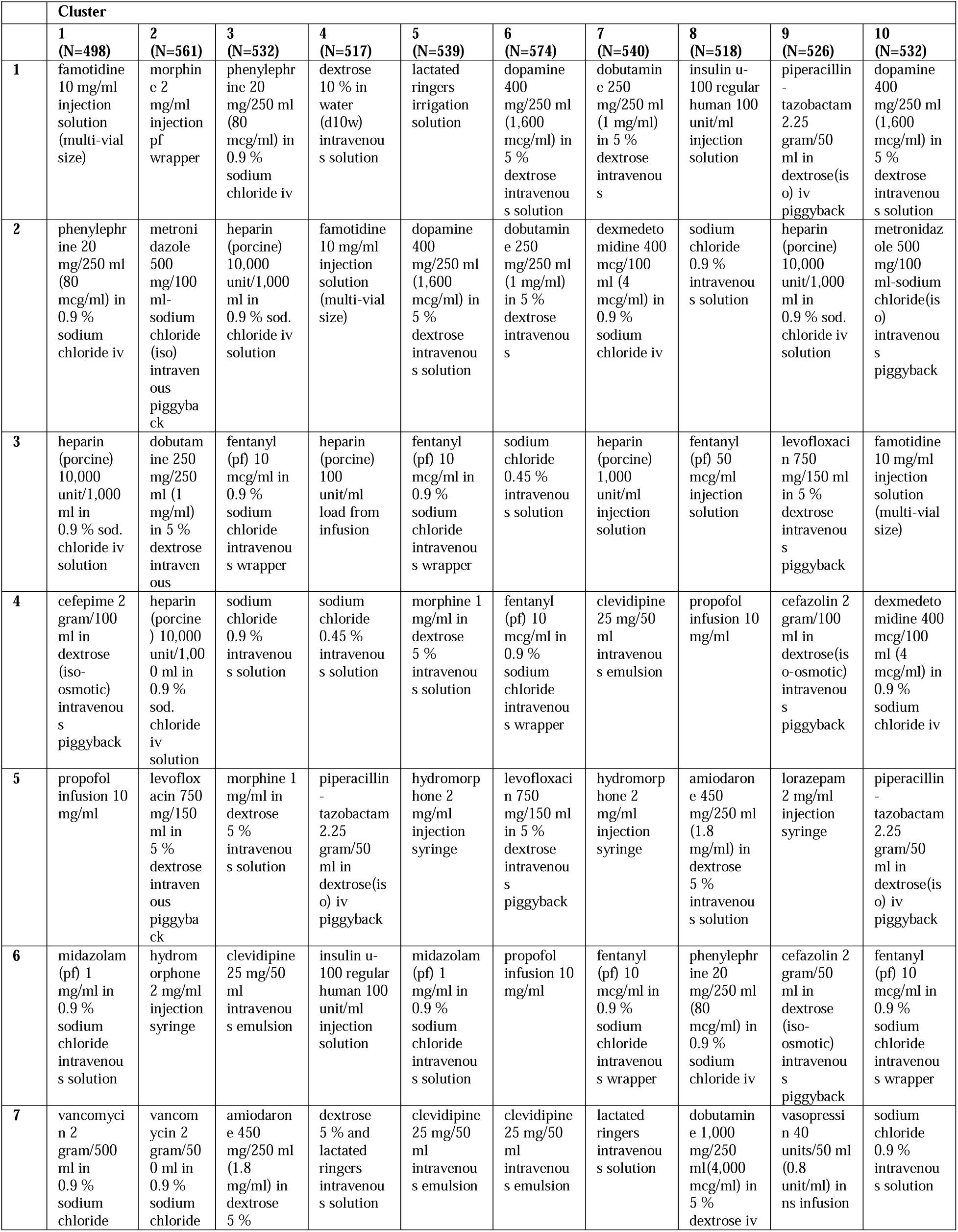
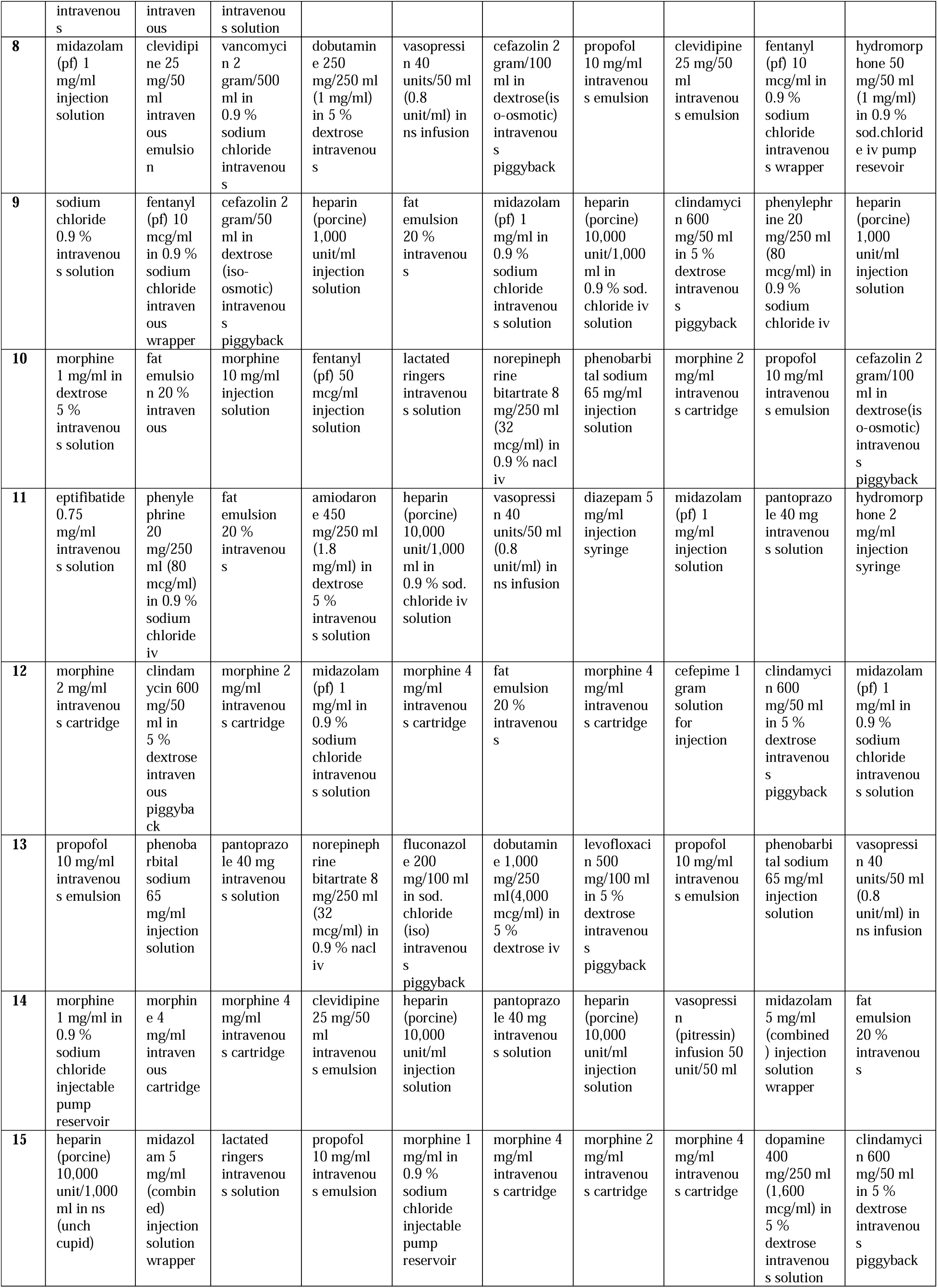
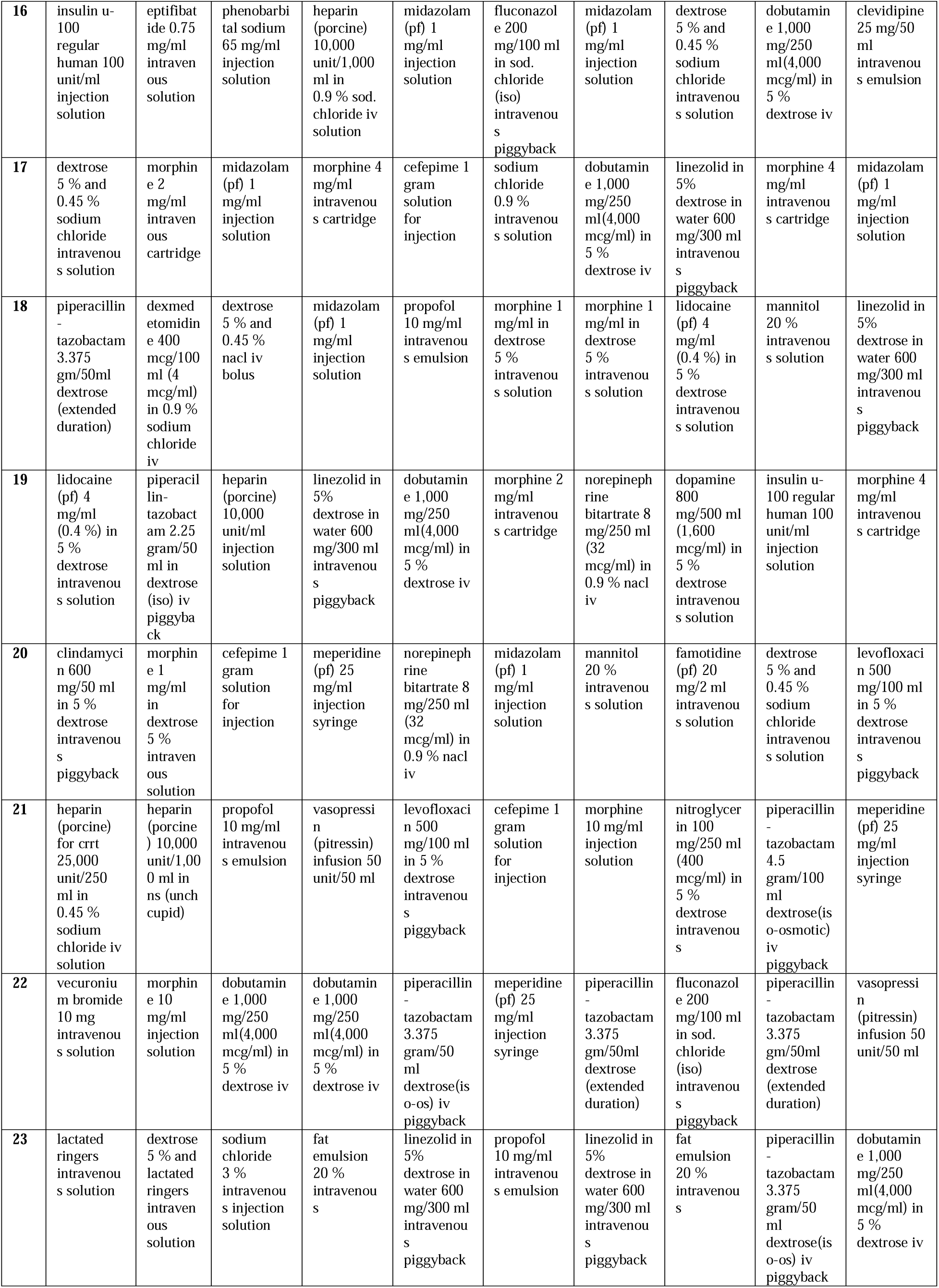
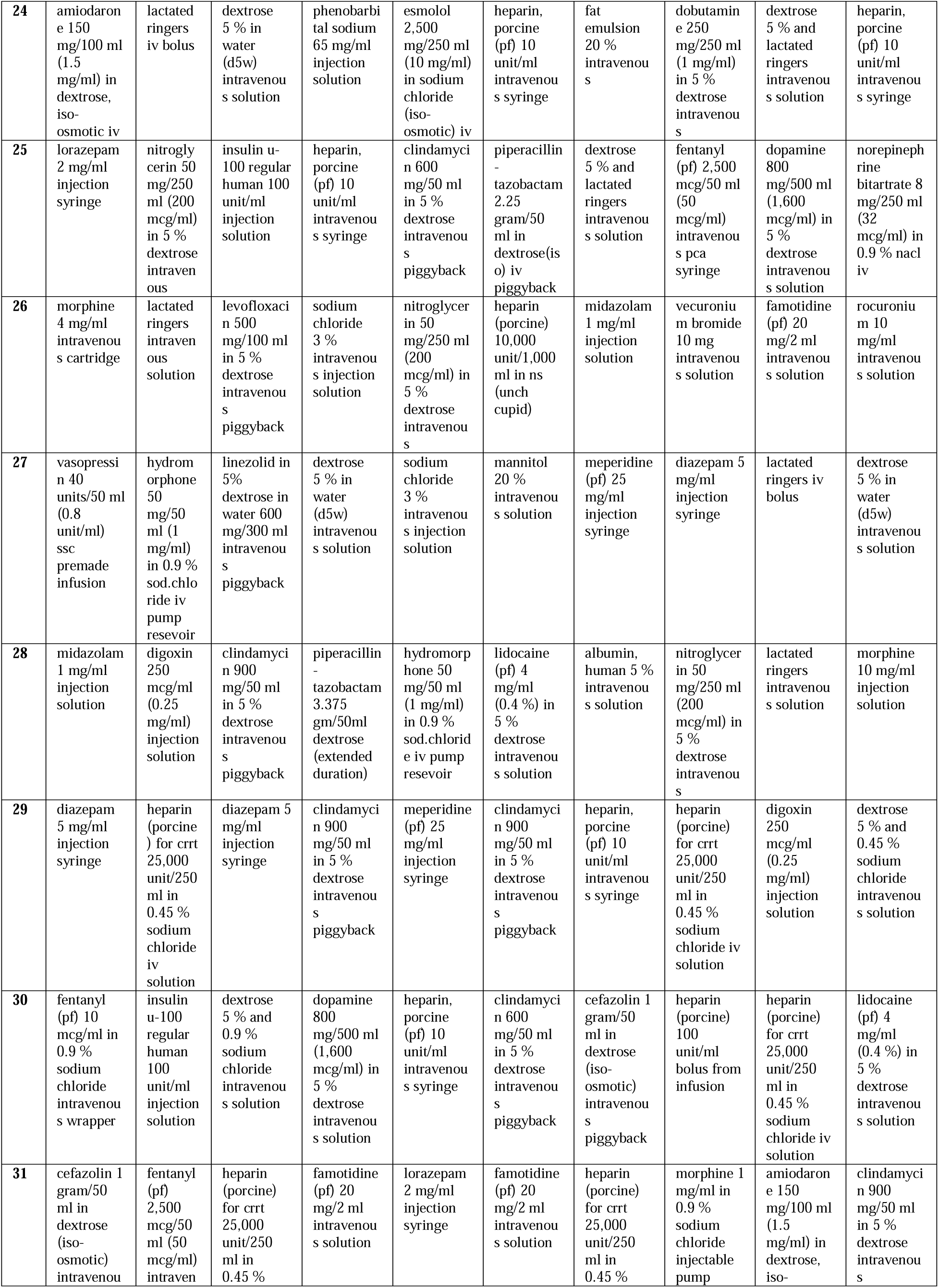
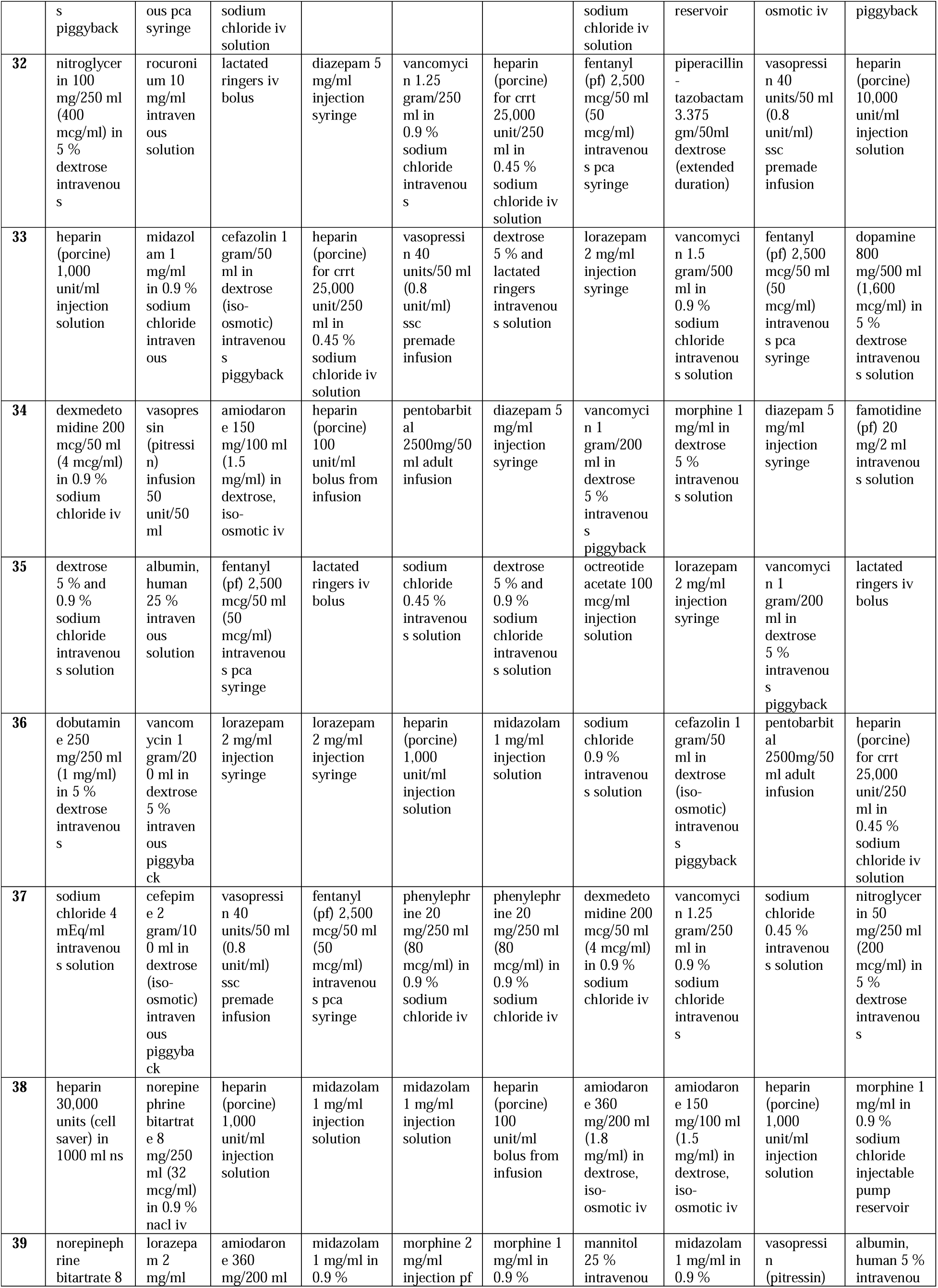
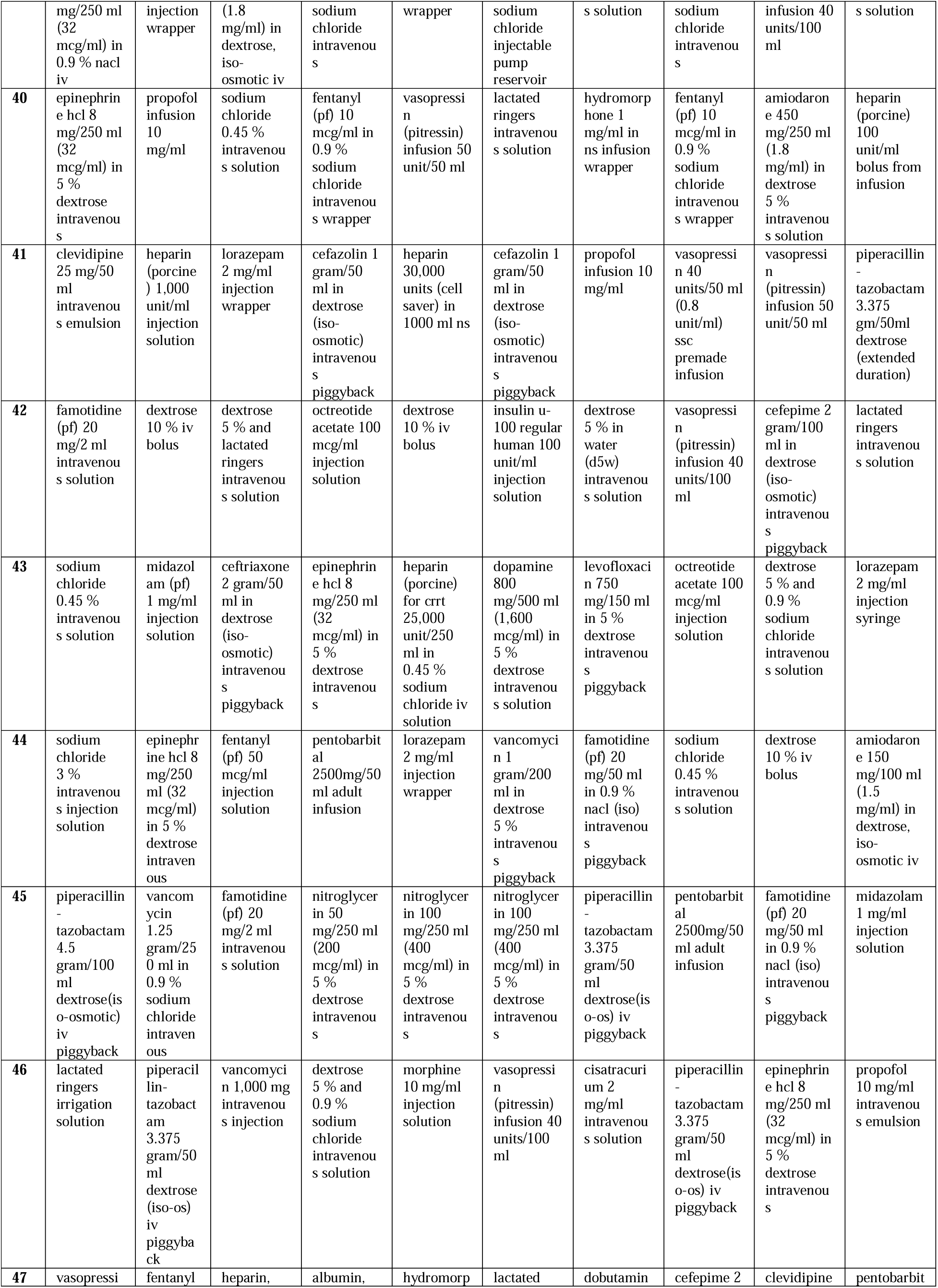
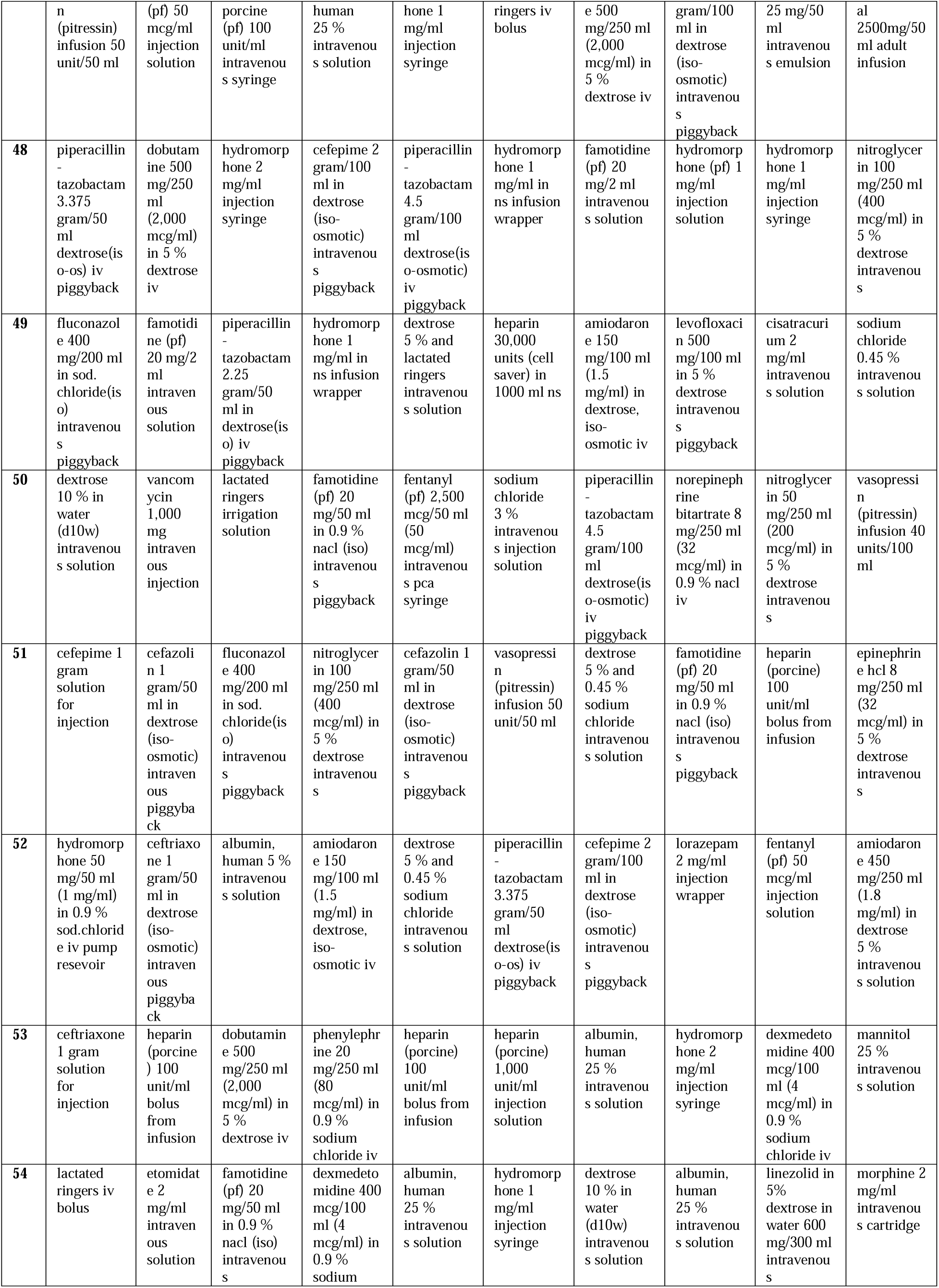
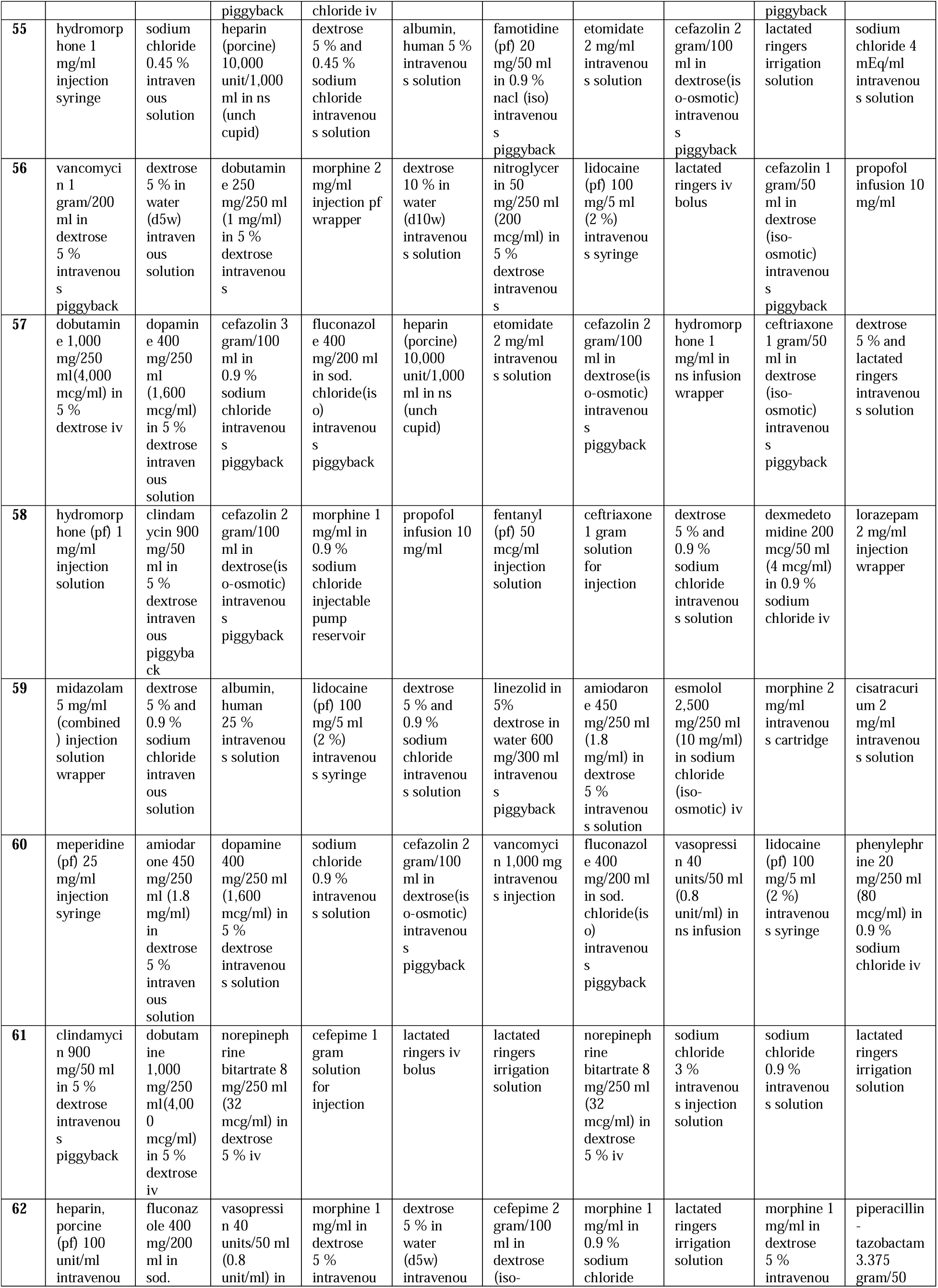
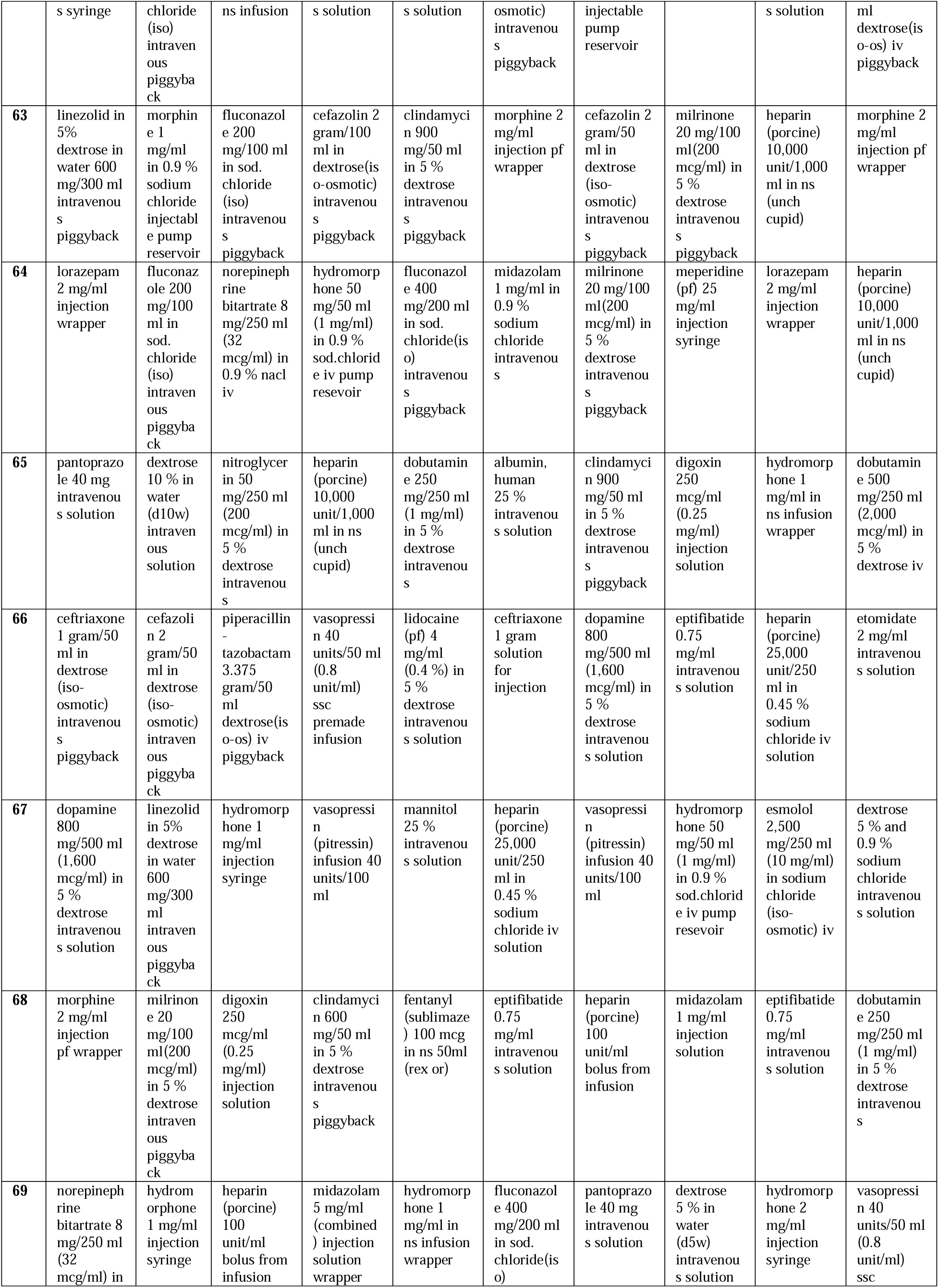
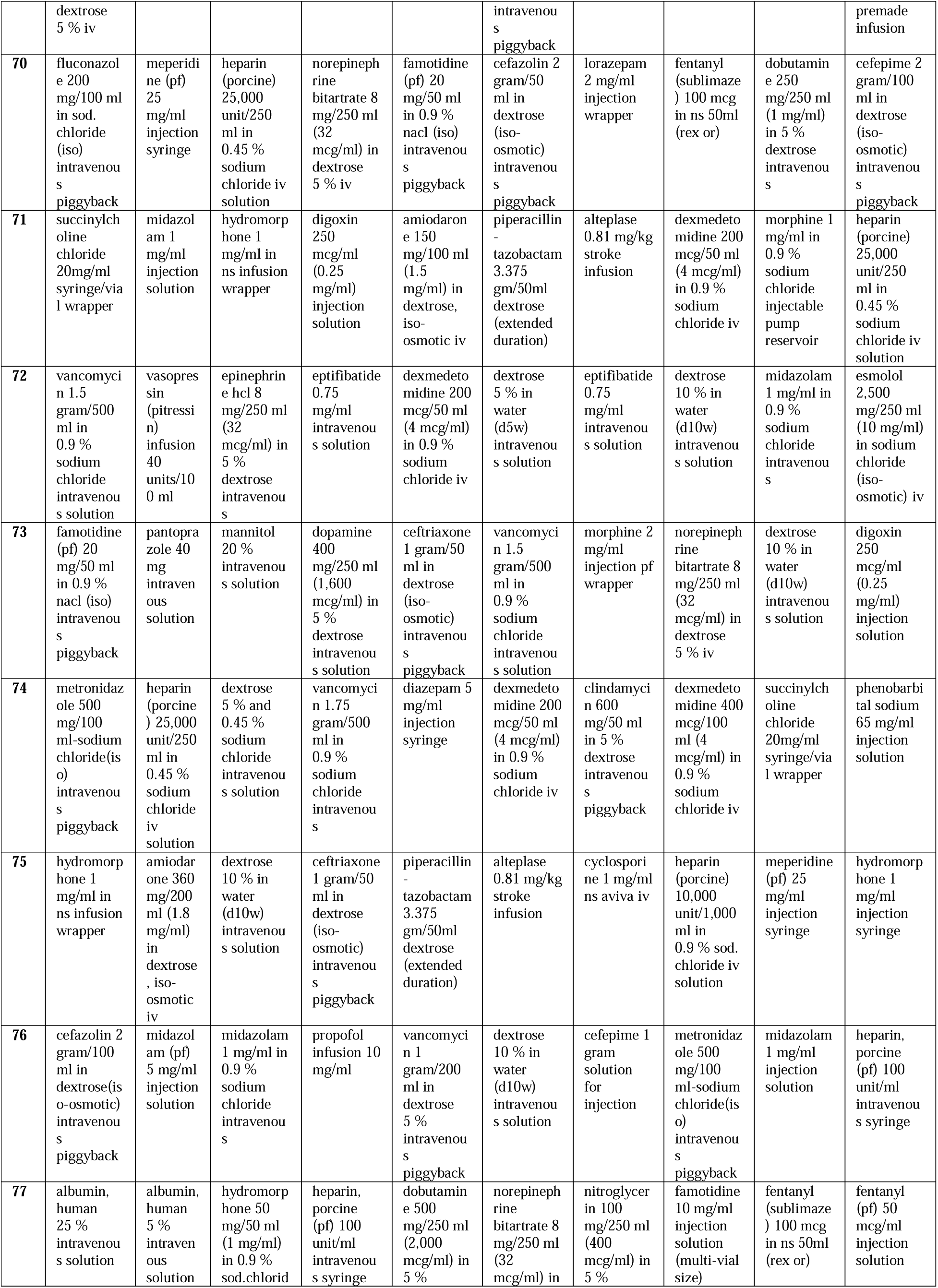
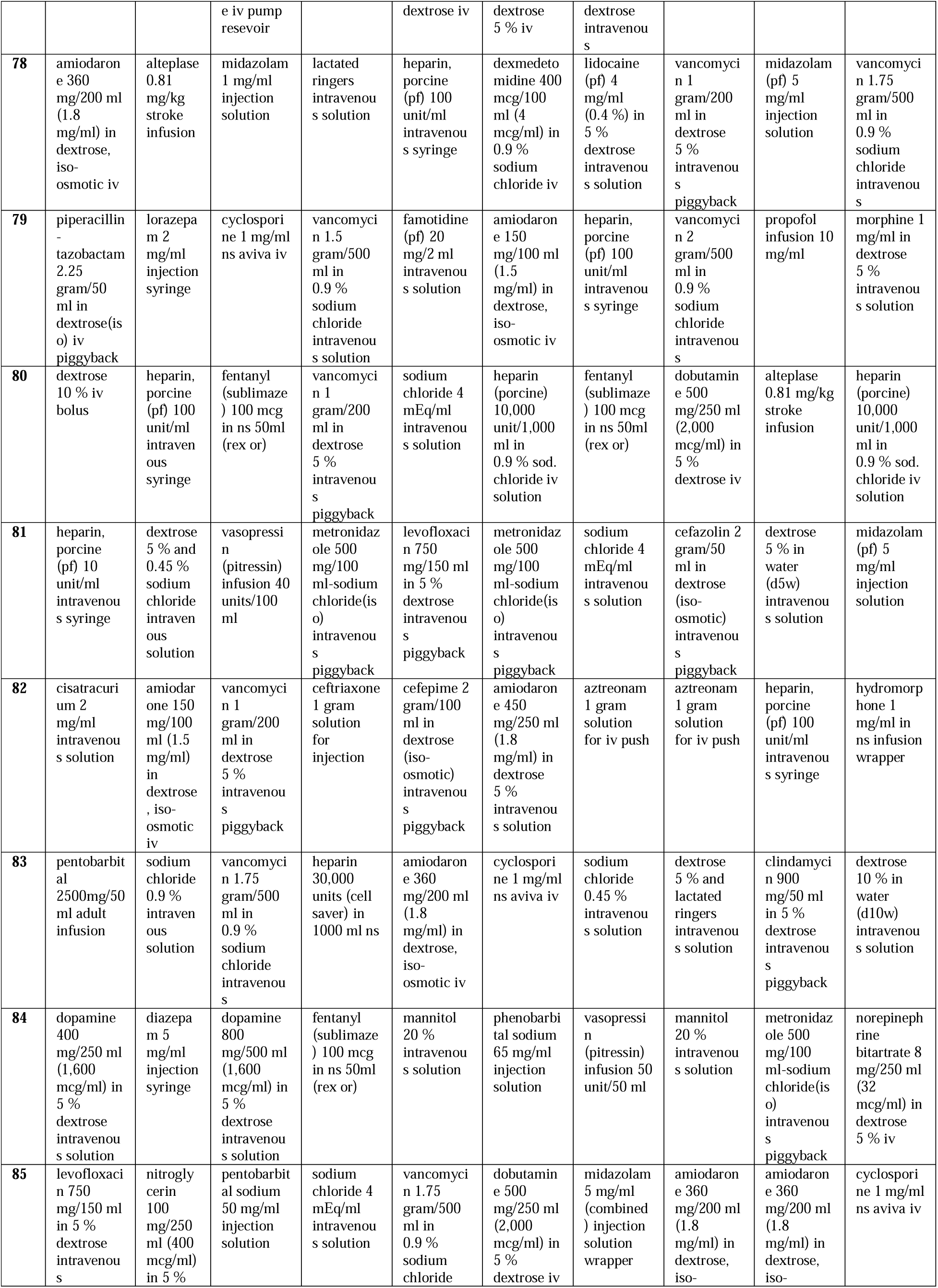
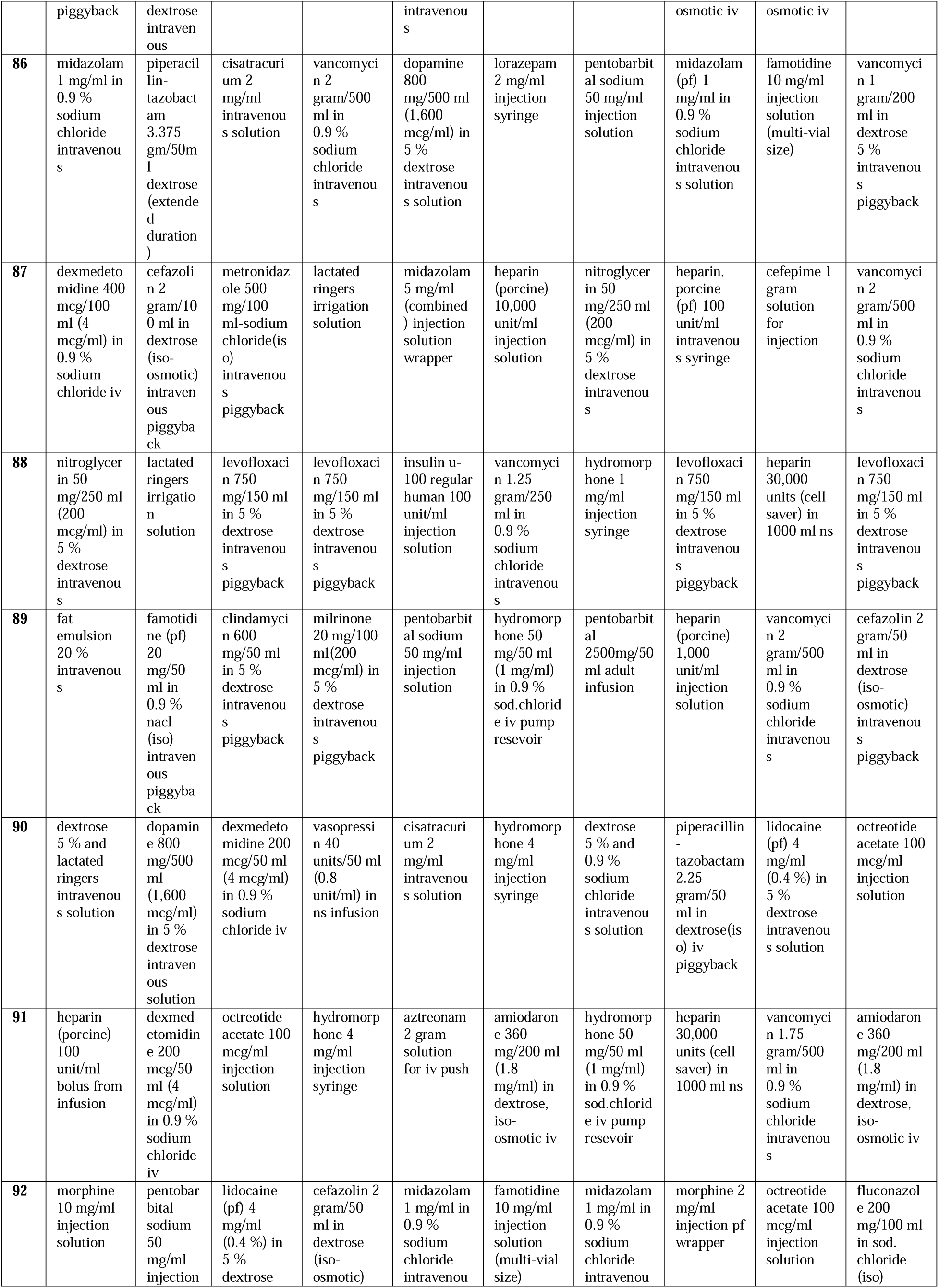
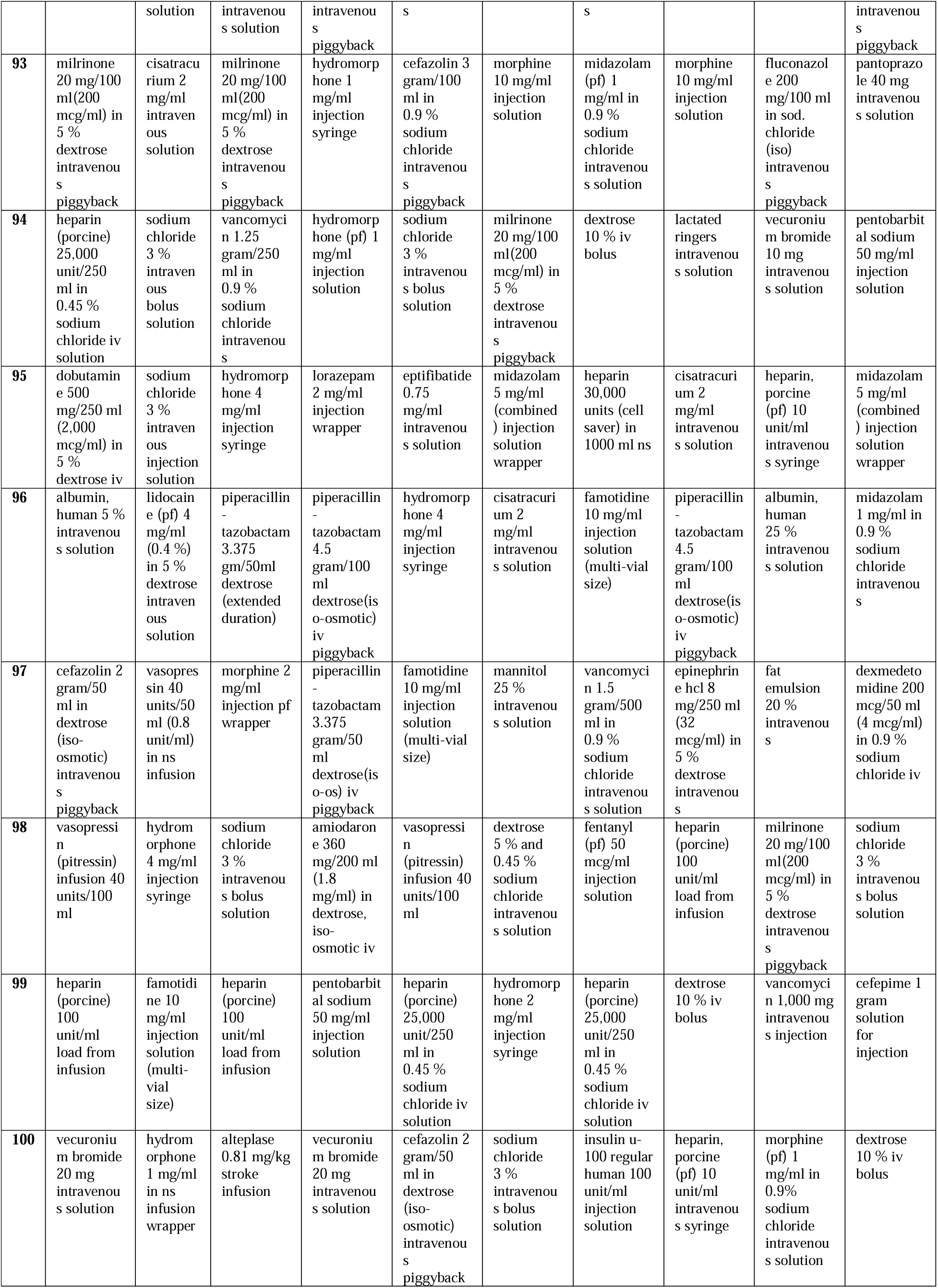
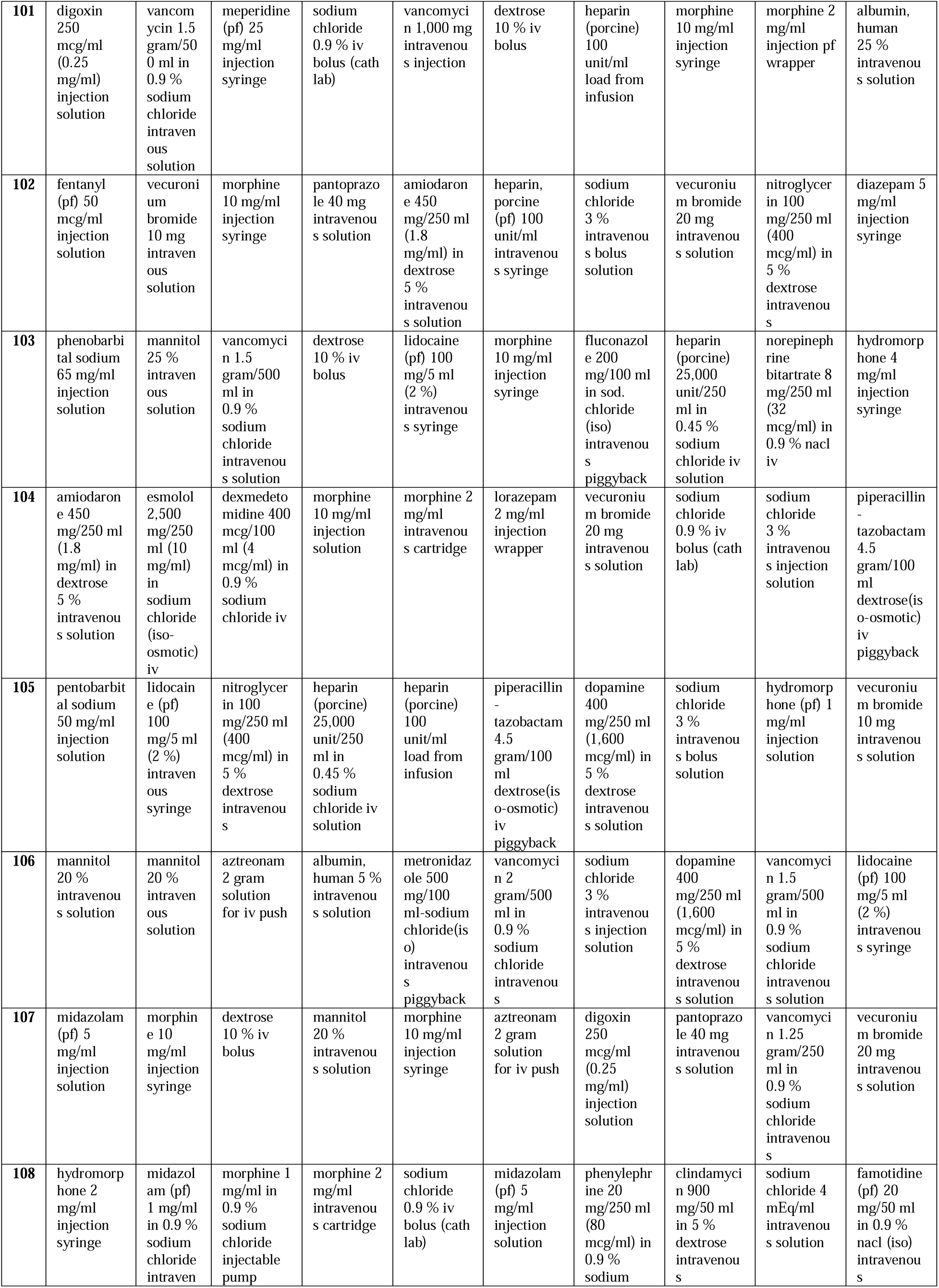
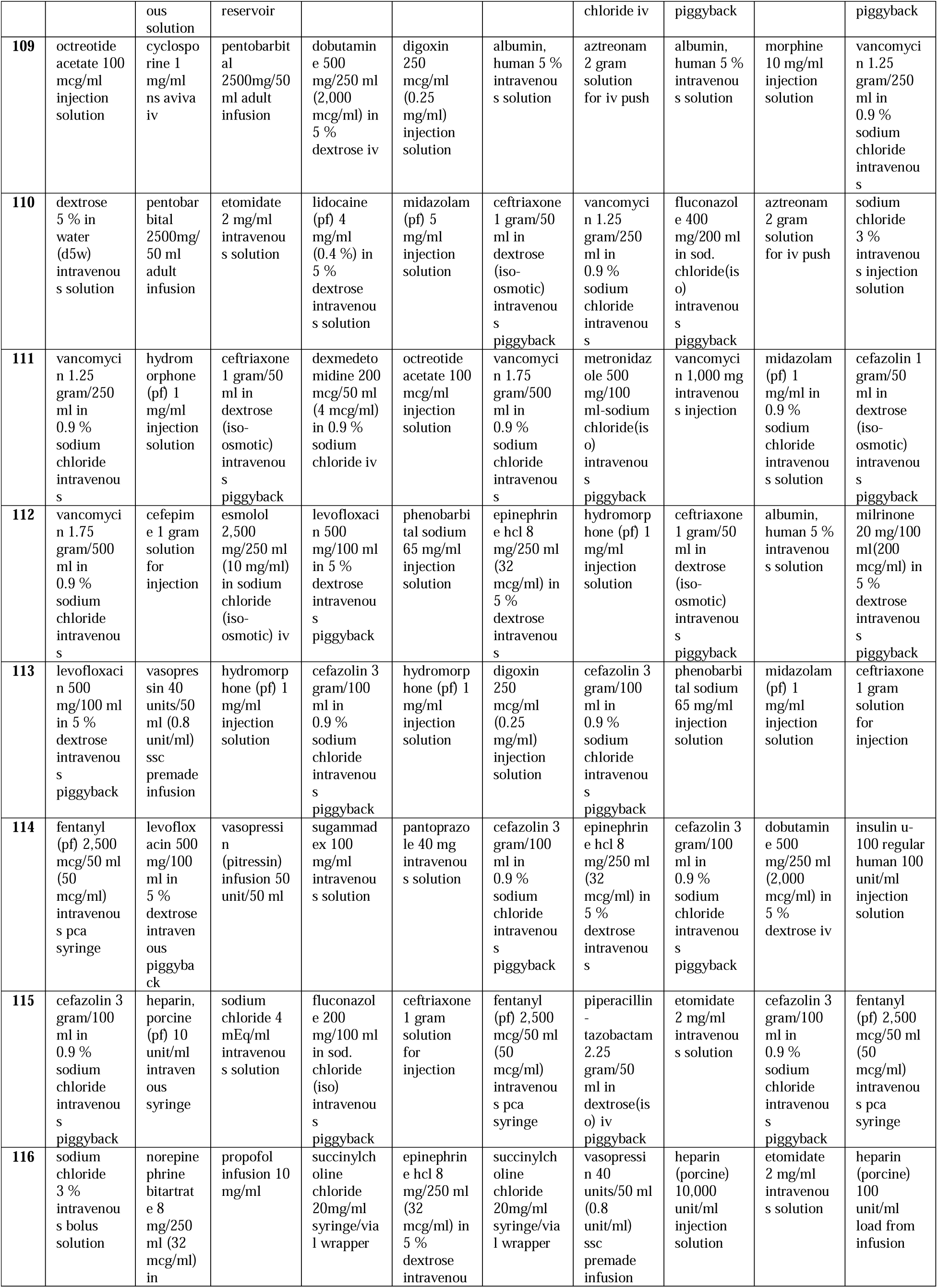
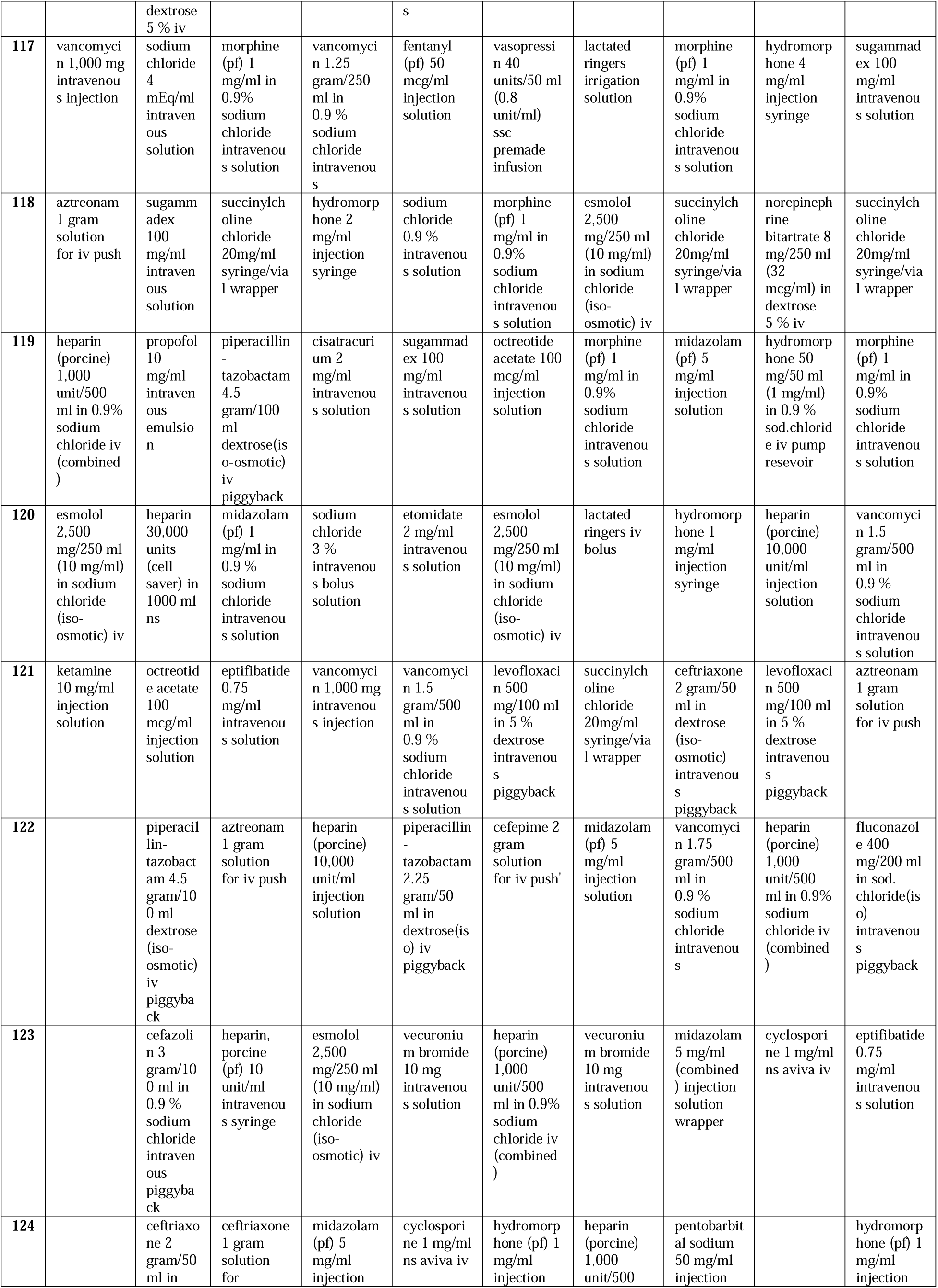
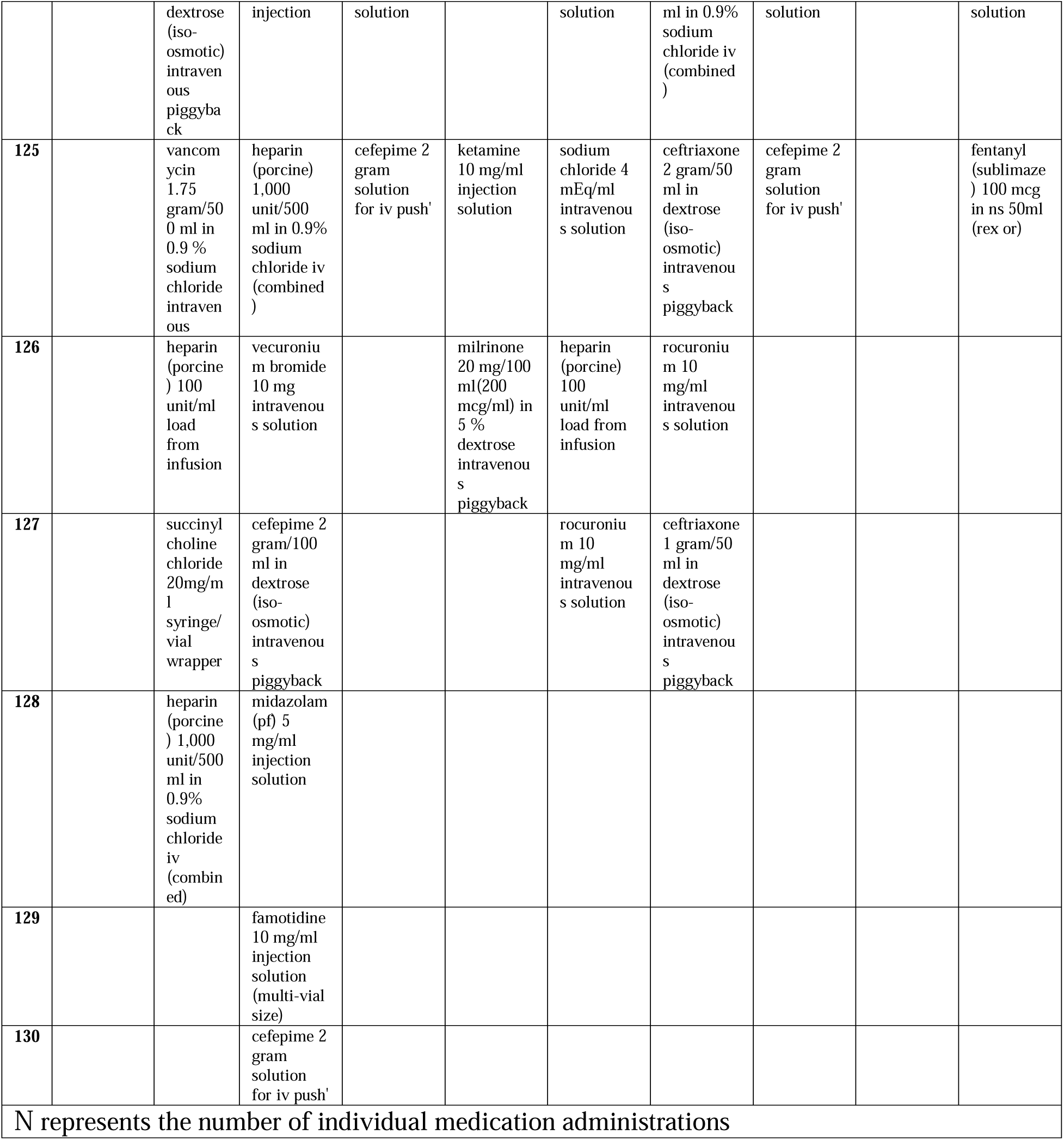
Composition of ten medication clusters identified with Restricted Boltzmann Machine.

**Table 2.**
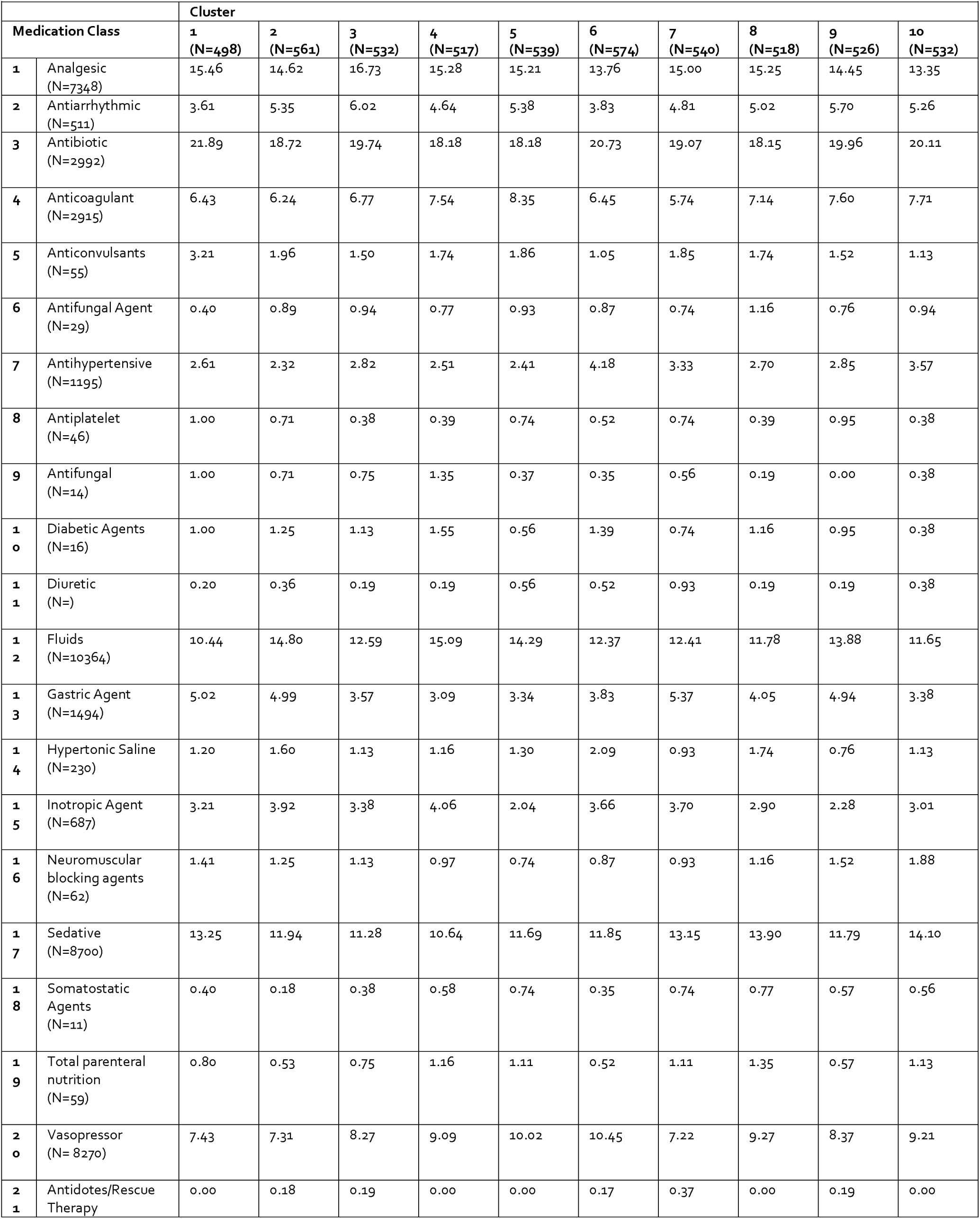
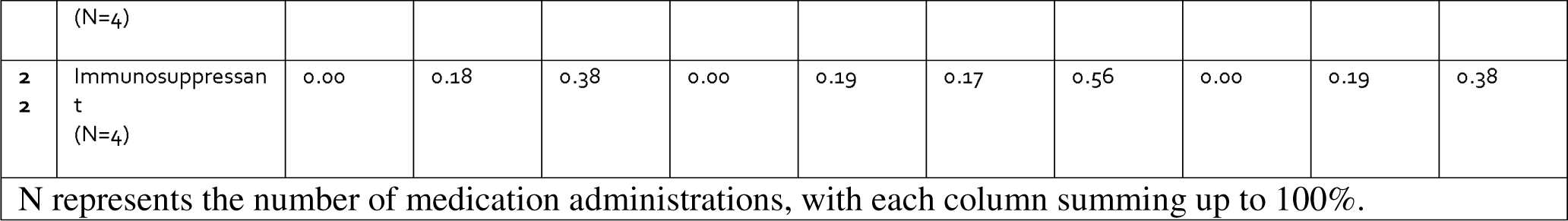
Distribution of medication classes within each medication cluster by total medication administrations.

**Table 3.** Logistic Regressions for Incidence of Fluid Overload with Varying Proportions of Medication Administrations Matching Cluster 7.

| Model | Logistic Regression for Fluid Overload with APACHE II Score at 24 hours and Diuretic Level |  |
| --- | --- | --- |
|  | Estimated Value | P-value |
| Greater than 10% of Medication Administrations Matching Cluster 7 | 0.68 | 0.00183 |
| Greater than 15% of Medication Administrations Matching Cluster 7 | 0.63 | 0.00481 |
| Greater than 20% of Medication Administrations Matching Cluster 7 | -0.29 | 0.601 |
| Greater than 30% of Medication Administrations Matching Cluster 7 | -16.2 | 0.9941 |

**Table 4.**
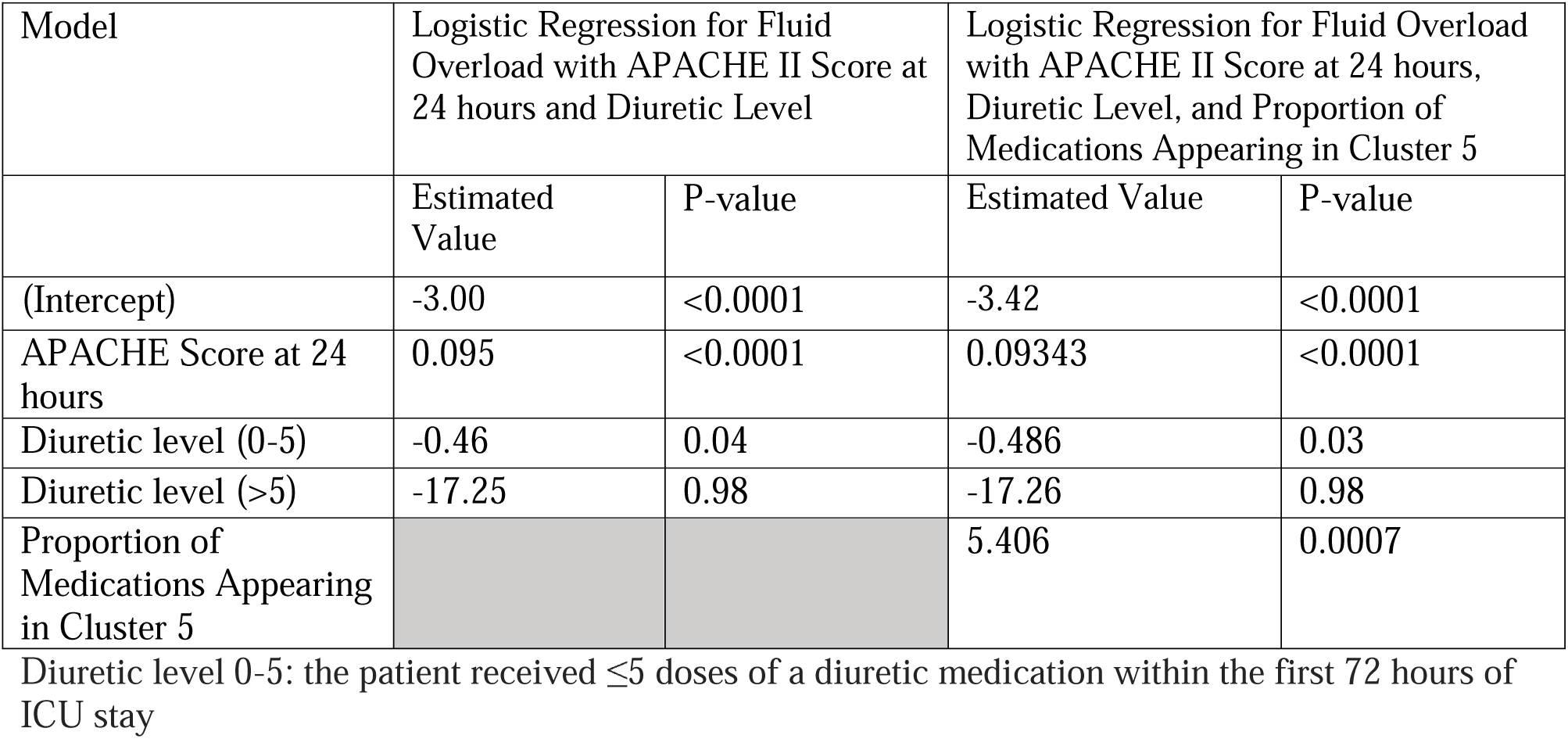
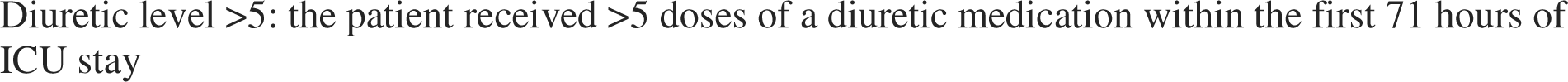
Logistic Regressions for Prediction of Fluid Overload with/without Cluster 5 information.

| Model | Logistic Regression for Fluid Overload with APACHE II Score at 24 hours and Diuretic Level |  | Logistic Regression for Fluid Overload with APACHE II Score at 24 hours, Diuretic Level, and Proportion of Medications Appearing in Cluster 5 |  |
| --- | --- | --- | --- | --- |
|  | Estimated Value | P-value | Estimated Value | P-value |
| (Intercept) | -3.00 | <0.0001 | -3.42 | <0.0001 |
| APACHE Score at 24 hours | 0.095 | <0.0001 | 0.09343 | <0.0001 |
| Diuretic level (0-5) | -0.46 | 0.04 | -0.486 | 0.03 |
| Diuretic level (>5) | -17.25 | 0.98 | -17.26 | 0.98 |
| Proportion of Medications Appearing in Cluster 5 |  |  | 5.406 | 0.0007 |

Diuretic level 0-5: the patient received $\leq 5$ doses of a diuretic medication within the first 72 hours of ICU stay
Diuretic level >5: the patient received >5 doses of a diuretic medication within the first 71 hours of ICU stay

**Figure 1.**
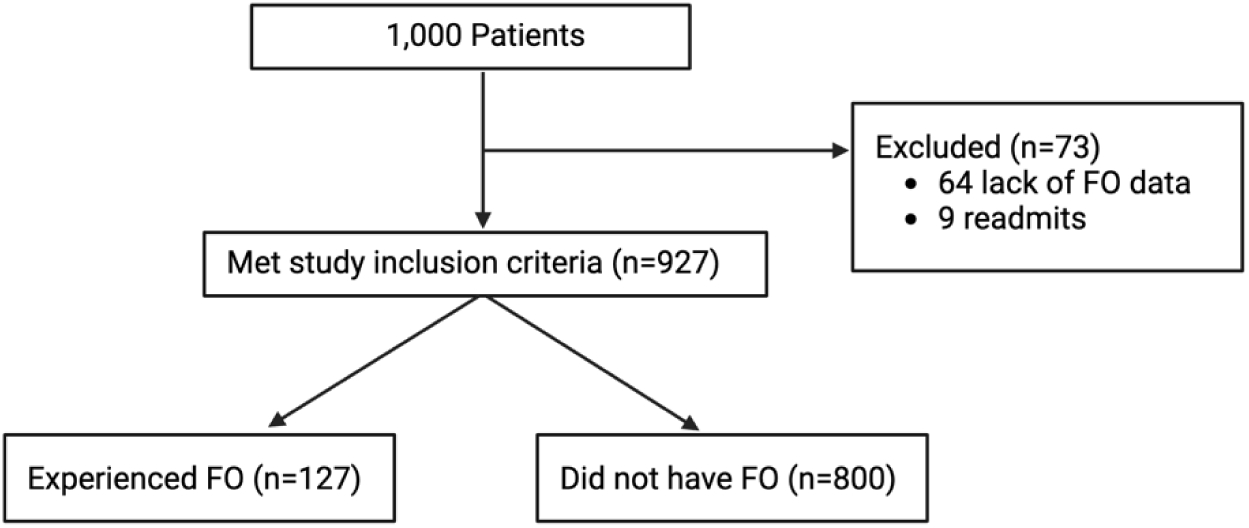
Consort Diagram of the Data Process Procedures

**Figure 2.**
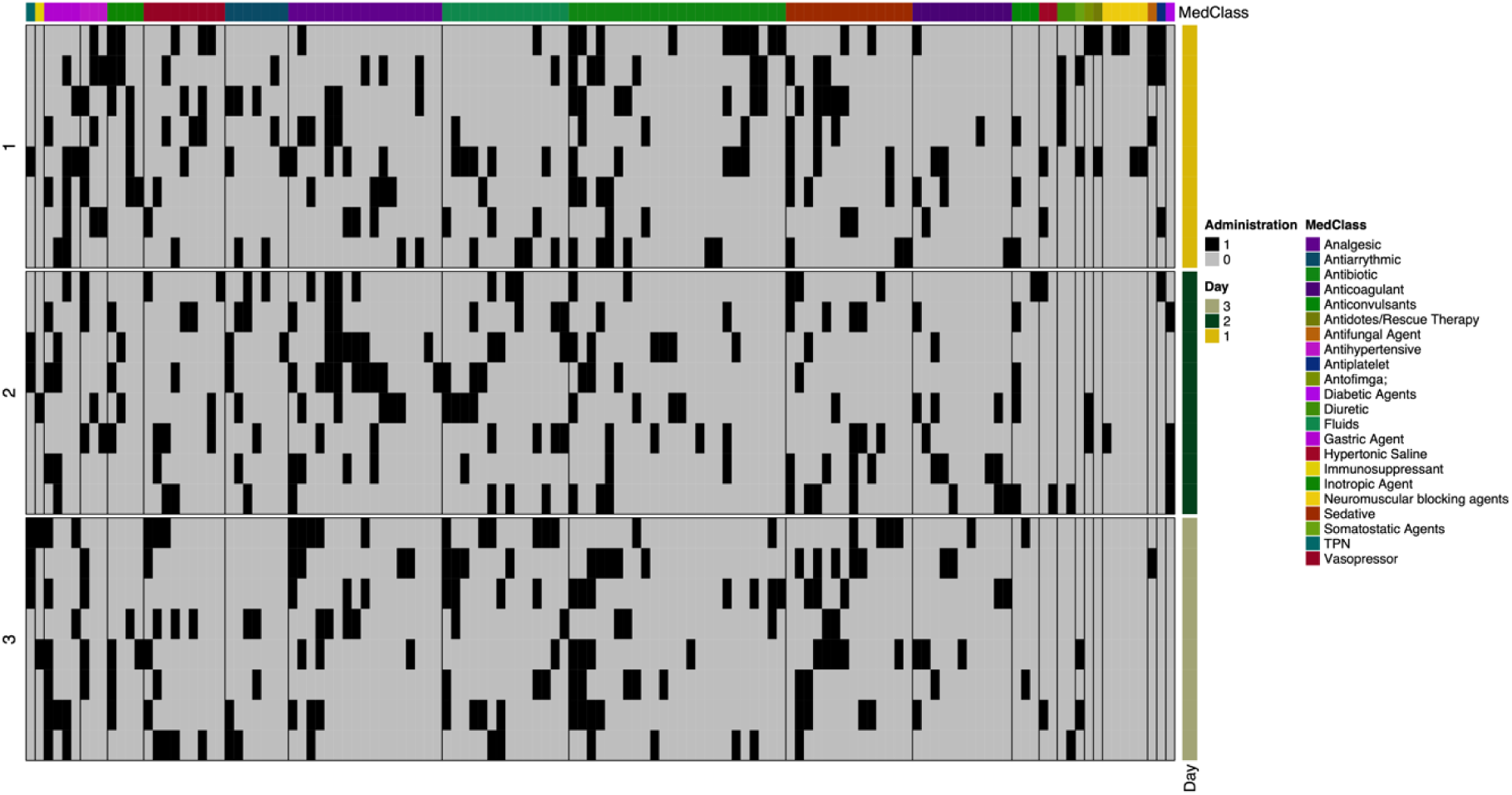
Cluster 7 medication administrations organized by timing of administration and medication class Black boxes indicate medication administration at specific time slots, while the width of column indicates amount of medications appearing within that class within the cluster Alternative version of Figure 6 from the main text

**Figure 3.**
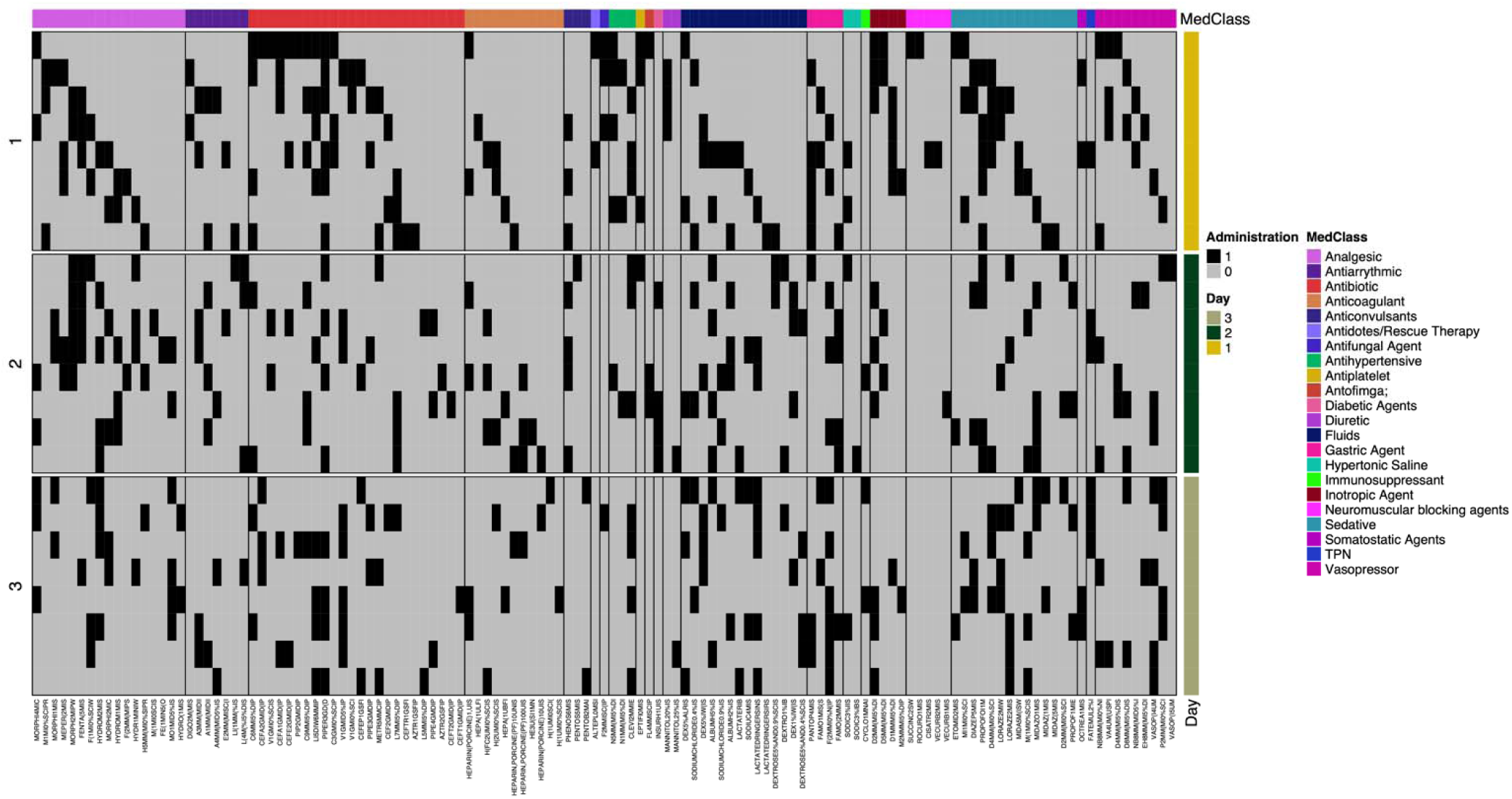
Cluster 7 medication administrations organized by timing of administration and medication class with classes in alphabetical order Black boxes indicate medication administration at specific time slots, while the width of column indicates amount of medications appearing within that class within the cluster Alternative version of Figure 6 from the main text

**Figure 4.**
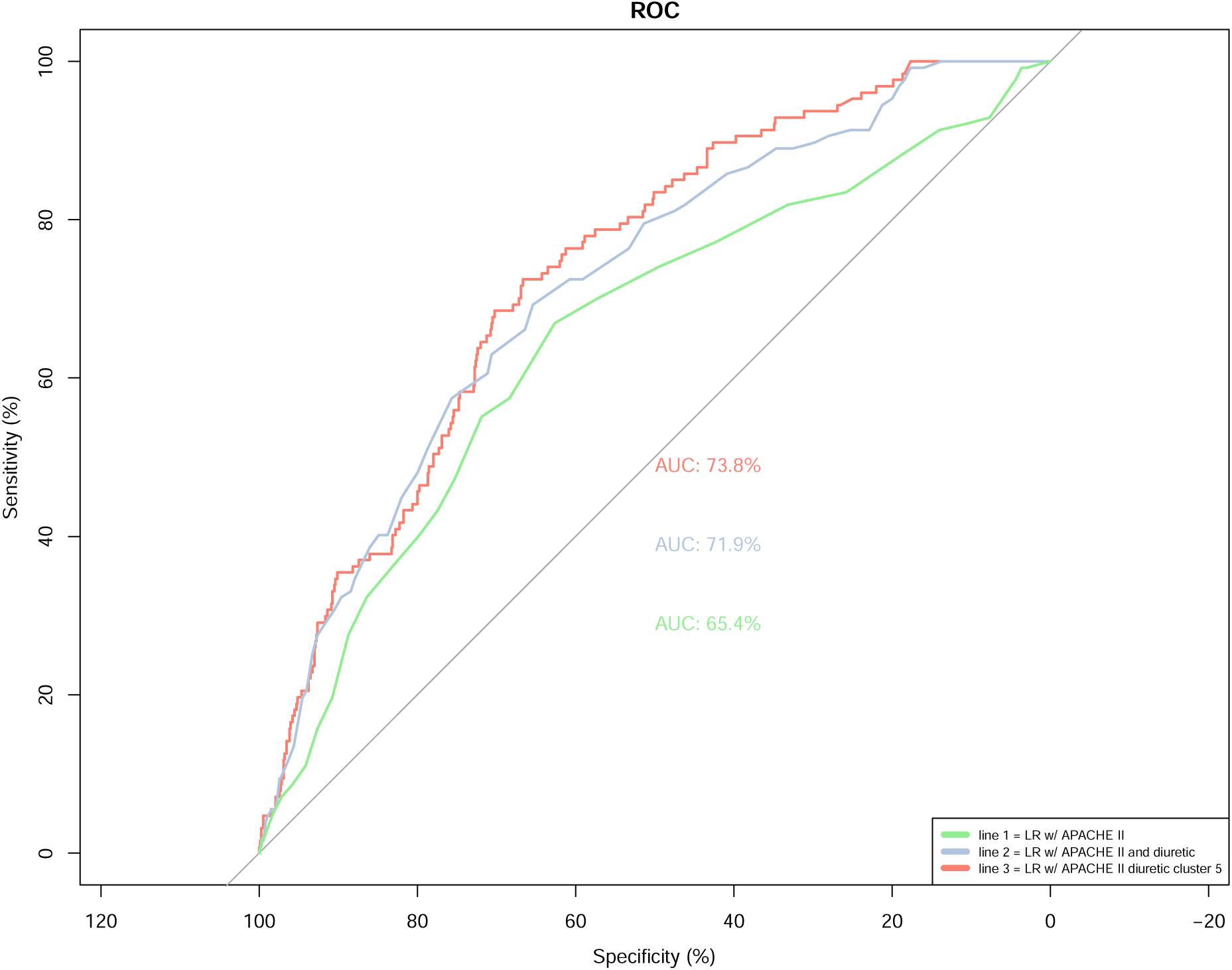
Logistic regression model for Cluster 5 Logistic regression for incidence of fluid overload, including Cluster 5, APACHE II score, and diuretic level

## References

1. Carr JR, Hawkins WA, Newsome AS, et al: Fluid stewardship of maintenance Intravenous fluids. J Pharm Pract 2021:8971900211008261

2. Hawkins WA, Smith SE, Newsome AS, et al: Fluid stewardship during critical illness: a call to action. J Pharm Pract 2020; 33(6):863–873

3. Bissell BD, Laine ME, Thompson Bastin ML, et al: Impact of protocolized diuresis for de-resuscitation in the intensive care unit. Crit Care 2020; 24(1):70

4. Jones TW, Chase AM, Bruning R, et al: Early diuretics for de-resuscitation in septic patients with left ventricular dysfunction. Clin Med Insights Cardiol 2022; 16:11795468221095875

5. Hawkins WA, Butler SA, Poirier N, et al: From theory to bedside: implementation of fluid stewardship in a medical ICU pharmacy practice. Am J Health Syst Pharm 2022 Jun 7;79(12):984–992.

6. Bissell BD, Donaldson JC, Morris PE, et al: A narrative review of pharmacologic de-resuscitation in the critically ill. J Crit Care 2020; 59:156–162

7. Messmer AS, Moser M, Zuercher P, et al: Fluid overload phenotypes in critical illness-a machine learning approach. J Clin Med 2022; 11(2)

8. Zhang Z, Ho KM, Hong Y: Machine learning for the prediction of volume responsiveness in patients with oliguric acute kidney injury in critical care. Crit Care 2019; 23(1):112

9. Olney WJ, Chase AM, Hannah SA, et al: Medication regimen complexity score as an indicator of fluid balance in critically ill Patients. J Pharm Pract 2021:897190021999792

10. Sikora A, Ayyala D, Rech MA, et al: Impact of pharmacists to improve patient care in the critically ill: a large multicenter analysis using meaningful metrics with the medication regimen complexity-ICU (MRC-ICU) score. Crit Care Med 2022; 50(9):1318–1328

11. Newsome AS, Smith SE, Olney WJ, et al: Multicenter validation of a novel medication-regimen complexity scoring tool. Am J Health Syst Pharm 2020; 77(6):474–478

12. Newsome AS, Anderson D, Gwynn ME, et al: Characterization of changes in medication complexity using a modified scoring tool. Am J Health Syst Pharm 2019; 76(Supplement_4):S92–s95

13. Gwynn ME, Poisson MO, Waller JL, et al: Development and validation of a medication regimen complexity scoring tool for critically ill patients. Am J Health Syst Pharm 2019; 76(Supplement_2):S34–s40

14. Al-Mamun MA, Brothers T, Newsome AS: Development of Machine Learning Models to Validate a Medication Regimen Complexity Scoring Tool for Critically Ill Patients. Ann Pharmacother 2021; 55(4):421–429

15. Smith SE, Shelley R, Sikora A: Medication regimen complexity vs patient acuity for predicting critical care pharmacist interventions. Am J Health Syst Pharm 2022; 79(8):651–655

16. Webb AJ, Rowe S, Newsome AS: A descriptive report of the rapid implementation of automated MRC-ICU calculations in the EMR of an academic medical center. Am J Health Syst Pharm 2022; 79(12):979–983

17. Newsome AS, Smith SE, Olney WJ, et al: Medication regimen complexity is associated with pharmacist interventions and drug-drug interactions: A use of the novel MRC-ICU scoring tool. Journal of the American College of Clinical Pharmacy. 2020; 3(1):47–56

18. Sanchez P, Voisey JP, Xia T, et al: Causal machine learning for healthcare and precision medicine. R Soc Open Sci 2022; 9(8):220638

19. Iwase S, Nakada TA, Shimada T, et al: Prediction algorithm for ICU mortality and length of stay using machine learning. Sci Rep 2022; 12(1):12912

20. Beil M, Sviri S, Flaatten H, et al: On predictions in critical care: The individual prognostication fallacy in elderly patients. J Crit Care 2021; 61:34–38

21. Lovejoy CA, Buch V, Maruthappu M: Artificial intelligence in the intensive care unit. Crit Care 2019; 23(1):7

22. Gutierrez G: Artificial Intelligence in the Intensive Care Unit. Crit Care 2020; 24(1):101

23. Goh KH, Wang L, Yeow AYK, et al: Artificial intelligence in sepsis early prediction and diagnosis using unstructured data in healthcare. Nat Commun 2021; 12(1):711

24. Hyland SL, Faltys M, Huser M, et al: Early prediction of circulatory failure in the intensive care unit using machine learning. Nat Med 2020; 26(3):364–373

25. DeGrave AJ, Janizek JD, Lee SI: AI for radiographic COVID-19 detection selects shortcuts over signal. medRxiv 2020

26. Nguyen D, Ngo B, vanSonnenberg E: AI in the Intensive Care Unit: Up-to-Date Review. J Intensive Care Med 2021; 36(10):1115–1123

27. Yoon JH, Pinsky MR, Clermont G: Artificial Intelligence in Critical Care Medicine. Crit Care 2022; 26(1):75

28. Farion KJ, Wilk S, Michalowski W, et al: Comparing predictions made by a prediction model, clinical score, and physicians: pediatric asthma exacerbations in the emergency department. Appl Clin Inform 2013; 4(3):376–391

29. Feng JZ, Wang Y, Peng J, et al: Comparison between logistic regression and machine learning algorithms on survival prediction of traumatic brain injuries. J Crit Care 2019; 54:110–116

30. World Medical Association: World Medical Association Declaration of Helsinki: Ethical principles for medical research involving human subjects. JAMA 2023: 312:2191–2194.

31. Von Elm E AD, Egger M, et al: STROBE Initiative: Strengthening the reporting of observational studies in epidemiology (STROBE) statement: Guidelines for reporting observational studies. BMJ 2007; 335:806–808.

32. Bouchard J, Soroko SB, Chertow GM, et al: Fluid accumulation, survival and recovery of kidney function in critically ill patients with acute kidney injury. Kidney Int 2009; 76(4):422–427

33. Carr JR, Hawkins WA, Newsome AS, et al: Fluid Stewardship of Maintenance Intravenous Fluids. J Pharm Pract 2022; 35(5):769–782

34. Malbrain M, Van Regenmortel N, Saugel B, et al: Principles of fluid management and stewardship in septic shock: it is time to consider the four D’s and the four phases of fluid therapy. Ann Intensive Care 2018; 8(1):66

35. Claure-Del Granado R, Mehta RL: Fluid overload in the ICU: evaluation and management. BMC Nephrol 2016; 17(1):109

36. O’Connor ME, Prowle JR: Fluid Overload. Crit Care Clin 2015; 31(4):803–821

37. Chen T, Guestrin C. XGBoost: A Scalable Tree Boosting System. In: Proceedings of the 22nd ACM SIGKDD International Conference on Knowledge Discovery and Data Mining [Internet]. New York, NY, USA: ACM; 2016. p. 785–94. Available from: http://doi.acm.org/10.1145/2939672.2939785

38. Cortes C, Vapnik V. Support-vector networks. Machine learning. 1995;20(3):273–97.

39. Ho TK. Random decision forests. In: Proceedings of 3rd international conference on document analysis and recognition. 1995. p. 278–82.

40. Rubin DB: Multiple imputation for nonresponse in surveys. Hoboken, N.J. ;, Wiley-Interscience, 2004

41. Topol EJ: Deep medicine : how artificial intelligence can make healthcare human again. First edition. ed. New York, Basic Books,, 2019, p^pp 1 online resource

42. Kahneman D, Sibony O, Sunstein CR: Noise : a flaw in human judgment. First edition. Edition. New York, Little, Brown Spark, 2021

43. D’Hondt E, Ashby TJ, Chakroun I, et al: Identifying and evaluating barriers for the implementation of machine learning in the intensive care unit. Commun Med (Lond) 2022; 2(1):162

44. van de Sande D, van Genderen ME, Huiskens J, et al: Moving from bytes to bedside: a systematic review on the use of artificial intelligence in the intensive care unit. Intensive Care Med 2021; 47(7):750–760

45. Moss L, Corsar D, Shaw M, et al: Demystifying the Black Box: The Importance of Interpretability of Predictive Models in Neurocritical Care. Neurocrit Care 2022; 37(Suppl 2):185–191

46. The Lancet Respiratory M: Opening the black box of machine learning. Lancet Respir Med 2018; 6(11):801

47. Malbrain M, Martin G, Ostermann M: Everything you need to know about deresuscitation. Intensive Care Med 2022; 48(12):1781–1786

48. Gelbart B, Serpa Neto A, Stephens D, et al: Fluid Accumulation in Mechanically Ventilated, Critically Ill Children: Retrospective Cohort Study of Prevalence and Outcome. Pediatr Crit Care Med 2022; 23(12):990–998

49. National Heart L, Blood Institute Acute Respiratory Distress Syndrome Clinical Trials N, Wiedemann HP, et al: Comparison of two fluid-management strategies in acute lung injury. N Engl J Med 2006; 354(24):2564–2575

50. Gamble KC, Smith SE, Bland CM, et al: Hidden Fluids in Plain Sight: Identifying Intravenous Medication Classes as Contributors to Intensive Care Unit Fluid Intake. Hosp Pharm 2022; 57(2):230–236

51. Branan T, Smith SE, Newsome AS, et al: Association of hidden fluid administration with development of fluid overload reveals opportunities for targeted fluid minimization. SAGE Open Med 2020; 8:2050312120979464

52. Mitchell KH, Carlbom D, Caldwell E, et al: Volume Overload: Prevalence, Risk Factors, and Functional Outcome in Survivors of Septic Shock. Ann Am Thorac Soc 2015; 12(12):1837–1844

53. Ouchi A, Sakuramoto H, Hoshino H, et al: Association between fluid overload and delirium/coma in mechanically ventilated patients. Acute Med Surg 2020; 7(1):e508

54. Murphy CV, Schramm GE, Doherty JA, et al: The importance of fluid management in acute lung injury secondary to septic shock. Chest 2009; 136(1):102–109

55. Boyd JH, Forbes J, Nakada TA, et al: Fluid resuscitation in septic shock: a positive fluid balance and elevated central venous pressure are associated with increased mortality. Crit Care Med 2011; 39(2):259–265

56. Woodward CW, Lambert J, Ortiz-Soriano V, et al: Fluid Overload Associates With Major Adverse Kidney Events in Critically Ill Patients With Acute Kidney Injury Requiring Continuous Renal Replacement Therapy. Crit Care Med 2019; 47(9):e753–e760

57. Hawkins WA, Butler SA, Poirier N, et al: From theory to bedside: Implementation of fluid stewardship in a medical ICU pharmacy practice. Am J Health Syst Pharm 2022; 79(12):984–992

58. Silversides JA, Perner A, Malbrain M: Liberal versus restrictive fluid therapy in critically ill patients. Intensive Care Med 2019; 45(10):1440–1442

59. Goldstein S, Bagshaw S, Cecconi M, et al: Pharmacological management of fluid overload. Br J Anaesth 2014; 113(5):756–763

60. Silversides JA, McAuley DF, Blackwood B, et al: Fluid management and deresuscitation practices: A survey of critical care physicians. J Intensive Care Soc 2020; 21(2):111–118

61. Burkov A: The hundred-page machine learning book. Quebec City, Canada, Andriy Burkov, 2019

62. Qin X, Zhang W, Hu X, et al: A deep learning model to identify the fluid overload status in critically ill patients based on chest X-ray images. Pol Arch Intern Med 2023; 133(2)

## Bibliography

1. Bouchard J, Soroko SB, Chertow GM, et al. Fluid accumulation, survival and recovery of kidney function in critically ill patients with acute kidney injury. Kidney Int 2009;76(4):422–7. (In eng). DOI: 10.1038/ki.2009.159.

2. Carr JR, Hawkins WA, Newsome AS, et al. Fluid Stewardship of Maintenance Intravenous Fluids. J Pharm Pract 2022;35(5):769–782. (In eng). DOI: 10.1177/08971900211008261.

3. Hawkins WA, Smith SE, Newsome AS, Carr JR, Bland CM, Branan TN. Fluid Stewardship During Critical Illness: A Call to Action. J Pharm Pract 2020;33(6):863–873. (In eng). DOI: 10.1177/0897190019853979.

4. Bissell BD, Donaldson JC, Morris PE, Neyra JA. A narrative review of pharmacologic de-resuscitation in the critically ill. J Crit Care 2020;59:156–162. (In eng). DOI: 10.1016/j.jcrc.2020.07.004.

5. Bissell BD, Laine ME, Thompson Bastin ML, et al. Impact of protocolized diuresis for de-resuscitation in the intensive care unit. Crit Care 2020;24(1):70. (In eng). DOI: 10.1186/s13054-020-2795-9.

6. Hawkins WA, Butler SA, Poirier N, Wilson CS, Long MK, Smith SE. From theory to bedside: Implementation of fluid stewardship in a medical ICU pharmacy practice. Am J Health Syst Pharm 2022;79(12):984–992. (In eng). DOI: 10.1093/ajhp/zxab453.

7. Jones TW, Chase AM, Bruning R, Nimmanonda N, Smith SE, Sikora A. Early Diuretics for De-resuscitation in Septic Patients With Left Ventricular Dysfunction. Clin Med Insights Cardiol 2022;16:11795468221095875. (In eng). DOI: 10.1177/11795468221095875.

8. Olney WJ, Chase AM, Hannah SA, Smith SE, Newsome AS. Medication Regimen Complexity Score as an Indicator of Fluid Balance in Critically Ill Patients. J Pharm Pract 2022;35(4):573–579. (In eng). DOI: 10.1177/0897190021999792.

9. Rafiei A, Rad MG, Sikora A, Kamaleswaran R. Improving irregular temporal modeling by integrating synthetic data to the electronic medical record using conditional GANs: a case study of fluid overload prediction in the intensive care unit. medRxiv 2023:2023.06.20.23291680. DOI: 10.1101/2023.06.20.23291680.

10. Sikora A, Zhang T, Murphy DJ, et al. Machine learning vs. traditional regression analysis for fluid overload prediction in the ICU. medRxiv 2023:2023.06.16.23291493. DOI: 10.1101/2023.06.16.23291493.

11. Messmer AS, Moser M, Zuercher P, Schefold JC, Müller M, Pfortmueller CA. Fluid Overload Phenotypes in Critical Illness-A Machine Learning Approach. J Clin Med 2022;11(2) (In eng). DOI: 10.3390/jcm11020336.

12. Zhang Z, Ho KM, Hong Y. Machine learning for the prediction of volume responsiveness in patients with oliguric acute kidney injury in critical care. Crit Care 2019;23(1):112. (In eng). DOI: 10.1186/s13054-019-2411-z.

13. Sikora A, Devlin JW, Yu M, et al. Evaluation of medication regimen complexity as a predictor for mortality. Sci Rep 2023;13(1):10784. DOI: 10.1038/s41598-023-37908-1.

14. Gwynn ME, Poisson MO, Waller JL, Newsome AS. Development and validation of a medication regimen complexity scoring tool for critically ill patients. Am J Health Syst Pharm 2019;76(Supplement_2):S34–s40. (In eng). DOI: 10.1093/ajhp/zxy054.

15. Murray B, Zhao B, Kong Y, Shen Y, Sikora A. 935: PREDICTING DURATION OF MECHANICAL VENTILATION WITH MEDICATION REGIMEN COMPLEXITY VARIABLES. Critical Care Medicine 2023;51(1):460. DOI: 10.1097/01.ccm.0000909468.03569.76.

16. Webb AJ, Rowe S, Sikora Newsome A. A descriptive report of the rapid implementation of automated MRC-ICU calculations in the EMR of an academic medical center. Am J Health Syst Pharm 2022. DOI: 10.1093/ajhp/zxac059.

17. Sikora A, Rafiei A, Rad MG, et al. Pharmacophenotype identification of intensive care unit medications using unsupervised cluster analysis of the ICURx common data model. Crit Care 2023;27(1):167. DOI: 10.1186/s13054-023-04437-2.

18. Chase A AH, Forehand C, Keats K, Taylor A, Wu S, Blotske K, Sikora A. An evaluation of medication regimen complexity’s relationship to medication errors in critically ill patients. Hospital Pharmacy 2023;Accepted.

19. Al-Mamun MA, Brothers T, Newsome AS. Development of Machine Learning Models to Validate a Medication Regimen Complexity Scoring Tool for Critically Ill Patients. Ann Pharmacother 2021;55(4):421–429. (In eng). DOI: 10.1177/1060028020959042.

20. Newsome AS, Anderson D, Gwynn ME, Waller JL. Characterization of changes in medication complexity using a modified scoring tool. Am J Health Syst Pharm 2019;76(Supplement_4):S92–s95. (In eng). DOI: 10.1093/ajhp/zxz213.

21. Sikora A, Ayyala D, Rech MA, et al. Impact of Pharmacists to Improve Patient Care in the Critically Ill: A Large Multicenter Analysis Using Meaningful Metrics With the Medication Regimen Complexity-ICU (MRC-ICU) Score. Crit Care Med 2022;50(9):1318–1328. (In eng). DOI: 10.1097/ccm.0000000000005585.

22. Sikora A, Rafiei A, Rad MG, et al. Pharmacophenotype identification of intensive care unit medications using unsupervised cluster analysis of the ICURx common data model. Crit Care 2023;27(1):167. (In eng). DOI: 10.1186/s13054-023-04437-2.

23. Sikora A JH, Yu M, Chen X, Murray B, Kamaleswaran R. Cluster analysis driven by unsupervised latent feature learning of intensive care unit medications to identify novel pharmacophenotypes of critically ill patients. Research Square 2022. DOI: 10.21203/rs.3.rs-1745568/v1.

24. World Medical Association Declaration of Helsinki: ethical principles for medical research involving human subjects. Jama 2013;310(20):2191–4. (In eng). DOI: 10.1001/jama.2013.281053.

25. Elm Ev, Altman DG, Egger M, Pocock SJ, Gøtzsche PC, Vandenbroucke JP. Strengthening the reporting of observational studies in epidemiology (STROBE) statement: guidelines for reporting observational studies. BMJ 2007;335(7624):806–808. DOI: 10.1136/bmj.39335.541782.AD.

26. Sikora A, Keats K, Murphy DJ, et al. A Common Data Model for the standardization of intensive care unit (ICU) medication features in artificial intelligence (AI) applications. medRxiv 2023:2023.09.18.23295727. DOI: 10.1101/2023.09.18.23295727.

27. Bro R, Smilde AK. Principal component analysis. Analytical Methods 2014;6(9):2812–2831. (10.1039/C3AY41907J). DOI: 10.1039/C3AY41907J.

28. Wold S, Esbensen K, Geladi P. Principal component analysis. Chemometrics and Intelligent Laboratory Systems 1987;2(1):37–52. DOI: 10.1016/0169-7439(87)80084-9.

29. Hinton GE. A Practical Guide to Training Restricted Boltzmann Machines. In: Montavon G, Orr GB, Müller K-R, eds. Neural Networks: Tricks of the Trade: Second Edition. Berlin, Heidelberg: Springer Berlin Heidelberg; 2012:599-619.

30. Zhang N, Ding S, Zhang J, Xue Y. An overview on Restricted Boltzmann Machines. Neurocomputing 2018;275:1186–1199. DOI: 10.1016/j.neucom.2017.09.065.

31. Deng S. Syuand/unsupervisedfoprediction-: A time-dependent, unsupervised machine learning (ML) analysis was conducted to determine patterns of IV medication use associated with fo. GitHub. (https://github.com/SyuanD/UnsupervisedFOPrediction-git).

32. Rafiei A, Rad MG, Sikora A, Kamaleswaran R. Improving irregular temporal modeling by integrating synthetic data to the electronic medical record using conditional GANs: a case study of fluid overload prediction in the intensive care unit. medRxiv 2023. DOI: 10.1101/2023.06.20.23291680.

33. Sikora A, Zhang T, Murphy DJ, et al. Machine learning vs. traditional regression analysis for fluid overload prediction in the ICU. medRxiv 2023:2023.06.16.23291493. DOI: 10.1101/2023.06.16.23291493.

34. Olney WJ, Chase AM, Hannah SA, Smith SE, Newsome AS. Medication Regimen Complexity Score as an Indicator of Fluid Balance in Critically Ill Patients. J Pharm Pract 2021:897190021999792. DOI: 10.1177/0897190021999792.

35. Sikora A, Zhao B, Kong Y, Murray B, Shen Y. Machine learning based prediction of prolonged duration of mechanical ventilation incorporating medication data. medRxiv 2023. DOI: 10.1101/2023.09.18.23295724.

36. Sikora A, Keats K, Murphy DJ, et al. A Common Data Model for the standardization of intensive care unit (ICU) medication features in artificial intelligence (AI) applications. medRxiv 2023:2023.09.18.23295727. DOI: 10.1101/2023.09.18.23295727.

37. Goh KH, Wang L, Yeow AYK, et al. Artificial intelligence in sepsis early prediction and diagnosis using unstructured data in healthcare. Nat Commun 2021;12(1):711. (In eng). DOI: 10.1038/s41467-021-20910-4.

38. Gutierrez G. Artificial Intelligence in the Intensive Care Unit. Crit Care 2020;24(1):101. (In eng). DOI: 10.1186/s13054-020-2785-y.

39. Hyland SL, Faltys M, Hüser M, et al. Early prediction of circulatory failure in the intensive care unit using machine learning. Nat Med 2020;26(3):364–373. (In eng). DOI: 10.1038/s41591-020-0789-4.

40. Iwase S, Nakada TA, Shimada T, et al. Prediction algorithm for ICU mortality and length of stay using machine learning. Sci Rep 2022;12(1):12912. (In eng). DOI: 10.1038/s41598-022-17091-5.

41. Beil M, Sviri S, Flaatten H, et al. On predictions in critical care: The individual prognostication fallacy in elderly patients. J Crit Care 2021;61:34–38. (In eng). DOI: 10.1016/j.jcrc.2020.10.006.

42. Lovejoy CA, Buch V, Maruthappu M. Artificial intelligence in the intensive care unit. Crit Care 2019;23(1):7. (In eng). DOI: 10.1186/s13054-018-2301-9.

